# The Urban Divide: Socioeconomic Status and Its Impact on Cancer in the Age of Urbanization

**DOI:** 10.1101/2025.08.17.25333879

**Authors:** Yuriko N. Koyanagi, Masanori Kawaura, Keitaro Matsuo, Yukino Kawakatsu, Yukari Taniyama, Isao Oze, Takahiro Otani, Kunihiko Takahashi, Rui Yamaguchi, Tomoki Nakaya, Hidemi Ito

## Abstract

Against accelerating global urbanization, we analyzed regional cancer health disparities in Japan using nationwide cancer incidence and mortality data. This cross-sectional ecological study included 1889 Japanese municipalities, categorized as urban, suburban, and rural. Using 2015 Census data, we estimated the normalized Areal Deprivation Index (normADI) and, using 2016-2018 cancer registry and vital statistics data, estimated standardized incidence and mortality ratios with a fully Bayesian hierarchical spatial model (FBSIR/FBSMR). We examined their association by urbanicity, analyzing 3,056,535 incident cases and 1,140,250 deaths. Higher normADI was associated with increased FBSMR for all cancers, particularly in urban areas (risk ratio 1.18 for males; 1.12 for females).

Mortality of stomach, colorectal, lung, and liver cancers increased with increasing deprivation in urban areas, with stronger effects for male colorectal, liver, and lung cancers as urbanicity increased. Sensitivity analyses using alternative modeling formulations, both spatial and non-spatial, yielded broadly similar results. The greater impact of ADI on cancer mortality in urban areas underscores the need for poverty-focused cancer prevention strategies with growing urban populations.

---

The worldwide shift from rural to urban living marks one of the most significant transformations affecting human societies. Over half of the global population is now thought to reside in urban areas, and this ratio is expected to increase.^1^ This urbanization brings not only opportunity but also a complex range of health challenges. Among these, the disparities in health outcomes based on socioeconomic status are clear.

Socioeconomic-based health disparities are widely recognized,^2–4^ extending from general health outcomes into the realm of cancer. Globally, incidence and mortality rates for most cancers are disproportionately higher among lower socioeconomic groups.^5^ Furthermore, a clear pattern of geographic disparities in health outcomes has emerged in both Western countries^6–10^ and Japan^11–13^. A number of studies have indicated that the association between areal socioeconomic deprivation and several health outcomes (e.g. premature mortality^14^, premature limiting long-term illness^15^, survival^16^, and all-cause mortality^17^) is more pronounced in areas of high urbanicity, suggesting that the forces driving health disparities in urban settings may be distinct, and potentially more severe than those in less urbanized areas. Nevertheless, few studies have investigated the impact of urbanization on health disparities, focusing on cancer outcomes, and results have been inconsistent.^18–21^

Here, we investigated the association between areal socioeconomic status and cancer incidence/mortality, with a particular focus on the role of urbanicity. In particular, we used data from the nationwide population-based cancer registry and vital statistics to identify potential heterogeneity in the impact of areal socioeconomic status on cancer burden within different levels of urbanicity.

## Results

### Characteristics of municipalities by urbanicity

We performed a cross-sectional ecological study of 1889 municipal geographic units in Japan. Of the 1889 municipalities, 201, 627, and 1061 were categorized as urban, suburban, and rural, respectively (Supplementary Figure 1). Urban municipalities had a higher percentage of young population (age <40) and a lower percentage of aged population (age ≥75) compared with suburban and rural municipalities (Table 1).

**Table 1.** Characteristics of municipalities in Japan by urbanicity (2015)

| Characteristic | Urban (*n*<sub>municipality</sub> = 201) |  | Suburban (*n*<sub>municipality</sub> = 627) |  | Rural (*n*<sub>municipality</sub> = 1,061) |  |
| --- | --- | --- | --- | --- | --- | --- |
|  | Male | Female | Male | Female | Male | Female |
| **Population (*n*)** | 18,355,035 | 19,422,844 | 24,533,744 | 25,631,885 | 18,123,729 | 19,572,762 |
| **Population Density (Persons/km<sup>2</sup>)** | 1308.9 | 1385.0 | 340.9 | 356.2 | 63.3 | 68.4 |
| **Age distribution (*n*, [%])** |  |  |  |  |  |  |
| **<40 years** | 8,032,257<br>[44%] | 7,876,303<br>[41%] | 10,298,472<br>[42%] | 9,845,396<br>[38%] | 7,079,129<br>[39%] | 6,748,258<br>[34%] |
| **40-<75 years** | 8,682,261<br>[47%] | 8,936,728<br>[46%] | 11,823,597<br>[48%] | 12,156,543<br>[47%] | 8,853,471<br>[49%] | 9,181,941<br>[47%] |
| **75- years** | 1,640,517<br>[9%] | 2,609,813<br>[13%] | 2,411,675<br>[10%] | 3,629,946<br>[14%] | 2,191,129<br>[12%] | 3,642,563<br>[19%] |
| **Mean normADI [SD]** | 0.44 [ 0.31 ] |  | 0.55 [ 0.28 ] |  | 0.78 [ 0.23 ] |  |
| **Median normADI [IQR]** | 0.42 [ 0.16 - 0.71 ] |  | 0.56 [ 0.33 - 0.80 ] |  | 0.85 [ 0.66 - 0.96 ] |  |

Derived from Census data for the year 2015.
ADI, Areal Deprivation Index; normADI, normalized ADI; SD, standard deviation; IQR, interquartile range.

### Cancer incidence and mortality

Using data from the population-based National Cancer Registry from 2016 to 2018 and Vital Statistics for 2016 to 2018, a total of 3,056,535 cancer incident cases and 1,140,250 cancer deaths were included in this study (Supplementary Figure 2). The number, crude rate per 100,000 population, and age-standardized rate of cancer incident cases and deaths by urbanicity and cancer site are shown in Table 2. The distribution of individual cancer sites remained largely consistent across urban, suburban, and rural areas (Supplementary Figure 3), suggesting no substantial variation in their proportional contributions. The cancer profile with the highest rate remained largely consistent across urbanicity levels. Specifically, the most common cancer sites were stomach or colorectum for males and breast for females for cancer incidence, and lung for males and colorectum for females for cancer death. The mean (standard deviation [SD]) and median (interquartile range [IQR]) of the standardized incidence/mortality ratios (SIR/SMR) and fully Bayesian SIR/SMR (FBSIR/FBSMR) – Bayesian estimates of regional relative risk of incidence/mortality – by urbanicity in Japan (2016-18) are shown in Supplementary Tables 1 and 2. To stabilize risk estimates and account for spatial dependencies, we used FBSIR/FBSMR, derived from the Besag-York-Mollié 2 (BYM2) model^22^, a fully Bayesian approach incorporating spatially structured and unstructured random effects, as the outcome indicator. For all cancers and most cancer sites, the mean FBSIR/FBSMR values in urban areas were above unity, whereas those in suburban areas were at or below unity and those in rural areas were below unity.

**Table 2.** Cancer incidence and mortality in Japan according to urbanicity (2016-18)

| Site | ICD-10 | Urban |  | CR | ASR | Suburban |  | CR | ASR | Rural |  | CR | ASR |
| --- | --- | --- | --- | --- | --- | --- | --- | --- | --- | --- | --- | --- | --- |
|  |  | *n* | % |  |  | *n* | % |  |  | *n* | % |  |  |
| **Male** |  |  |  |  |  |  |  |  |  |  |  |  |  |
| **Incidence<sup>a</sup>** |  |  |  |  |  |  |  |  |  |  |  |  |  |
| All | C00-C96* | 493,214 | 100% | 895.7 | 355.1 | 685,764 | 100% | 931.7 | 337.0 | 565,900 | 100% | 1,040.8 | 337.7 |
| Stomach | C16 | 73,280 | 14.9% | 133.1 | 49.0 | 106,788 | 15.6% | 145.1 | 49.0 | 90,583 | 16.0% | 166.6 | 50.8 |
| Colorectum | C18-C20 | 77,734 | 15.8% | 141.2 | 58.2 | 106,130 | 15.5% | 144.2 | 55.0 | 88,175 | 15.6% | 162.2 | 55.9 |
| Lung | C33-C34 | 72,119 | 14.6% | 131.0 | 47.9 | 100,154 | 14.6% | 136.1 | 45.0 | 84,033 | 14.8% | 154.6 | 45.5 |
| Liver | C22 | 23,804 | 4.8% | 43.2 | 16.3 | 32,262 | 4.7% | 43.8 | 14.9 | 26,579 | 4.7% | 48.9 | 15.0 |
| Pancreas | C25 | 18,008 | 3.7% | 32.7 | 12.7 | 25,261 | 3.7% | 34.3 | 12.2 | 21,136 | 3.7% | 38.9 | 12.2 |
| Breast | C50 |  |  |  |  |  |  |  |  |  |  |  |  |
| Prostate | C61 | 75,659 | 15.3% | 137.4 | 48.5 | 109,982 | 16.0% | 149.4 | 47.5 | 88,592 | 15.7% | 162.9 | 46.1 |
| Cervix | C53 |  |  |  |  |  |  |  |  |  |  |  |  |
| **Mortality<sup>b</sup>** |  |  |  |  |  |  |  |  |  |  |  |  |  |
| All | C00-C97* | 184,336 | 100% | 334.8 | 114.5 | 261,593 | 100% | 355.4 | 111.0 | 225,062 | 100% | 413.9 | 111.5 |
| Stomach | C16 | 23,440 | 12.7% | 42.6 | 14.2 | 35,401 | 13.5% | 48.1 | 14.7 | 29,887 | 13.3% | 55.0 | 14.6 |
| Colorectum | C18-C20 | 22,711 | 12.3% | 41.2 | 14.9 | 31,445 | 12.0% | 42.7 | 14.2 | 27,648 | 12.3% | 50.9 | 14.7 |
| Lung | C33-C34 | 43,803 | 23.8% | 79.5 | 26.3 | 62,000 | 23.7% | 84.2 | 25.3 | 52,732 | 23.4% | 97.0 | 25.1 |
| Liver | C22 | 15,190 | 8.2% | 27.6 | 9.4 | 21,166 | 8.1% | 28.8 | 8.9 | 17,420 | 7.7% | 32.0 | 8.8 |
| Pancreas | C25 | 14,384 | 7.8% | 26.1 | 9.5 | 20,496 | 7.8% | 27.8 | 9.3 | 17,711 | 7.9% | 32.6 | 9.5 |
| Breast | C50 |  |  |  |  |  |  |  |  |  |  |  |  |
| Prostate | C61 | 9,622 | 5.2% | 17.5 | 4.8 | 13,856 | 5.3% | 18.8 | 4.7 | 12,693 | 5.6% | 23.3 | 4.6 |
| Cervix | C53 |  |  |  |  |  |  |  |  |  |  |  |  |
| **Female** |  |  |  |  |  |  |  |  |  |  |  |  |  |
| **Incidence<sup>a</sup>** |  |  |  |  |  |  |  |  |  |  |  |  |  |
| All | C00-C96* | 387,535 | 100% | 665.1 | 282.5 | 502,475 | 100% | 653.5 | 262.2 | 421,647 | 100% | 718.1 | 260.1 |
| Stomach | C16 | 33,611 | 8.7% | 57.7 | 17.8 | 46,726 | 9.3% | 60.8 | 17.7 | 42,015 | 10.0% | 71.6 | 18.1 |
| Colorectum | C18-C20 | 59,853 | 15.4% | 102.7 | 35.3 | 78,818 | 15.7% | 102.5 | 33.3 | 68,148 | 16.2% | 116.1 | 32.7 |
| Lung | C33-C34 | 38,621 | 10.0% | 66.3 | 21.3 | 48,095 | 9.6% | 62.5 | 18.7 | 41,249 | 9.8% | 70.2 | 18.6 |
| Liver | C22 | 11,151 | 2.9% | 19.1 | 5.0 | 15,665 | 3.1% | 20.4 | 4.8 | 13,438 | 3.2% | 22.9 | 4.7 |
| Pancreas | C25 | 17,195 | 4.4% | 29.5 | 8.6 | 23,007 | 4.6% | 29.9 | 8.2 | 21,184 | 5.0% | 36.1 | 8.3 |
| Breast | C50 | 91,712 | 23.7% | 157.4 | 86.3 | 113,506 | 22.6% | 147.6 | 78.6 | 84,757 | 20.1% | 144.3 | 74.9 |
| Prostate | C61 |  |  |  |  |  |  |  |  |  |  |  |  |
| Cervix | C53 | 10,342 | 2.7% | 17.7 | 11.4 | 13,019 | 2.6% | 16.9 | 10.9 | 9,993 | 2.4% | 17.0 | 11.3 |
| **Mortality<sup>b</sup>** |  |  |  |  |  |  |  |  |  |  |  |  |  |
| All | C00-C97* | 131,403 | 100% | 225.5 | 64.2 | 176,413 | 100% | 229.4 | 61.5 | 161,443 | 100% | 274.9 | 61.0 |
| Stomach | C16 | 12,111 | 9.2% | 20.8 | 5.3 | 17,812 | 10.1% | 23.2 | 5.5 | 16,796 | 10.4% | 28.6 | 5.6 |
| Colorectum | C18-C20 | 19,357 | 14.7% | 33.2 | 8.7 | 26,107 | 14.8% | 34.0 | 8.3 | 24,851 | 15.4% | 42.3 | 8.4 |
| Lung | C33-C34 | 19,354 | 14.7% | 33.2 | 8.3 | 24,354 | 13.8% | 31.7 | 7.3 | 21,239 | 13.2% | 36.2 | 6.8 |
| Liver | C22 | 7,749 | 5.9% | 13.3 | 2.8 | 10,870 | 6.2% | 14.1 | 2.8 | 9,806 | 6.1% | 16.7 | 2.8 |
| Pancreas | C25 | 14,166 | 10.8% | 24.3 | 6.2 | 18,871 | 10.7% | 24.5 | 5.9 | 17,910 | 11.1% | 30.5 | 6.0 |
| Breast | C50 | 13,456 | 10.2% | 23.1 | 10.0 | 16,796 | 9.5% | 21.8 | 9.1 | 13,036 | 8.1% | 22.2 | 8.7 |
| Prostate | C61 |  |  |  |  |  |  |  |  |  |  |  |  |
| Cervix | C53 | 2,412 | 1.8% | 4.1 | 2.0 | 3,340 | 1.9% | 4.3 | 2.1 | 2,731 | 1.7% | 4.7 | 2.1 |

Derived from the population-based National Cancer Registry from 2016 to 2018<sup>a</sup> and the Vital Statistics for 2016 to 2018<sup>b</sup>.
\*Invasive blood cancers (D45,D46,D460,D461,D462,D464,D465,D466,D467,D469,D471,D473,D474,D475) were also included.
ICD-10, International Statistical Classification of Diseases and Related Health Problems; CR, Crude rate per 100,000 population; ASR, age-standardized rate.

### Geography of FBSIR/FBSMR for all cancers and by cancer site

To simplify geographic descriptions, we established several definitions by defining three densely populated areas – Tokyo, Nagoya, and Osaka –as metropolitan areas, and the islands comprising the Japanese archipelago as Hokkaido, Honshu, Shikoku, Kyushu, and Okinawa (Supplementary Figure 4). Figure 1 presents choropleth maps of FBSIR, FBSMR, and their exceedance probabilities for all cancers by sex. High FBSIR for all cancers among males was observed in Japan’s northern coastal areas, as well as in the Kanto (Tokyo area) and Kansai (Osaka area) regions and Hiroshima. FBSMR was higher in Hokkaido, the northern part of Honshu and Kyushu, and the Tokyo, Nagoya, and Osaka metropolitan areas for both males and females. Notably, exceedance probabilities were also high in these regions, reinforcing the consistency of these findings.

**Figure 1.**
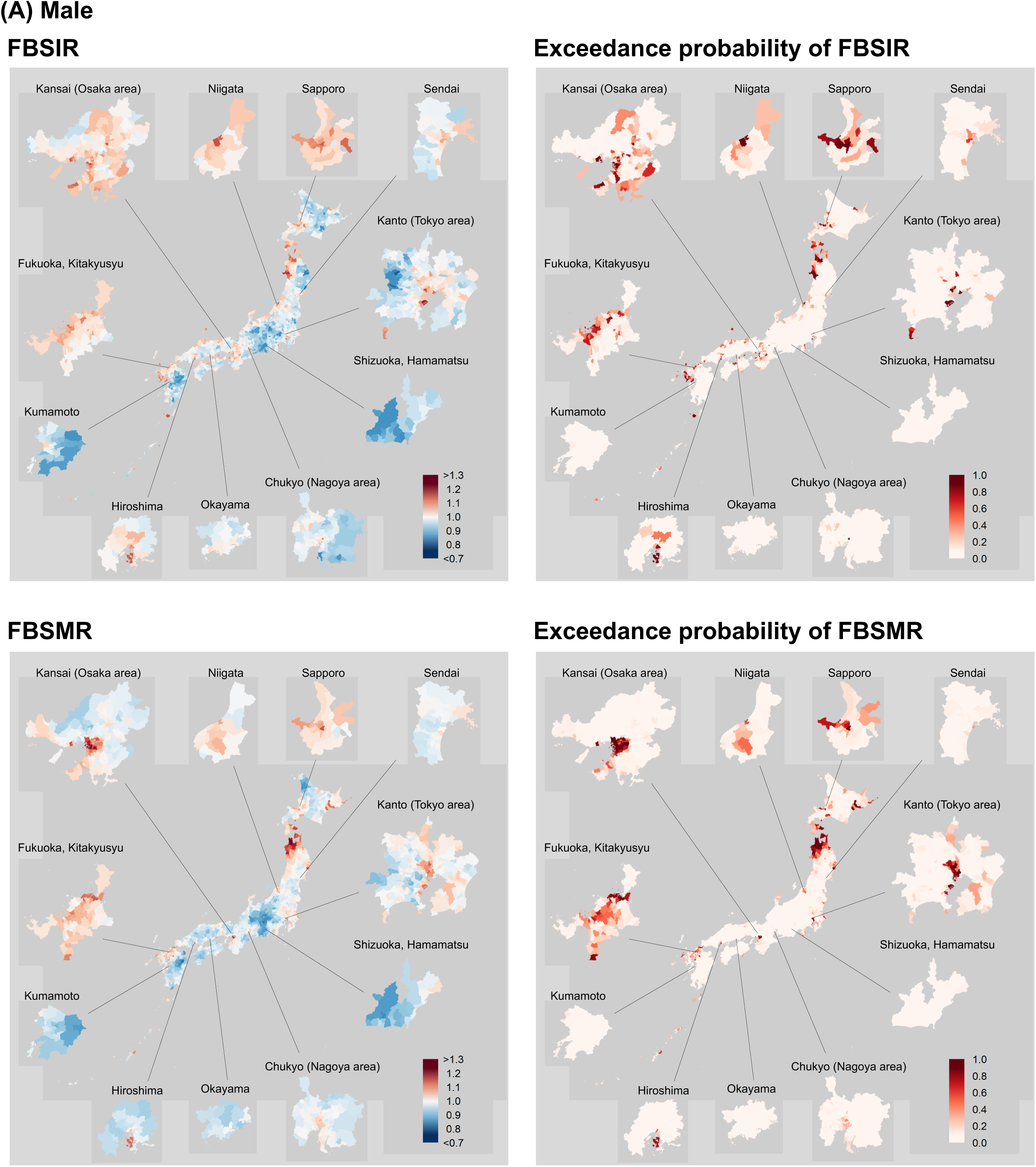
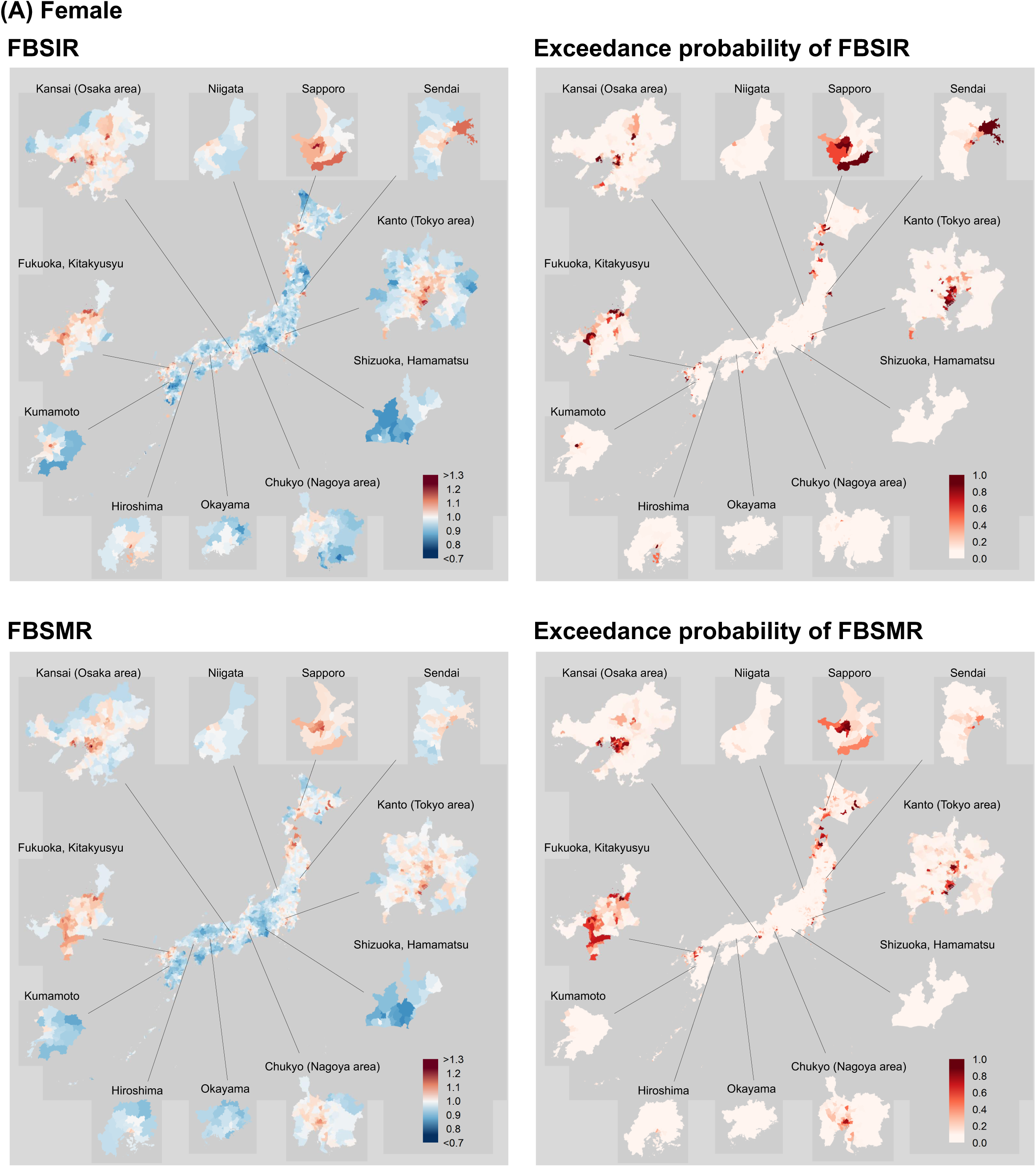
Geographic Distribution of FBSIR/FBSMR and Their Exceedance Probabilities by Sex and Municipality for All Cancers. Color-coded maps of FBSIR and FBSMR (ranging from red [high] to blue [low]) and their exceedance probabilities of all cancers are shown for males (A) and females (B). The exceedance probabilities represent the probabilities of FBSIR/FBSMR values being greater than 1.1 within individual municipalities. Major central cities and their surrounding areas are broken out. As this study follows census-based regional definitions for consistency, some discrepancies between municipality names and their corresponding prefectures arise (e.g.,’Hiroshima’ refers to the Hiroshima metropolitan area rather than Hiroshima Prefecture). FBSIR, fully Bayesian estimate of standardized incidence ratio; FBSMR, fully Bayesian estimate of standardized mortality ratio.

Site-specific analyses revealed distinct geographic distributions in both FBSIR and FBSMR (Supplementary Figure 5). For stomach and colorectal cancers, we observed higher FBSIR/FBSMR in eastern Japan, especially in the northern coastal areas (for stomach) and the northern part of Honshu. Pancreatic cancer showed the same tendency as stomach and colorectal cancers, but the absolute values were relatively low, indicating only weak differences in pancreatic cancer incidence/mortality among municipalities. In contrast, higher FBSIR/FBSMR was observed in western Japan for liver cancer. For lung cancer, lower FBSIR/FBSMR was observed in central Japan in males, whereas higher FBSIR/FBSMR was observed in major central cities for both sexes. For breast cancer, higher FBSIR/FBSMR values were found in major central cities and surrounding cities, especially in Kanto (Tokyo area). FBSIR for prostate cancer had an uncharacteristic distribution, but FBSMR appeared to be higher in eastern Japan, although absolute values were relatively low. For cervical cancer, municipalities with high FBSIR and FBSMR values are scattered across Japan’s Pacific coastal regions, including major central cities (Tokyo, Nagoya, Osaka, and Fukuoka) and surrounding cities and in Hokkaido. Again, in regions with high FBSIR and FBSMR, the exceedance probabilities were also elevated, further supporting the robustness of our findings.

### Geography of the normalized Area Deprivation Index (normADI)

We assessed area-level socioeconomic status using the normADI^17,23,24^, estimated with data from Census^25^ data for 2015 (Census 2015). The mapping of normADI across Japan municipalities is shown in Figure 2. Two geographic disparities can be recognized: (1) the central and peripheral areas of Japan as a whole, and (2) inner city and suburban areas within metropolitan areas. Specifically, the low normADI (less deprived) areas were located mainly in the urban areas in the center of the main island (Honshu), while the high normADI (more deprived) areas were located in the geographically peripheral regions of Honshu and other islands. High normADI areas were found to exist within metropolitan areas (inner city-like neighborhoods), such as the eastern and northern parts of Tokyo, and the central parts of the Nagoya and Osaka metropolitan areas. These inner city-like neighborhoods were surrounded by suburban areas consisting of large metropolitan areas with low normADI. The distribution of normADI by urbanicity is shown in Figure 3. The mean (SD) and median (IQR) normADI values were 0.440 (0.308) and 0.420 (0.164-0.704) for urban areas, 0.552 (0.281) and 0.555 (0.332-0.799) for suburban areas, and 0.776 (0.226) and 0.850 (0.658-0.961) for rural areas (Table 1). Consequently, urban areas exhibited lower normADI values with greater regional variability, whereas rural areas demonstrated higher normADI values with lesser regional variability.

**Figure 2.**
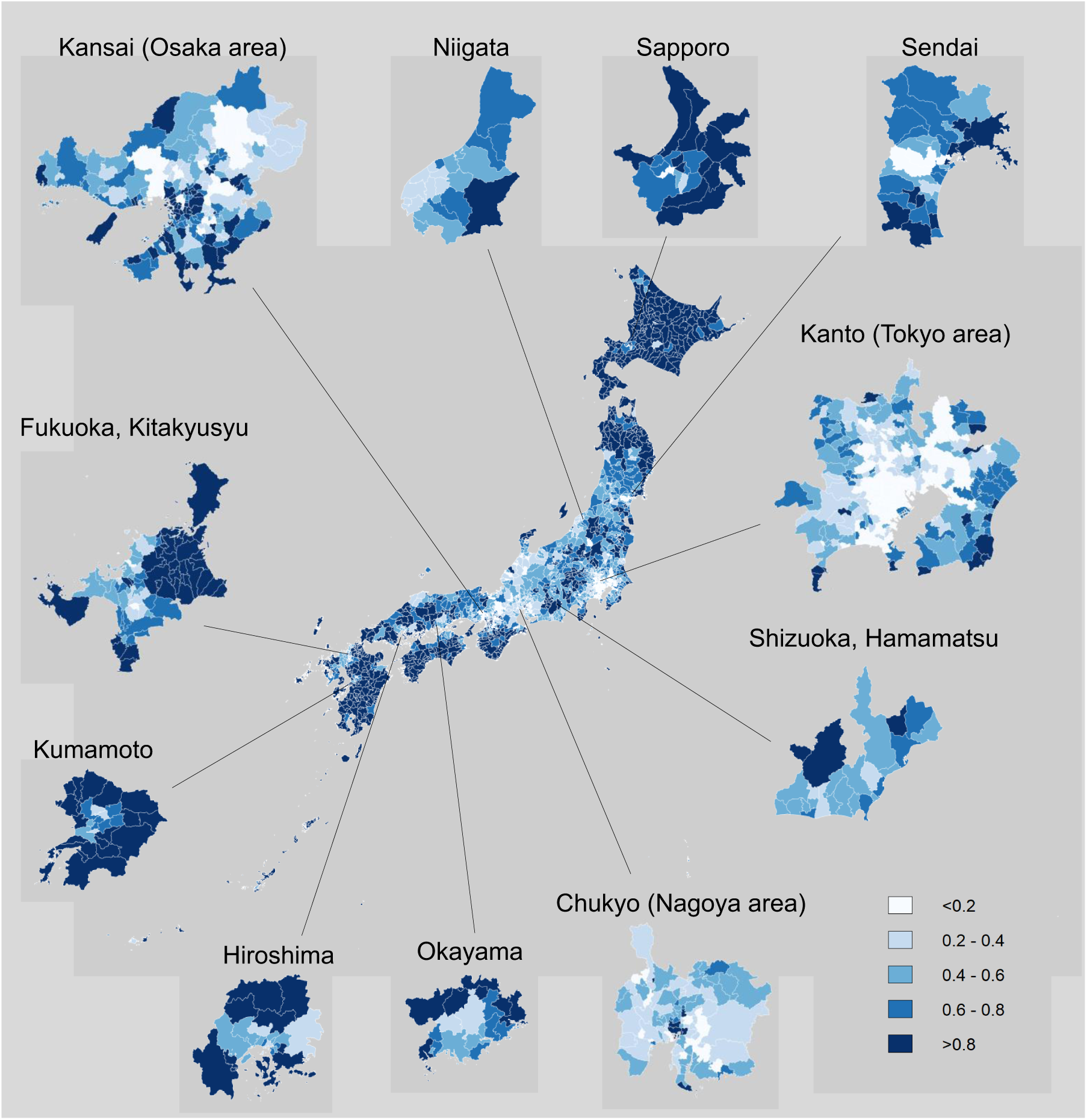
Map of normADI in 2015. Normalized ADI (normADI) represents the relative position of a municipality among all municipalities in Japan, ranging from zero (least deprived) to one (most deprived). ADI, Areal Deprivation Index. Major central cities and their surrounding areas are broken out.

**Figure 3.**
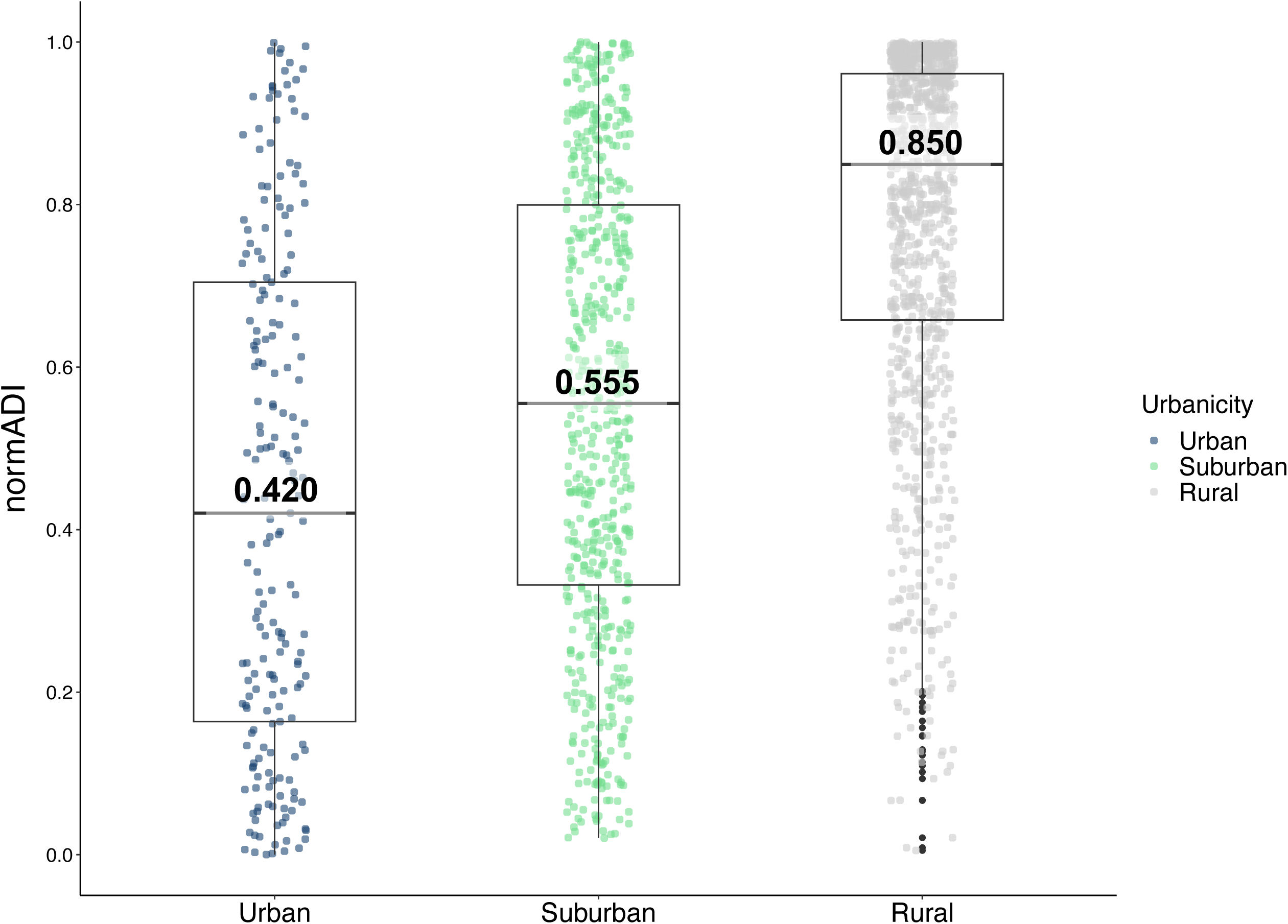
Differences in normADI by Urbanicity Boxplot showing the distribution of normADI by urbanicity. normADI, normalized areal deprivation index.

### Impact of normADI on cancer incidence and mortality

The impact of normADI of municipalities on FBSIR/FBSMR, using risk ratio (RR) and its 95% credible interval (CR) as the measure, is demonstrated in Figure 4. Greater deprivation was associated with increased FBSMR for all cancers particularly in urban areas (males: RR_urban_ = 1.18 [95%CR: 1.14-1.22]; females: RR_urban_ = 1.12 [95%CR: 1.07-1.17]), with heterogeneity in this association observed across levels of urbanicity (heterogeneity RR for suburban vs. urban [Het RR_suburban_], males: 0.93 [95%CR: 0.89-0.97]; females: 0.93 [95%CR: 0.88-0.97]; Het RR for rural vs. urban [Het RR_rural_], males: 0.90 [95%CR: 0.85-0.94]; females: 0.92 [95%CR: 0.87-0.98]). By cancer site, stomach cancer FBSIR tended to be lower in regions with greater deprivation, and this effect was more pronounced in less urbanized areas. However, this association was attenuated in FBSMR, and in urban areas, greater deprivation was associated with increased FBSMR, especially in males (RR_urban_ = 1.16 [95%CR: 1.06-1.26]). For colorectal cancer, FBSIR increased with deprivation in urban areas for both sexes (males: RR_urban_ = 1.13 [95%CR: 1.07-1.20]; females: RR_urban_ = 1.08 [95%CR: 1.01-1.15]), while an inverse association was observed in suburban and rural areas for females. FBSMR showed a positive association with ADI in urban areas (males: RR_urban_ = 1.26 [95%CR: 1.17-1.36]; females: RR_urban_ = 1.12 [95%CR: 1.04-1.21]); heterogeneity due to urbanicity was evident for both sexes, with the effect of deprivation being stronger as urbanicity increased for males. For lung and liver cancers, greater deprivation showed an increased FBSIR/FBSMR. This association was stronger with higher urbanicity, especially in male mortality (lung: RR_urban_ = 1.27 [95%CR: 1.19-1.35]; liver: RR_urban_ = 1.45 [95%CR: 1.30-1.62]). However, a heterogeneous effect of urbanicity was not evident among females. Prostate and breast cancer FBSIR tended to be lower in regions with greater deprivation, with no apparent heterogeneity due to urbanicity (prostate: RR for the entire region of Japan (RR_all_, without accounting for urbanicity) = 0.89 [95%CR: 0.86-0.92]; breast: RR_all_ = 0.91 [95%CR: 0.89-0.94]). However, the direction of the effect of ADI on FBSMR was opposite that of FBSIR in prostate cancer (RR_all_ = 1.08 [95%CR: 1.03-1.14]), and no clear association between ADI and FBSMR was observed in breast cancer. Cervical cancer FBSIR/FBSMR was higher in greater deprivation regions regardless of urbanicity, although FBSMR in suburban areas was not evident. For pancreatic cancer, there was no clear association with ADI and no apparent heterogeneity due to urbanicity.

**Figure 4.**
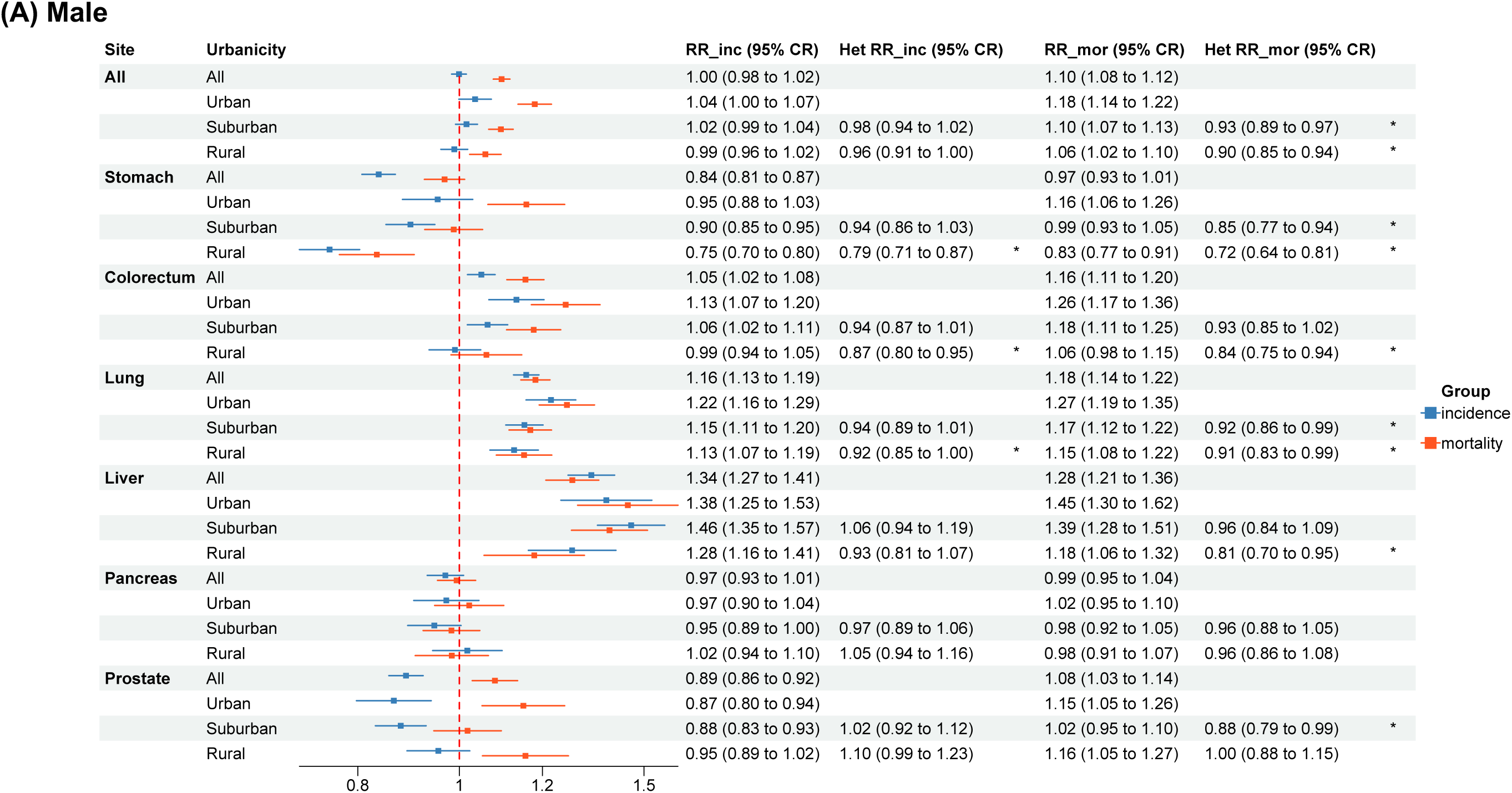
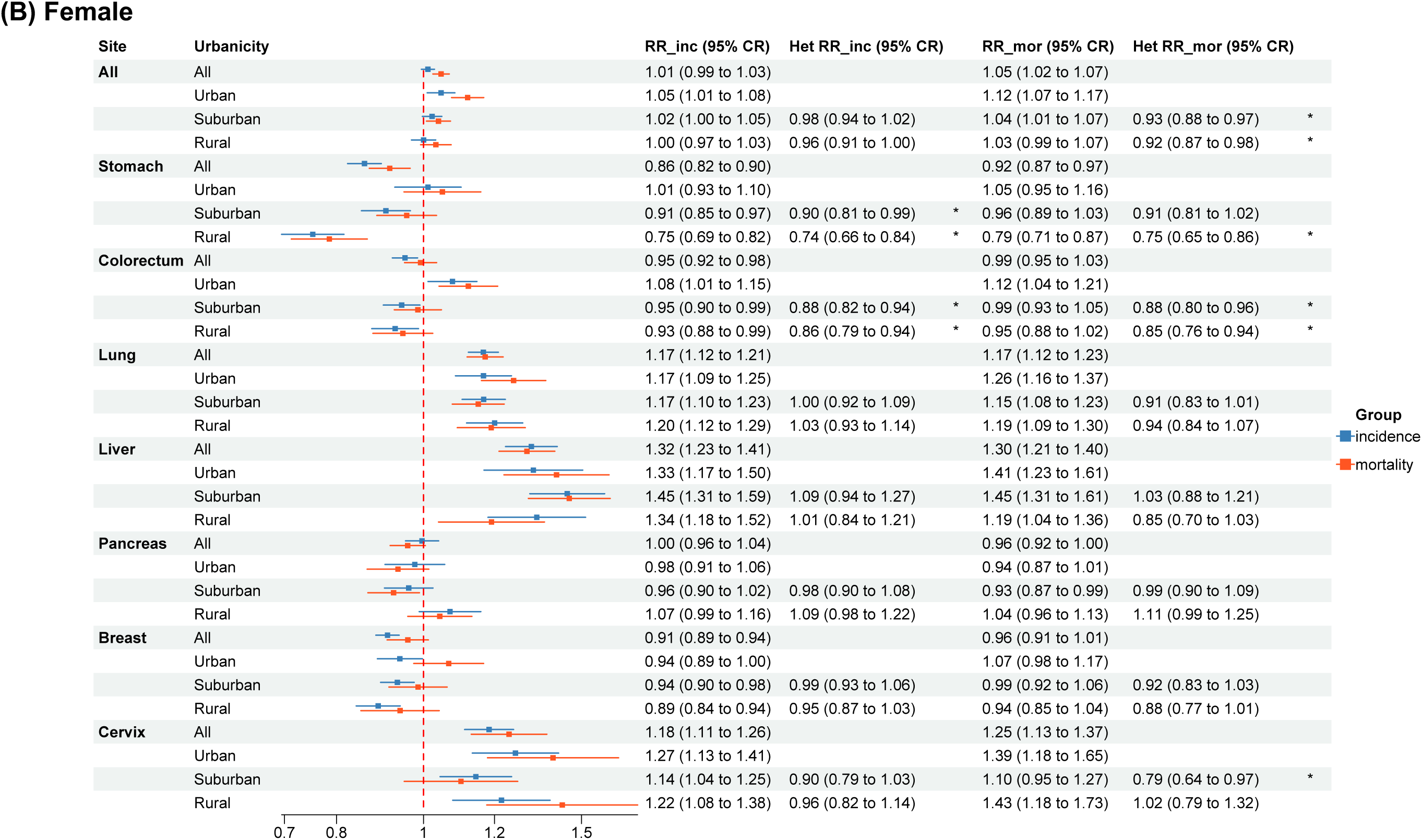
Impact of normADI on FBSIR/FBSMR by Sex and Urbanicity Forest plots illustrating the risk ratio (RR) and its 95% credible interval (CR) of FBSIR/FBSMR for the hypothetical worst ADI (normADI = 1) compared to the hypothetical best ADI (normADI = 0) for males (A) and females (B). Evidence of a difference was considered when the 95% CR of the RR did not include 1. Heterogeneity estimates marked with a single asterisk indicated notable variation. ADI, Areal Deprivation Index; FBSIR, fully Bayesian estimate of standardized incidence ratio; FBSMR, fully Bayesian estimate of standardized mortality ratio; Het, heterogeneity through levels of urbanicity (suburban vs. urban; rural vs. urban); normADI, normalized ADI; inc, incidence; mor, mortality.

Detailed results for the evaluation of model performance and the assessment of validity of incorporating spatial random effects, including the Watanabe-Akaike Information Criterion (WAIC)^26^, the probability integral transform (PIT)^27^, posterior predictive checks (PPC)^27^, and the Phi (𝜙) index^27^, are provided in Supplementary information, Supplementary Table 3, and Supplementary Figures 6 and 7. These assessments confirmed that the BYM2 model provided a well-calibrated fit, with WAIC indicating the best model performance, PIT histograms showing a generally uniform pattern, and PPC scatter plots demonstrating strong agreement between predictions and observations in most analyses. Furthermore, Phi values were generally high, with many cancer sites exceeding 0.8, suggesting that models that do not account for spatial effects may provide insufficient evaluations. Furthermore, we conducted a sensitivity analysis using ADI data from the 2010 Census, excluding 45 municipalities that could not be matched with cancer incidence or mortality data. No substantial differences were observed, as shown in Supplementary Figure 8.

## Discussion

To our knowledge, this study is the first attempt to evaluate the association between overall or site-specific cancer incidence/mortality and regional socioeconomic status by urbanicity using nationwide population-based data in an East Asian country, namely Japan. With 3,056,535 incident cases and 1,140,250 cancer deaths, this study showed that the impact of area-level socioeconomic status (here assessed as normADI) on cancer incidence/mortality may be heterogeneous among three categories of urbanicity, namely urban, suburban, and rural. The correspondence of higher levels of area deprivation to increased overall cancer mortality was more pronounced in urban areas than in suburban/rural areas for both sexes. In site-specific analyses, mortality of stomach, colorectal, lung, and liver cancers increased with increasing deprivation in urban areas, with the effect of deprivation being stronger for male colorectal, liver, and lung cancers as urbanicity increased. For incidence, the effects of normADI showed substantial variation by site and urbanicity, both in magnitude and direction: greater deprivation was associated with increased incidence for lung, liver, and cervical cancers, whereas inverse associations were noted for stomach, prostate, and breast cancers, with such association for stomach cancer being evident in rural and suburban areas and becoming stronger as urbanicity decreased. For colorectal cancer, incidence increased with deprivation in urban areas for both sexes, but an inverse association was observed in suburban and rural areas for females.

A striking finding of this study was the stronger positive association between normADI and mortality in urban areas than in suburban/rural areas for overall and some specific cancers. Among possible explanations, first, there is evidence that people with lower socioeconomic status living in urban areas are less likely to participate in cancer screening than those living in rural areas.^28–30^ Second, the differential magnitude of social capital between urban and rural areas might modify the impact of socioeconomic indicators on cancer outcomes. Social isolation is more common in urban than rural areas, and is associated with all-cause mortality.^31^ Our study may support this association in relation to cancer. In addition, social isolation is also more likely to occur in males than in females,^32^ which may explain why heterogeneity by urbanicity was more pronounced for males although the larger number of males in the study might have contributed to greater statistical power, potentially leading to more pronounced effects among males. Finally, a recent study showed that urban residents had contact with a less socioeconomically diverse range of individuals,^33^ which may also explain why regional disparities were more strongly reflected in urban areas. Whatever the reasons, demonstrating the potential of cancer prevention methods based on areal socioeconomic status and urbanicity will help determine the direction of cancer prevention measures at the local (or community) level; specifically, intensive interventions to aid accessibility to cancer screening and treatment for urban poor areas may be more effective in cancer mortality reduction.

We hypothesize that regional poverty impacts the risk behaviors of residents and their access to prevention and medical care, and thereby affects individual health outcomes: for lung cancer, a higher prevalence of smoking in impoverished areas; and for liver cancer, a higher prevalence of hepatitis B virus (HBV) and hepatitis C virus (HCV) infections in high poverty areas, coupled with fewer individuals receiving testing and treatment in these areas. In this regard, a previous study in Japan reported that a greater prevalence of HBsAg positivity (signifying active hepatitis B infection) and HCV infection was observed in municipalities with higher poverty levels and those with higher population density,^34^ supporting our findings that the association between normADI and liver cancer was strong, especially with higher urbanicity in male mortality. Our additional analyses, which accounted for non-viral liver cancer risk factors^35–37^ (‘*Effect of ADI on liver cancer considering non-viral risk factors*’ section in Supplementary information, Supplementary Tables 4 and 5, and Supplementary Figures 9 and 10), provided further validation for this hypothesis. These analyses also highlighted that in addition to viral etiologies, non-viral etiologies may play a significant role in the association between ADI and liver cancer among males, particularly in rural areas. Additionally, the association with cervical cancer incidence may imply a higher prevalence of human papillomavirus (HPV) infection in impoverished areas, as reported in the UK^38^. More importantly, studies in the US have shown lower vaccination rates in poorer regions,^39^ and the future spread of HPV vaccination in Japan may accentuate the heightened risk of incidence in impoverished areas. A comprehensive study which accounts for various risk factors to identify intervention points specific to each cancer site in impoverished areas now appears crucial from a preventative standpoint.

Among other findings, we found a higher incidence of breast and prostate cancers in wealthy regions, regardless of urbanicity. Several studies have shown a higher breast cancer incidence in wealthy areas,^40,41^ suggesting a higher proportion of women with increased risk of breast cancer (e.g. older age at first childbirth and higher attendance at mammography^42^) in these areas. The observed association in prostate cancer is also consistent with previous findings.^43^ Notably, however, this inverse association in incidence was attenuated in mortality for both cancers. In particular, for prostate cancer, the direction of the association was reversed, with greater deprivation associated with increased prostate cancer deaths. This may be partially explained by the limited availability of essential healthcare resources for individuals in socioeconomically disadvantaged areas.^44,45^ In fact, a large population-based analysis across 148 countries suggested that the significant recent decrease in breast cancer mortality in higher-income countries is due to the increasing proportion of cases diagnosed at an early stage and the availability of timely access to affordable and effective treatments.^46^ Furthermore, prostate cancer patients of lower socio-economic status were more likely to be diagnosed when their cancer was already at an advanced stage and had higher mortality than those of higher status, indicating the need for more education and screening for people of lower socioeconomic status.^43^ Therefore, while people living in wealthy regions remain at high risk of breast and prostate incidence and mortality, addressing secondary and tertiary prevention strategies in impoverished areas should also help alleviate the cancer mortality burden associated with both cancers.

Importantly, the observed relationships among regional socioeconomic status, urbanicity, and cancer incidence/mortality may differ across countries and populations. In contrast to our results, cancer mortality is higher in non-metropolitan than metropolitan areas in the US, and the impact of regional socioeconomic status is larger.^18^ A US-based study using SEER data further examined this association, reporting that the rural impact was greatest among those with localized disease and in high deprivation areas, though this study did not explicitly assess the interaction between urbanicity and ADI.^47^ This may partially reflect the lower uptake of cancer screening in US areas of greater deprivation and rurality^48^ versus the higher participation in rural than urban areas in Japan.^28,29^ Consistent with our findings, however, a retrospective cohort study of pediatric cancer using cancer registry data for Washington State in 1992-2013 showed that ADI-related survival disparities were greater in non-rural than rural areas.^21^ An Australian study on breast cancer survival also investigated the interaction between socioeconomic status (measured at the area level) and urbanicity.^49^ Their results indicated no significant interaction, a finding consistent with our study. Moreover, cancer incidence and mortality in Costa Rica, a middle-income country, were higher in urban areas than in rural areas and also higher among the wealthy than the poor.^19,20^ Thus, the impact of regional socioeconomic status and urbanicity on cancer risk varies by country and population. While studies evaluating the interaction between urbanicity and ADI in cancer epidemiology have been primarily conducted in Western countries, research from East Asia remains limited. Our study provides substantial evidence for understanding the reality of socioeconomic inequality in cancer risk in high-income countries of East Asia.

The strengths and limitations of this study should be noted. Strengths include 1) nationwide large-scale population-based data on cancer incidence and mortality, 2) validated local socioeconomic indicators based on the national census, and 3) analysis from a spatial viewpoint. As part of this spatial analysis, we chose fully Bayesian standardized incidence/mortality ratios (FBSIR/FBSMR) as the outcome indicator. SIR and SMR represent the ratio of observed cases to expected cases within a given region. Although these measures are conceptually straightforward, they tend to exhibit considerable variability, especially in areas with small populations or when analyzing rare diseases. This issue is particularly pronounced in high-dimensional spatial data, where limited case counts can result in unstable estimates. To address this issue, we employed a fully Bayesian approach using the BYM2 model, which stabilizes risk estimates by incorporating spatially structured and unstructured random effects. This allows for borrowing of information from neighboring areas, effectively reducing variability in small populations and improving the reliability of estimates. Furthermore, model comparison using WAIC demonstrated that the BYM2 model had the lowest WAIC, confirming its superior predictive performance over alternative models (Supplementary Table 3).

Additionally, the Phi index indicated the presence of spatial effects in the data (Supplementary Table 3), suggesting that models which neglect spatial dependencies may be subject to insufficient evaluation. These findings support the necessity of adopting a fully Bayesian spatial model in this study. In addition, while the mean FBSIR/FBSMR values in urban areas were above the national average and those in suburban and rural areas were at or below the national average, the differences in the mean FBSIR/FBSMR values by urbanicity were not substantial in many cancer sites (Supplementary Table 2). However, as illustrated in the exceedance probability maps (Figure 1 and Supplementary Figure 5), particularly in cancer site-specific evaluations, notable spatial variations in FBSIR/FBSMR values were observed at the municipal level. Furthermore, when incorporating ADI into the spatial analysis, clear differences by urbanicity became evident for overall cancer as well as for several specific cancer sites. This contrast suggests that while mean values may not strongly differentiate urban, suburban, and rural areas, deprivation plays a crucial role in shaping spatial disparities in cancer incidence and mortality.

Limitations include the lack of individual-level socioeconomic circumstances,^50^ albeit that analysis using area-level socioeconomic circumstances likely facilitates policies which take account of the combined effects of socioeconomic factors based on urbanicity. Second, area deprivation measured at a single point in time may not accurately reflect the deprivation experienced by cancer cases during an etiologically relevant period. The choice of a cross-sectional design and the use of ADI data from the 2015 Census were driven by the large-scale municipal consolidation in Japan which occurred primarily between 1999 and 2010. Since the national census is conducted every five years in Japan, use of ADI data from before this period would have resulted in municipalities that could not be completely matched with cancer incidence or mortality data, limiting the interpretability of our association analysis. Moreover, the results of a sensitivity analysis using ADI data from the 2010 Census showed no substantial differences, as shown in Supplementary Figure 8, further supporting the robustness of our interpretation. Third, this study lacks municipal-level data on environmental exposures such as air pollution and greenness, which may influence cancer incidence and mortality. Given evidence that pollution levels are strongly correlated with cancer incidence across various cancer sites,^51^ further research is needed to clarify the potential impact of environmental exposures on our findings.

In conclusion, the impact of areal socioeconomic status on cancer incidence/mortality is heterogeneous by cancer site and level of urbanicity. This study highlights the importance of considering urbanicity when examining the impact of ADI on cancer mortality; urban areas tend to show stronger associations between ADI and cancer mortality, suggesting that urban environments might exacerbate the negative effects of deprivation. Therefore, strategies for secondary and tertiary cancer prevention that focus on regional poverty are likely to become increasingly important, in light of the anticipated continued rise in urban populations. In addition, the varying impact of ADI on incidence for different cancer types underscores the need for tailored public health interventions.

Future research must adopt a multifaceted approach, incorporating international comparisons and multidisciplinarity to rectify cancer disparities, and requiring collaboration among urban planners, public health experts, and healthcare providers.

## Methods

### Geographical units

We conducted a cross-sectional study of geographic units at the municipal level in Japan. Geographic units are defined under the Local Autonomy Law as cities designated by ordinance, cities, towns, and villages. Among these, 20 cities designated by ordinance (Sapporo, Sendai, Saitama, Chiba, Yokohama, Kawasaki, Sagamihara, Niigata, Shizuoka, Hamamatsu, Nagoya, Kyoto, Osaka, Sakai, Kobe, Okayama, Hiroshima, Kitakyushu, Fukuoka, and Kumamoto) are further broken down into administrative wards, and Tokyo into special wards. As of 2015, a total of 1922 municipalities are recognized, consisting of 1724 municipalities, 23 special wards of Tokyo, and 175 wards of ordinance-designated cities. Of these, 1889 were included in the present analysis, excluding only 33 municipalities without population or within the evacuation zone established in Fukushima Prefecture following the Great East Japan Earthquake in 2011.

We then stratified these municipalities by urbanicity into three categories, namely, urban, suburban, and rural areas, by utilizing the criteria for establishing “central cities” and “surrounding areas” used in Census 2015^25^. In this study, we defined “central cities” as urban, “surrounding areas” as suburban, and areas other than these as rural areas (Supplementary Figure 1). The criteria are detailed in the English website of the 2018 Housing and Land Survey of Japan,^52^ which used the same criteria as Census 2015.

### Incidence and death data

Cancer was recorded based on the International Statistical Classification of Diseases and Related Health Problems (ICD-10), using C00-C96 (mortality: C00-C97) and invasive blood cancers (D45, D46, D460, D461, D462, D464, D465, D466, D467, D469, D471, D473, D474, and D475) as cancer incidence and mortality. In Japan National Cancer Registry, C97 is excluded from incidence data as it represents multiple primary malignancies rather than individual cancer occurrences. We also performed site-specific analyses on the eight most common cancers in Japan, namely stomach (C16), colorectum (C18-C20), lung (C33-C34), liver (C22), pancreas (C25), female breast (C50), prostate (C61), and cervix (C53). For incidence, we used data from the population-based National Cancer Registry from 2016 to 2018. The Japan National Cancer Registry is a nationwide, population-based system that collects and manages data on all cancer diagnoses across Japan.^53^ Introduced in 2016, the system ensures that cancer cases are systematically reported from all medical institutions, regardless of the patient’s residential area, through prefectural cancer registry offices. This centralized database enables comprehensive monitoring of cancer incidence and survival rates. Of the total 3,057,503 all cancer incident cases, 1,744,878 males and 1,311,657 females were included after excluding 213 case patients whose residence could not be determined, 657 case patients residing in seven municipalities in the evacuation zones established in Fukushima following the Great East Japan Earthquake in 2011, and 98 case patients with unknown sex or age. For mortality, we used mortality data from the Vital Statistics database for 2016 to 2018. The Vital Statistics of Japan are compiled by the Ministry of Health, Labor, and Welfare, based on legally mandated reports from local municipalities. These statistics comprehensively record births, deaths, marriages, divorces, and stillbirths reported under the Family Registration Act. We used cause-specific mortality by counting deaths where cancer was identified as the cause. Of the total 1,141,125 all cancer deaths, 670,991 males and 469,259 females were included in the analysis after excluding 210 case patients whose residence could not be determined and 665 case patients residing in the Fukushima evacuation zones (Supplementary Figure 2). For incidence we used residential location at the time of diagnosis, and for mortality we used residential location at the time of death.

### Population

Data on the sex-and 5-year age-specific population size of municipalities were available from the Census 2015; the census is conducted every five years in Japan.^25^

### Area-level socioeconomic status

Area-level socioeconomic status was evaluated using the ADI, a widely used composite indicator of geographical socioeconomic position by municipality,^11,17,23,24,54,55^ which is described in detail elsewhere.^17,23,24^ In this study, we estimated ADI as the weighted sum of eight census-based variables at the municipality level using data from the Census 2015 as follows:

ADI*_i_* = *k* × (2.99 × proportion of aged couple households*_i_* + 7.57 × proportion of aged single households*_i_* + 17.4 × proportion of single-mother households*_i_* + 2.22 × proportion of rented houses*_i_* + 4.03 × proportion of sales and service workers*_i_* + 6.05 × proportion of agricultural workers*_i_* + 5.38 × proportion of blue-collar workers*_i_* + 18.3 × unemployment rate*_i_*)

where *i* is the area index and *k* is a positive constant.^17^ A higher ADI value indicates greater socioeconomic deprivation, meaning that municipalities with higher ADI scores experience more severe socioeconomic disadvantages. The municipalities were sorted in ascending order according to ADI. We then calculated the normADI of a municipality as the cumulative proportion of population from the lower side ranging from 0 to 1 as follows:

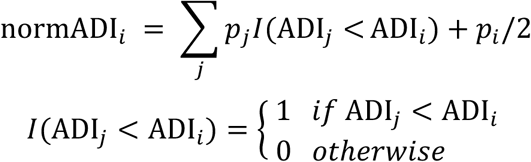

where *p_i_* is the ratio of the population of municipality *i* to the entire population in Japan, and normADI represents the relative position of a municipality among all municipalities in Japan ranging from zero (least deprived) to one (most deprived).^23^ This method provides a clear and direct way to interpret how variations in ADI correspond to changes in FBSIR/FBSMR. A key advantage of this approach is that it enhances the transparency and intuitiveness of the relationship between ADI and outcomes. Similar to the widely used Relative Index of Inequality (RII), which is typically estimated using Poisson regression analysis in epidemiological and public health research,^56–58^ it provides an indicator which shows how many times the SIR/SMR in the most deprived areas (normADI = 1) exceeds that in the least deprived areas (normADI = 0). Additionally, normADI standardizes values while explicitly accounting for population size, ensuring robustness in municipalities with limited data. Therefore, we utilized normADI as an indicator of area-level socioeconomic status throughout the study.

## Statistical analysis

### Descriptive analysis

We estimated municipality-level FBSIR (FBSMR) and their exceedance probabilities^59^, and visualized them using choropleth maps. To calculate FBSIR (FBSMR) for 2016–2018, we applied the indirect standardization method under the Poisson distribution:

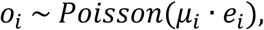

where 𝑜_i_ and 𝑒_i_ are the observed and expected numbers of incidences (mortalities) in municipality *i*, respectively, and 𝜇_*i*_ is the relative risk of incidence (mortality) in municipality *i*. The SIR (SMR) in each municipality is obtained as 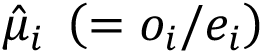 the maximum likelihood estimate quantity of 𝜇_i_. The expected numbers were derived based on the age-specific incidence (mortality) rates of the reference population (Japan’s total population) and the corresponding age distribution of the observed population. Age groups were categorized into five-year strata, with the highest group being 85+, ranging from 0–4 to 85+. To model incidence and mortality counts and estimate FBSIR (FBSMR), we then applied the Besag-York-Mollié 2 (BYM2) model^22^ using the Integrated Nested Laplace Approximation (INLA) framework^60^:

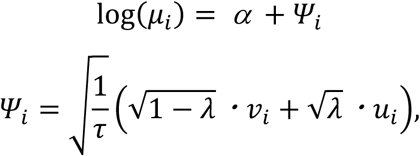

where 𝜏 is the reciprocal of the overall variance, λ is the mixing parameter controlling the balance between spatially structured and unstructured effects, and 𝑣_!_ and 𝑢_!_ represent the spatially unstructured random effect and the scaled spatially structured random effect for municipality *i*, respectively. The scaled spatially structured component 𝑢_!_ in the BYM2 model is derived from an intrinsic Conditional AutoRegressive prior, which enforces spatial smoothing by introducing dependencies between neighboring municipalities. This dependency structure is typically defined using an adjacency matrix, which represents the spatial relationships between municipalities. In this study, we constructed an adjacency matrix using the poly2nb() function in the R spdep package, which determines neighboring municipalities based on shared borders. The priors used in the analysis correspond to the default parameter values specified in the INLA framework. The FBSIR (FBSMR) for each municipality was obtained by estimating the mean of the posterior distribution of the relative risk (𝜇_!_). Exceedance probabilities^59^ were determined based on a threshold of a 10% increased risk (FBSIR [FBSMR] = 1.1).

### Association analysis

To identify the association between normADI and FBSIR (FBSMR), we included a linear prediction term with explanatory variables in the BYM2 model:

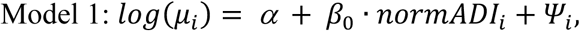

where 𝛼 is intercept and 𝛽_0_ is coefficient of normADI.

We then evaluated potential heterogeneity in the impact of normADI within different levels of urbanicity by including interaction terms. Specifically, urbanicity was modeled as a categorical variable with three levels (urban = 1, suburban = 2, and rural = 3), where suburban and rural areas were represented using dummy variables and with urban areas serving as the reference category. The model was specified as follows:

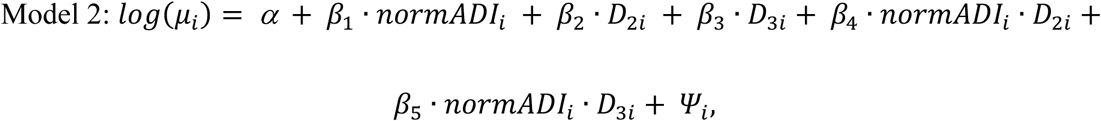

where 𝐷_2𝑖_ and 𝐷_3𝑖_ are dummy variables indicating suburban (level2) and rural (level3) areas, respectively:

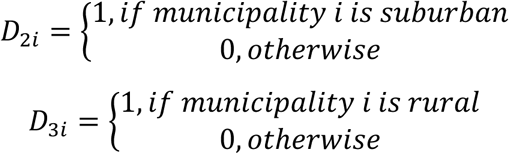

In this formulation, 𝛼 represents the intercept, and coefficient 𝛽_$_is the overall effect of normADI in urban areas. The coefficients 𝛽_%_ and 𝛽_&_ capture the main effects of suburban and rural areas, respectively, relative to urban areas. The interaction terms 𝛽_’_ and 𝛽_(_ assess the differential impact of normADI in suburban and rural areas compared to urban areas.

### Presentation of model results

The risk ratio (RR) of FBSIR (FBSMR) for the hypothetical worst ADI (normADI = 1) compared to the hypothetical best ADI (normADI = 0) in the entire region of Japan is defined using the coefficients in Model 1 as follows:

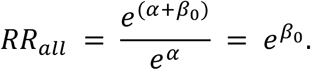

Accordingly, the RR for normADI by urbanicity can be considered using the coefficients in Model 2 as follows:

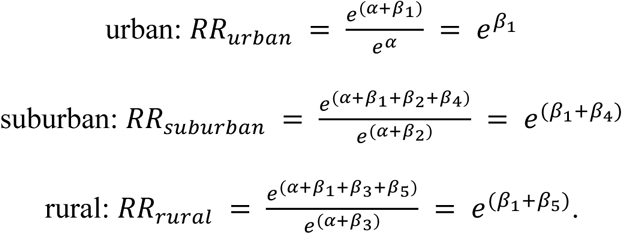

The heterogeneity of suburban and rural areas compared to urban areas was evaluated using 𝛽_4_ and 𝛽_5_, respectively, in Model 2 as follows:

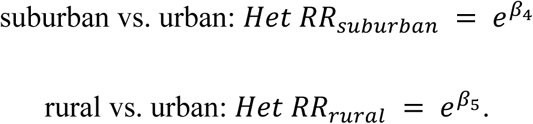

### Model evaluation and validation

To assess model performance and validation of the use of spatial random effects, we employed several evaluation metrics^61^, including the WAIC^26^, the PIT^62^, PPC^62^, and the Phi (𝜙) index^27^.

Details on the evaluation methods and interpretation of these indicators are provided in the Supplementary information.

All statistical analyses were performed using R (version 4.2.1 or 4.3.2). For model estimation, we used the R package ‘R-INLA’^63^ (version 24.02.09). The R code used to call INLA is provided in the Supplementary information. Evidence of a difference was considered when the 95% CR of the RR did not include 1.

## Contributors

YNK, KM, YK, RY and HI designed the study concept. MK, YK, and HI obtained and managed the data. YNK, MK, KM, YK, TO, KT, RY, TN, and HI designed the analytical methods. All authors provided input into interpretation of results. YNK, MK, KM, YK, TN, and HI wrote the first draft of the paper; all authors contributed to the revision and finalization of the paper. YNK, MK, YK, YT, TO, KT, RY, and HI had full access to all data used in this study. Due to data permission restrictions, not all authors were able to access the underlying data used in the study.

## Ethics statements

Approval of the research protocol by an institutional review board: This study was approved by the institutional review board of the Aichi Cancer Center (approval number: 2021-1-0006). We obtained the anonymized dataset from the Japanese NCR under the Cancer Registry Promotion Act and independently processed it. Mortality data were obtained from the Vital Statistics of Japan provided under the Statistics Act (Act No. 53 of 2007) and were independently processed. Additionally, data on lifestyle habits were obtained from the Lifestyle Health Check-ups and Health Guidance Program under the Act on Assurance of Medical Care for Elderly People and were also independently processed. Due to privacy and ethical restrictions, these datasets are not publicly accessible and are available only upon request.

## Data availability

Provision of the Japan National Cancer Registry data, the Vital Statistics data, and the Lifestyle Health Check-ups and Health Guidance Program data to third parties is prohibited. Census data (population, urbanicity classifications, indicators of ADI calculation) are available at https://www.e-stat.go.jp/en/ (Accessed 3 Mar 2025).

## Acknowledgements

This study was supported by JSPS KAKENHI (Grants-in-Aid for Scientific Research) B Grant Number JP21H03210, the Aichi Cancer Center Joint Research Project on Priority Areas (2018-07 and 2021-06) and the National Cancer Center Research and Development Fund (2021-A-20).

We also thank Guy Harris DO of Dmed (http://dmed.co.jp) for editing drafts of this manuscript.

---

**Supplementary Figure 1. Map of Urbanicity by Municipality in Japan**

Municipalities were geographically divided into three groups of urbanicity by utilizing the criteria for establishing “central cities” and “surrounding areas” used in the Census data of the year 2015. We defined “central cities” as urban (navy), “surrounding areas” as suburban (green), and areas other than “central cities” and “surrounding areas” as rural (gray).

**Supplementary Figure 2. Analysis Subjects**

**Supplementary Figure 3. Distribution of Individual Cancer Types Across the Three Urbanicity Categories for Incidence (A) and Mortality (B)**

**Supplementary Figure 4. Map of the Islands of Japan and the Three Major Metropolitan Cities**

The islands comprising Japan are indicated by color coding. The three major metropolitan cities are indicated by dots on the map.

**Supplementary Figure 5. Geographic Distribution of FBSIR/FBSMR and Their Exceedance Probabilities by Sex and Municipality for Each Cancer**

Color-coded maps of FBSIR and FBSMR (ranging from red [high] to blue [low]) and their exceedance probabilities of all cancers are shown for males (A) and females (B). The exceedance probabilities represent the probabilities of FBSIR/FBSMR values being greater than 1.1 within individual municipalities. Major central cities and their surrounding areas are broken out. As this study follows census-based regional definitions for consistency, some discrepancies between municipality names and their corresponding prefectures arise (e.g.,’Hiroshima’ refers to the Hiroshima metropolitan area rather than Hiroshima Prefecture). FBSIR, fully Bayesian estimate of standardized incidence ratio; FBSMR, fully Bayesian estimate of standardized mortality ratio. (A) Stomach; (B) Colorectum; (C) Lung; (D) Liver; (E) Pancreas; (F) Breast; (G) Prostate; (H) Cervix.

**Supplementary Figure 6. Histograms of the Probability Integral Transform (PIT) per Municipality by Cancer Site and Sex for Incidence and Mortality**

**Supplementary Figure 7. Posterior Predictive Check (PPC): Scatterplots of Posterior Predictions vs. Observed Counts and Histograms of Posterior Predictive P-Values per Municipality by Cancer Site and Sex for Incidence and Mortality**

**Supplementary Figure 8. Impact of normADI (based on 2010 Census) on FBSIR/FBSMR by Sex and Urbanicity**

Forest plots illustrating the risk ratio (RR) and its 95% credible interval (CR) of FBSIR/FBSMR for the hypothetical worst ADI (normADI = 1) compared to the hypothetical best ADI (normADI = 0) for males (A) and females (B). Evidence of a difference was considered when the 95% CR of the RR did not include 1. Heterogeneity estimates marked with a single asterisk indicated notable variation. ADI, Areal Deprivation Index; FBSIR, fully Bayesian estimate of standardized incidence ratio; FBSMR, fully Bayesian estimate of standardized mortality ratio; Het, heterogeneity through levels of urbanicity (suburban vs. urban; rural vs. urban); normADI, normalized ADI; inc, incidence; mor, mortality.

**Supplementary Figure 9. Distribution of the Prevalence of BMI≥25, Diabetes, Dyslipidemia, Insufficient Exercise, Smoking, and Drinking Across the Three Urbanicity Categories by Sex**

**Supplementary Figure 10. Conceptual Directed Acyclic Graph (DAG) Outlining the Assumed Causal Relationships in Our Adjusted Models**

The adjusted risk ratio estimates the direct effect* of ADI on liver cancer FBSIR/FBSMR, reflecting the impact of ADI through pathways independent of mediators (i.e., obesity, diabetes, dyslipidemia, insufficient exercise, smoking, and drinking). This adjustment was confirmed to be correct in DAGitty (https://www.dagitty.net/). Certain paths that do not directly link the exposure and outcome are represented with dashed lines for visual clarity. ADI, Areal Deprivation Index; FBSIR, fully Bayesian estimate of standardized incidence ratio; FBSMR, fully Bayesian estimate of standardized mortality ratio.

