## Supplementary information for "The Urban Divide: Socioeconomic Status and Its Impact on Cancer in the Age of Urbanization"

**Supplementary Tables** (Electronic Excel file)

Supplementary Table 1. Mean (SD) and Median (IQR) Values of SIR and SMR in Japan by Urbanicity (2016–2018)

Supplementary Table 2. Mean (SD) and Median (IQR) Values of FBSIR and FBSMR in Japan by Urbanicity (2016–2018)

Supplementary Table 3. Comparison of Bayesian Models by Cancer Site and Sex: A Summary of WAIC, DIC, RRs, and Phi (for BYM2 Model Only) for Incidence and Mortality

Supplementary Table 4. Prevalence of Individuals Meeting Each Criterion for Liver Cancer Risk Factors in Data from the Lifestyle Health Check-ups and Health Guidance Program (2016–2018), Stratified by Urbanicity

Supplementary Table 5. Comparison of Models With and Without Liver Cancer-Related Risk Factors: Impact of normADI on Liver Cancer FBSIR/FBSMR by Sex and Urbanicity

### *Supplementary Methods and Results*

#### *Model Evaluation and Validation*

To assess model performance and validation of the use of spatial random effects, we employed several evaluation metrics[1], including the Watanabe-Akaike Information Criterion (WAIC)[2], the probability integral transform (PIT)[3], posterior predictive checks (PPC)[3], and the Phi ($\phi$) index[4]. The WAIC was used to assess out-of-sample predictive accuracy, serving as a fully Bayesian alternative to traditional information criteria. Lower WAIC values indicate a better performance in terms of trade-off between model complexity and goodness of fit. To validate selection of the Besag-York-Mollié 2 (BYM2) model[5] as the primary model, we compared the WAIC values of five candidate models (BYM2 model, BYM model, Poisson-Gamma model, Generalized linear mixed model [GLMM], and Generalized linear model [GLM]). PIT is an indicator that assesses the goodness of fit between the predictive distribution of a model and the observed data. PIT values range from 0 to 1 and, under a well-calibrated model, are expected to follow a uniform distribution. We computed PIT values and visualized them using a histogram. PPC were conducted to assess the model's ability to reproduce observed data. We compared simulated datasets generated from the posterior predictive distribution with actual observations, assessing discrepancies using both graphical and numerical summaries. These included a scatter plot of the posterior mean for predictive distributions versus observed values and a histogram of posterior predictive p-values (PPP). Ideally, the scatter plot displays points distributed around the diagonal (y = x) line, indicating good agreement between the predictions and observations. The histogram of PPP exhibits a reasonable spread of values toward the middle of the range, without excessive clustering near 0 or 1, which would indicate model misfit. The Phi ($\phi$) index is a measure of the contribution of spatial random effects in Bayesian spatial models. It quantifies the proportion of total variance attributed to the spatially structured random effect relative to the total variability in the model. In the context of spatial hierarchical models, including those implemented using INLA, Phi ($\phi$) is defined as:

$$\phi = \frac{\sigma^2_{\text{spatial}}}{\sigma^2_{\text{spatial}} + \sigma^2_{\text{unstructured}}}$$

where $\sigma^2_{\text{spatial}}$ represents the variance of the spatially structured random effect, and $\sigma^2_{\text{unstructured}}$ represents the variance of the unstructured random effect. The value of $\phi$ ranges between 0 and 1, where $\phi \approx 1$ represents that the model is dominated by spatial random effects, meaning spatially structured variation explains most of the variance; $\phi \approx 0$ represents that the model is dominated by unstructured random effects, meaning that local noise is more influential than spatial dependence.

Among the five models tested (GLM, GLMM, Poisson-Gamma model, BYM model, and BYM2 model), the BYM2 model had the lowest WAIC, indicating the best model performance (**Supplementary Table 3**). For the BYM2 model, the PIT histogram, as shown in **Supplementary Figure 6**, exhibited a generally uniform pattern, although some analyses showed slight deviations in the tails. This suggested that the model provided a reasonable overall fit to the data, while minor discrepancies at the extremes may indicate potential limitations in capturing extreme values. Furthermore, the PPC scatter plot for the BYM2 model in **Supplementary Figure 7** demonstrated that data points were generally distributed around the diagonal (y = x) line in most analyses, indicating good agreement between the predictions and observations. Similarly, the histogram of PPP with the BYM2 model showed a reasonable spread of values toward the middle of the range, with no excessive clustering near 0 or 1 in most analyses. These results suggest that the model provides a well-calibrated fit to the data and appropriately captures the underlying patterns. Finally, as shown in **Supplementary Table 3**, the Phi values were generally high, with many cancer sites exceeding 0.8. This result suggests the presence of spatial effects in the data, indicating that evaluations using models that do not account for spatial effects may be insufficient. Based on the above results, this study mainly focused on the results of the BYM2 model, while the results from the other models are provided in **Supplementary Table 3**.

#### *Effect of ADI on liver cancer considering non-viral risk factors*

We observed strong associations between normADI and liver cancer. Besides viral etiologies, non-viral factors such as metabolic dysfunction-associated steatotic liver disease (MASLD), obesity, and diabetes have been rising in prevalence across many countries.[6,7] To assess how liver cancer risk factors may have influenced the observed outcomes in this study, we incorporated municipal-level data on the prevalence (%) of a body mass index (BMI) ≥25, diabetes, dyslipidemia, and insufficient exercise. Furthermore, we also included information on smoking and alcohol consumption, which are both well-established risk factors for liver cancer[8]. We applied for access to data from the Lifestyle Health Check-ups (Tokutei Kenshin) and Health Guidance Program provided by Japan's Ministry of Health, Labour and Welfare and obtained municipal-level data on the prevalence of non-viral liver cancer risk factors. Participants in the Lifestyle Health Check-ups were individuals aged 40 to 74 years who underwent the examination between 2016 and 2018. In this study, the prevalence of certain conditions was determined by calculating the proportion of participants who met the following criteria based on the health checkup assessment items. Obesity was defined by BMI of ≥25 at the time of the health checkup; diabetes was defined as a fasting blood glucose level of ≥126 mg/dL, HbA1c level of ≥6.5%, or a response of "yes" to a question on the use of "glucose-lowering medications or insulin injections"; dyslipidemia was defined as triglyceride levels of ≥150 mg/dL, HDL cholesterol levels of <40 mg/dL, LDL cholesterol levels of ≥140 mg/dL, or a response of "yes" to a question on the use of "cholesterol- or triglyceride-lowering medications". Insufficient exercise was defined as a response of "no" to all three of the following questionnaire items regarding physical activity: (1) engaging in exercise for ≥30 minutes at least twice a week for ≥1 year, (2) engaging in walking or equivalent activity for ≥1 hour per day, and (3) having a walking speed faster than peers of the same age and sex. Smoking was defined as a response of "yes" to a question on habitual tobacco use; drinking was defined as a response to the questionnaire indicated a daily alcohol intake of ≥2 *gou* (equivalent to approximately 360 mL of sake or 40 g of ethanol).

As shown in **Supplementary Table 4** and **Supplementary Figure 9**, the distribution of these factors (i.e., prevalence of BMI≥25, diabetes, dyslipidemia, insufficient exercise, smoking, and drinking) remained largely consistent across the three urbanicity categories, suggesting no substantial urban–rural patterning among these factors.

To investigate how these factors might explain the observed liver cancer outcomes, we accounted for them as covariates, assuming the causal structure outlined in **Supplementary Figure 10**. Under this causal framework, the adjusted RRs estimate the direct effects of ADI on liver cancer FBSIR (FBSMR), reflecting the impact of ADI through pathways independent of mediators (i.e., obesity, diabetes, dyslipidemia, insufficient exercise, smoking, and drinking).[9] This adjustment was confirmed to be correct in DAGitty.[10]

The overall pattern of association remained largely unchanged after adjustment (**Supplementary Table 5**). However, while we observed consistent point estimates between the unadjusted and adjusted models in females, the point estimates in males declined following adjustment, and the previously evident association between ADI and liver cancer FBSIR/FBSMR in rural areas was no longer observed. These additional analyses demonstrated that the impact of ADI on liver cancer FBSIR/FBSMR was evident in males in urban and suburban areas, as well as in females, even after adjustment for non-viral risk factors such as obesity, diabetes, dyslipidemia, insufficient exercise, smoking, and drinking. This finding suggests that key factors influencing liver cancer incidence and mortality, such as hepatitis B virus (HBV) and hepatitis C virus (HCV) infections—both of which are more prevalent in high-poverty areas[11] and are often accompanied by lower rates of testing and treatment—may contribute to the observed associations, though other unmeasured factors may also be involved. Furthermore, these analyses highlighted that non-viral etiologies also play a significant role in the association between ADI and liver cancer among males, particularly in rural areas. However, as this study is an ecological study, assessing the validity of the observed associations necessitates well-designed prospective studies.

### *R code used to call INLA*

```
### Data Structure (1): Description of the "DATAset" contents
# About [DATAset]
# DATAset$cityID : ID of each municipality in Japan
# DATAset$UR : Urbanicity in each municipality in Japan
# DATAset$ADI : normalized Areal Deprivation Index in each municipality in Japan
# DATAset$Observed :  Observed number of Incidences/deaths of Certain Cancer Sites
#                     among Males/Females in each municipality in Japan
# DATAset$Expected :  Expected number of Incidences/deaths of Certain Cancer Sites
#                     among Males/Females in each municipality in Japan

### Data Structure (2): Description of "JPgeometry" (shapefile, polygon data) contents
# About [JPgeometry]
# JPgeometry$cityID : ID of each municipality in Japan
# JPgeometry$geometry : Geographic Polygon Data in each municipality in Japan

### Creation of Neighbor Matrix: Used for BYM/BYM2 models
library(spdep)
library(INLA)
Shape  <- merge(JPgeometry, DATAset, by="cityID")
nbcity <- poly2nb(Shape)
nb2INLA("nblist.adj", nbcity)

### Loading the Neighbor Matrix: Specifying gcity for use in INLA's BYM/BYM2 models
gcity <- inla.read.graph("nblist.adj")

### Prior Settings: Using the default values
prior <- list(prec = list(prior = "pc.prec", param = c(1 , 0.01)), phi = list(prior = "pc", param = c(0.5, 0.5)))

######## Model Setup and INLA Execution ###########

library(INLA)

### Model 0: Model for Calculating Values for Choropleth Maps ###
#BYM2
formula0 <- Observed ~ f(cityID, model = "bym2", graph = gcity)
results0 <- inla(formula0, family = "poisson", data = DATAset, E = Expected,
   control.compute = list(dic = TRUE, waic = TRUE), control.predictor = list(compute = TRUE))

### Model 1: Model for Estimating β Without Urban Classification ###
#BYM2
formula1 <- Observed ~ ADI + f(cityID, model = "bym2", graph = gcity )
results1 <- inla(formula1, family = "poisson", data=DATAset, E = Expected,
   control.compute = list(dic = TRUE, waic = TRUE), control.predictor = list(compute = TRUE))

### Model 2: Model for Estimating β for Each Urban Classification ###
#BYM2
formula2  <- Observed ~ ADI + factor(UR) + ADI*factor(UR) + f(cityID, model = "bym2", graph = gcity )
results2  <- inla(formula2, family = "poisson", data = DATAset, E = Expected,
   control.compute = list(dic = TRUE, waic = TRUE), control.predictor = list(compute = TRUE))
```

```
### Comparison Model for Model 1 ###
#BYM
formula1_bym  <- Observed ~ ADI + f(cityID, model = "bym", graph = gcity )
results1_bym  <- inla(formula1_bym, family = "poisson", data = DATAset, E = Expected,
   control.compute = list(dic = TRUE, waic = TRUE), control.predictor = list(compute = TRUE))

#poisson-gamma
formula1_poga <- Observed ~ ADI + f(cityID, model = "iid")
resluts1_poga <- inla(formula1_poga, family = "nbinomial", data = DATAset, E = Expected,
   control.compute = list(dic = TRUE, waic = TRUE), control.predictor = list(compute = TRUE))

#GLMM
formula1_GLMM <- Observed ~ ADI + f(cityID, model = "iid") + offset(log(Expected))
results1_GLMM <- inla(formula1_GLMM, family = "poisson", data = DATAset,
   control.compute = list(dic = TRUE, waic = TRUE), control.predictor = list(compute = TRUE))

#GLM
formula1_GLM  <- Observed ~ ADI + offset(log(Expected))
results1_GLM  <- inla(formula1_GLM, family = "poisson", data = DATAset,
   control.compute = list(dic = TRUE, waic = TRUE), control.predictor = list(compute = TRUE))

### Comparison Model for Model 2 ###
#BYM
formula2_bym  <- Observed ~ ADI + factor(UR) + ADI*factor(UR) + f(cityID, model = "bym", graph =
gcity )
results2_bym  <- inla(formula2_bym, family = "poisson", data = DATAset, E = Expected,
   control.compute = list(dic = TRUE, waic = TRUE), control.predictor = list(compute = TRUE))

#poisson-gamma
formula2_poga <- Observed ~ ADI + factor(UR) + ADI*factor(UR) + f(cityID, model = "iid")
results2_poga <- inla(formula2_poga, family = "nbinomial", data = DATAset, E = Expected,
   control.compute = list(dic = TRUE, waic = TRUE), control.predictor = list(compute = TRUE))

#GLMM
formula2_GLMM <- Observed ~ ADI + factor(UR) + ADI*factor(UR) + f(cityID, model = "iid") +
offset(log(Expected))
results2_GLMM <- inla(formula2_GLMM, family = "poisson", data = DATAset,
   control.compute = list(dic = TRUE, waic = TRUE), control.predictor = list(compute = TRUE))

#GLM
formula2_GLM  <- Observed ~ ADI + factor(UR) + ADI*factor(UR) + offset(log(Expected))
results2_GLM  <- inla(formula2_GLM, family = "poisson",data = DATAset,
   control.compute = list(dic = TRUE, waic = TRUE), control.predictor = list(compute = TRUE))
```

```
### Predictive Distribution ###

# Execute INLA (with cpo=TRUE to generate PIT)
formula2  <- Observed ~ ADI + factor(UR) + ADI*factor(UR) + f(cityID, model = "bym2", graph = gcity )
results2  <- inla(formula2, family = "poisson", data = DATAset, E = Expected,
    control.compute = list(dic = TRUE, waic = TRUE, cpo = TRUE, return.marginals.predictor = TRUE),
    control.predictor = list(compute = TRUE))

# Extracting pit values
pit_values <- results2$cpo$pit

# Calculate retrieve predicted risk ratios
predicted_RRs <- as.numeric(results2$summary.fitted.values[, "mean", drop = FALSE]$mean)

# Calculate predicted counts (risk ratio * expected value)
predicted_counts <- predicted_RRs * DATAset$Expected

# Observed values from DATAset
observed_counts <- DATAset$Observed

# Simulate and generate posterior predictions
simulated_data <- replicate(5000, rpois(n = length(observed_counts), lambda = predicted_counts))

# Calculate the posterior predictive mean for each observation
ppd_means <- rowMeans(simulated_data)

# Calculate the posterior predictive mean per municipality
ppp_per_city <- rowMeans(simulated_data <= observed_counts)

# Create dataframe
pd_data <- data.frame(CityID = DATAset$cityID,
   PIT= pit_values, Observed = observed_counts, PredictedMean = ppd_means, PPP= ppp_per_city)

## plot ##

## <Histogram of Probability Integral Transform (PIT)> ##
hist(pd_data$PIT, breaks = 20, col = "blue", main = " Histogram of PIT",
   xlab = "Probability", ylab = "Count of Municipalities")

## <Posterior Predictive Check> (Observed vs. Posterior Predictive Mean) ##
ggplot(pd_data, aes(x = Observed, y = PredictedMean)) +
geom_point(alpha = 0.5) +
geom_abline(slope = 1, intercept = 0, color = "red", linetype = "dashed") +
labs(title = "Posterior Predictive Check",
   x = "Observed Counts", y = "Predicted Mean (Posterior Predictive Distribution)") +
theme_minimal()

## <Histogram of Posterior Predictive p-values> ##
hist(pd_data$PPP, breaks = 10, col = "lightblue", main = "Histogram of Posterior Predictive p-values",
   xlab = "PPP (Posterior Predictive p-value)", ylab = "Count of Municipalities")
```

**Supplementary Figure 1. Map of Urbanicity by Municipality in Japan**

Municipalities were geographically divided into three groups of urbanicity by utilizing the criteria for establishing "central cities" and "surrounding areas" used in the Census data of the year 2015. We defined "central cities" as urban (navy), "surrounding areas" as suburban (green), and areas other than "central cities" and "surrounding areas" as rural (gray).

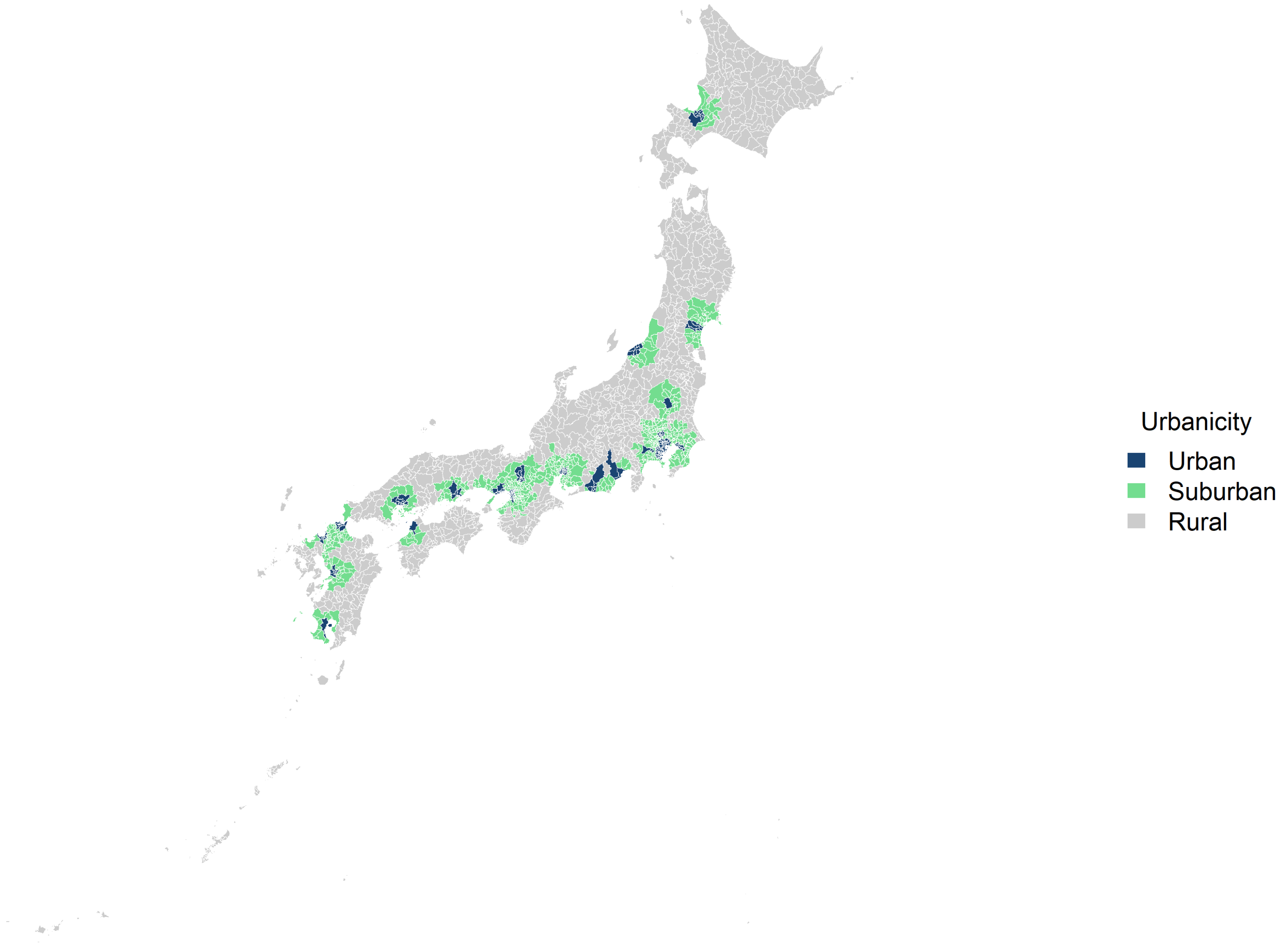

**Supplementary Figure 2. Analysis Subjects**

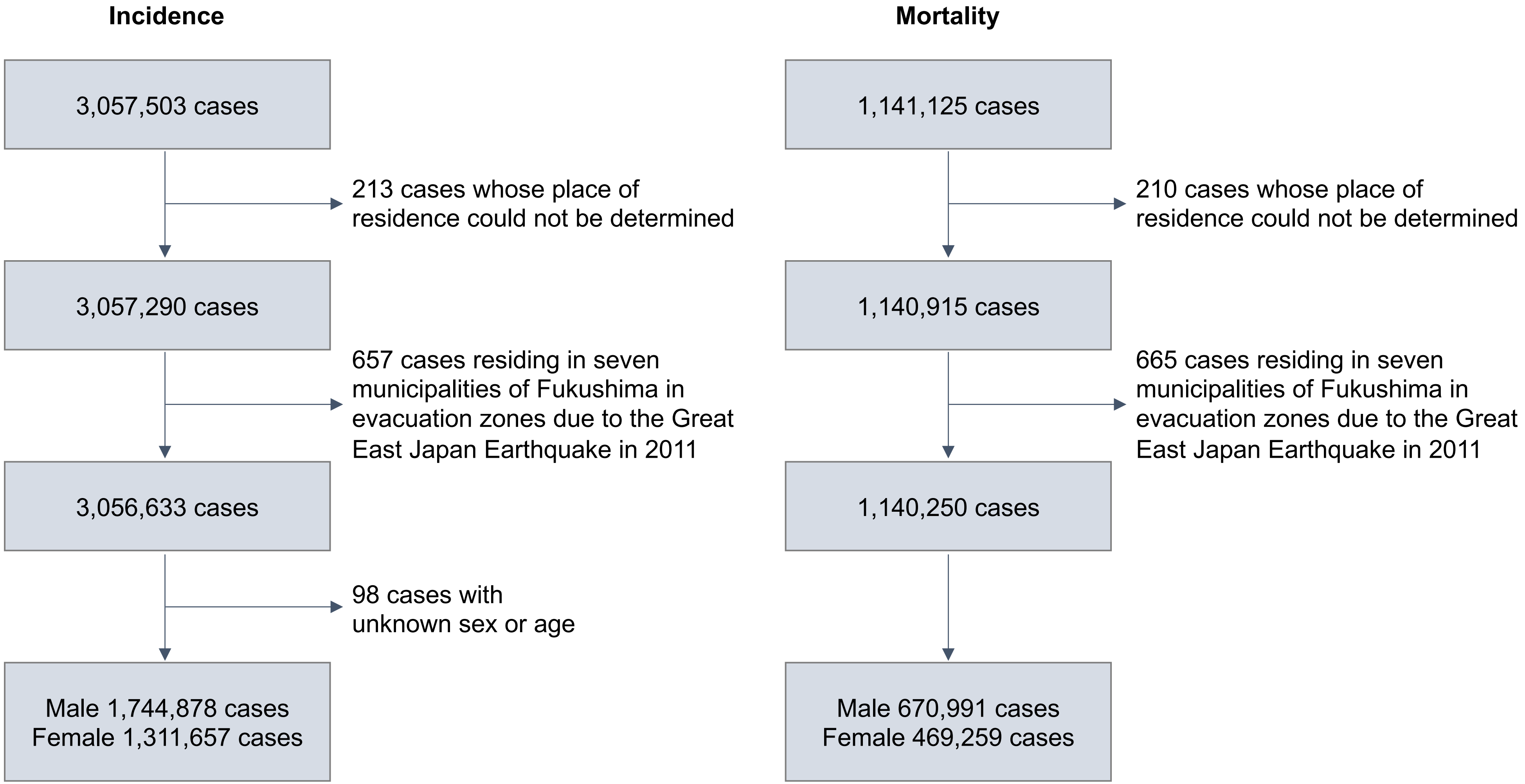

**Supplementary Figure 3. Distribution of Individual Cancer Types Across the Three Urbanicity Categories for Incidence (A) and Mortality (B)**

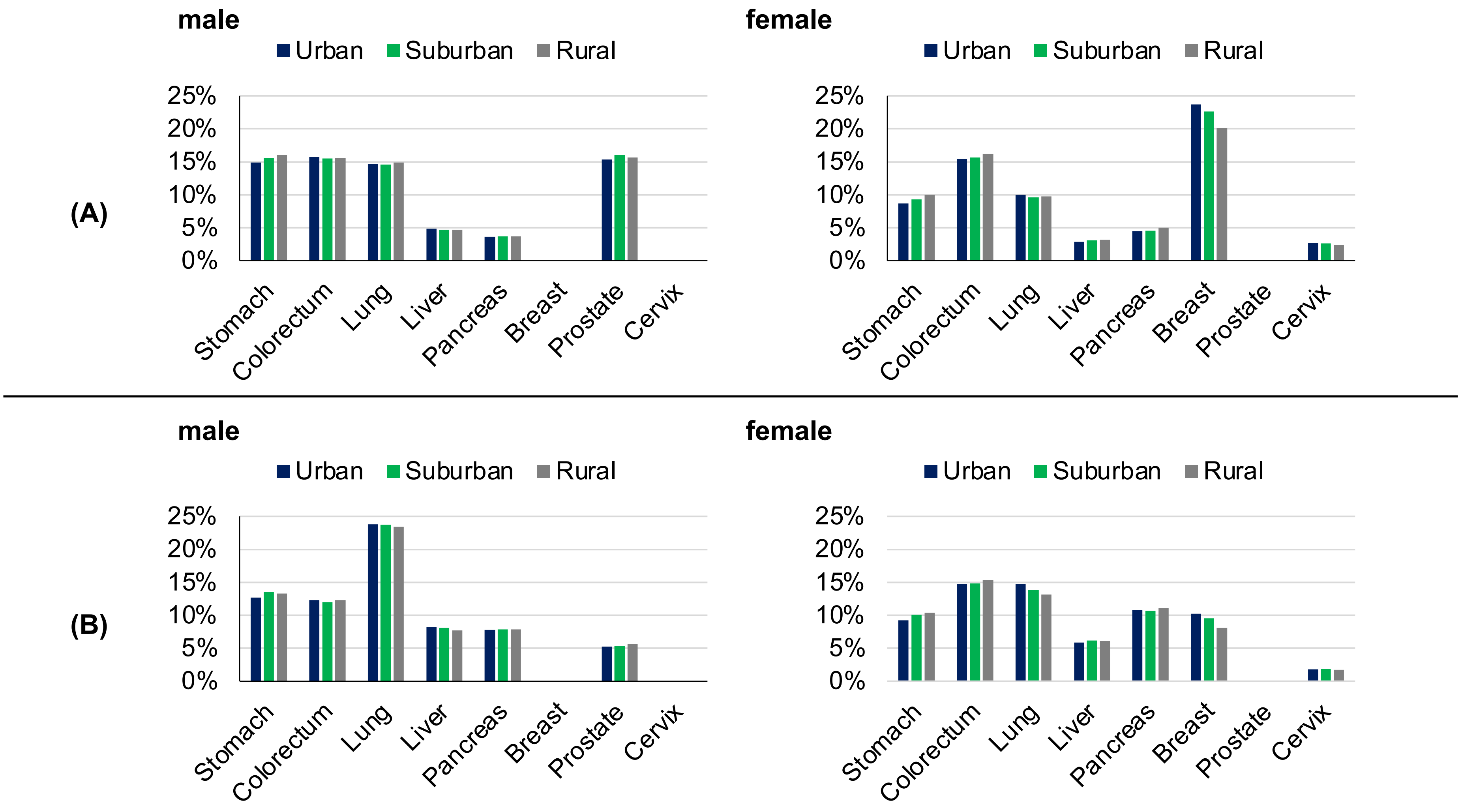

**Supplementary Figure 4. Map of the Islands of Japan and the Three Major Metropolitan Cities**
The islands comprising Japan are indicated by color coding. The three major metropolitan cities are indicated by dots on the map.

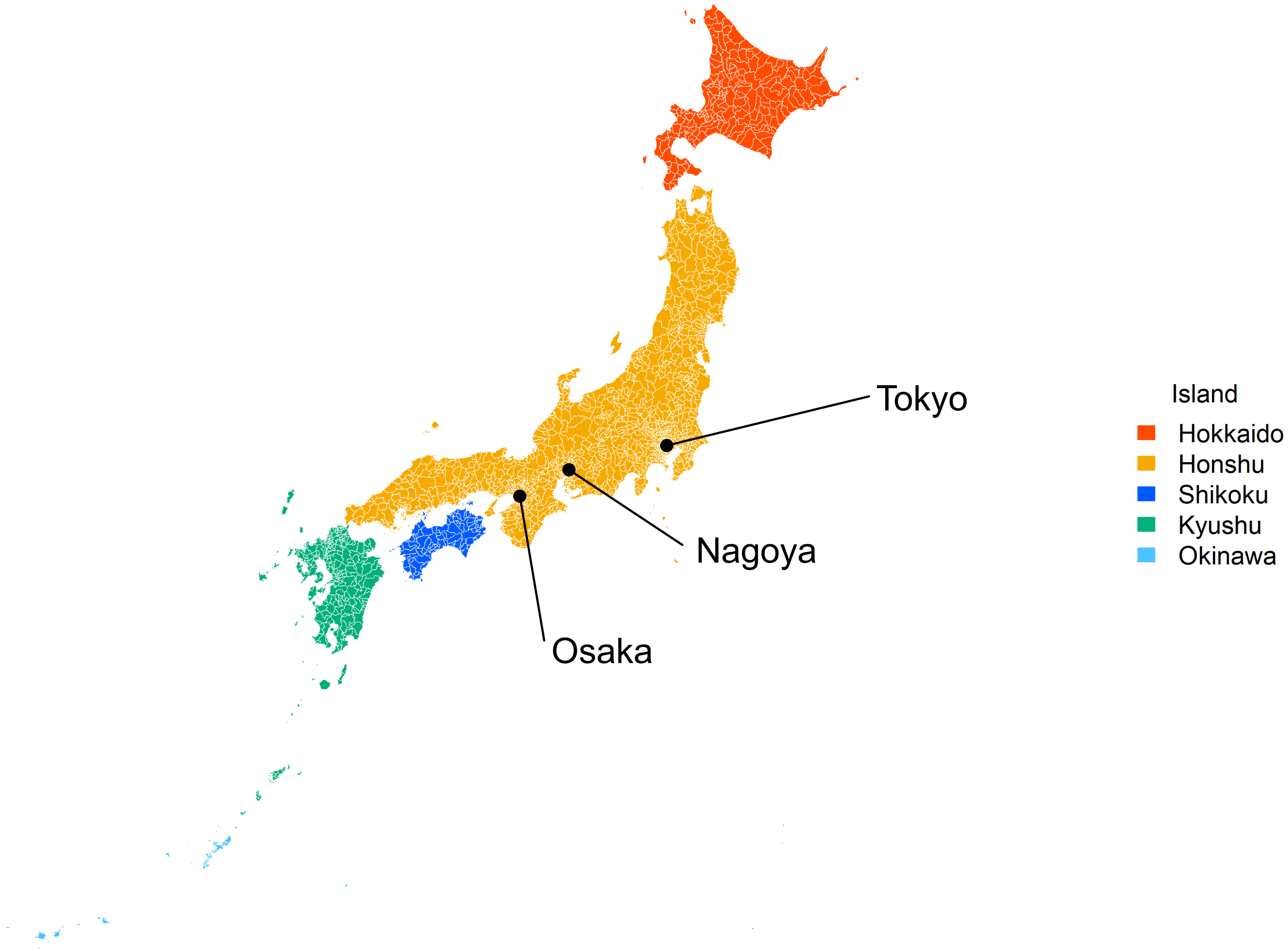

**Supplementary Figure 5. Geographic Distribution of FBSIR/FBSMR and Their Exceedance Probabilities by Sex and Municipality for Each Cancer**

Color-coded maps of FBSIR and FBSMR (ranging from red [high] to blue [low]) and their exceedance probabilities of all cancers are shown for males (A) and females (B). The exceedance probabilities represent the probabilities of FBSIR/FBSMR values being greater than 1.1 within individual municipalities. Major central cities and their surrounding areas are broken out. As this study follows census-based regional definitions for consistency, some discrepancies between municipality names and their corresponding prefectures arise (e.g., 'Hiroshima' refers to the Hiroshima metropolitan area rather than Hiroshima Prefecture). FBSIR, fully Bayesian estimate of standardized incidence ratio; FBSMR, fully Bayesian estimate of standardized mortality ratio. (A) Stomach; (B) Colorectum; (C) Lung; (D) Liver; (E) Pancreas; (F) Breast; (G) Prostate; (H) Cervix.

### (A) Stomach, Male

#### FBSIR

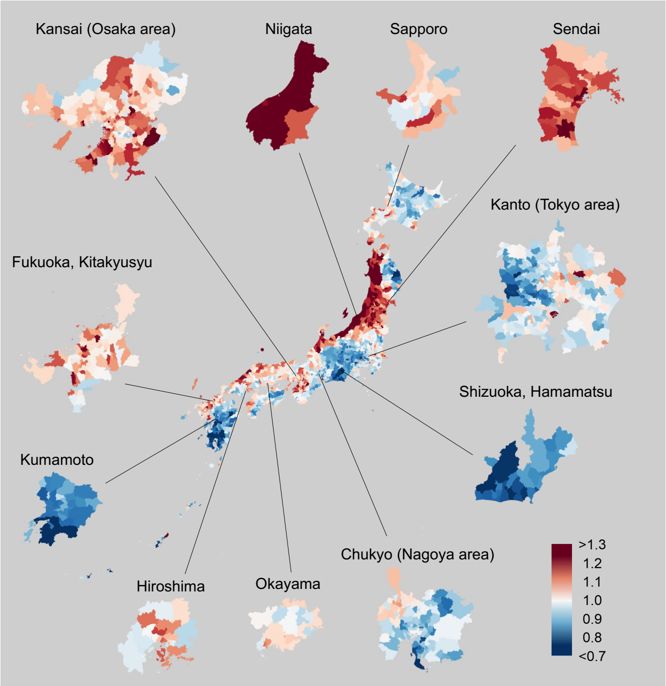

#### Exceedance probability of FBSIR

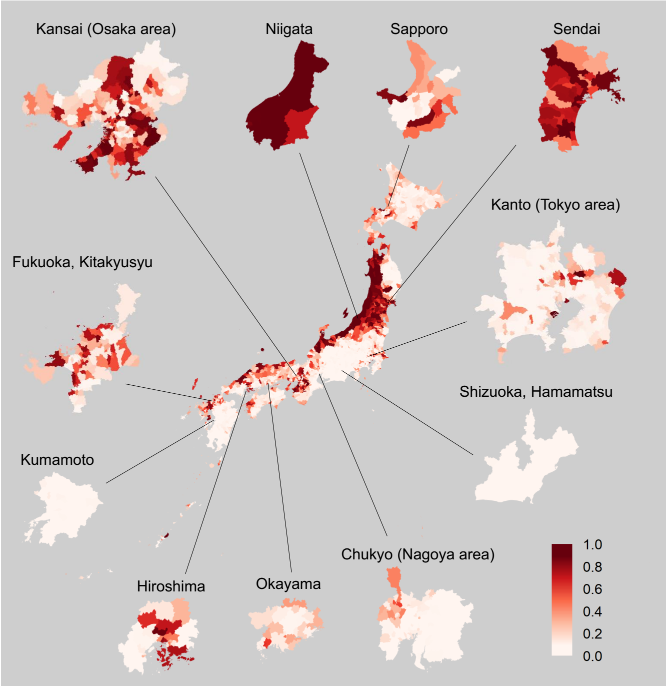

#### FBSMR

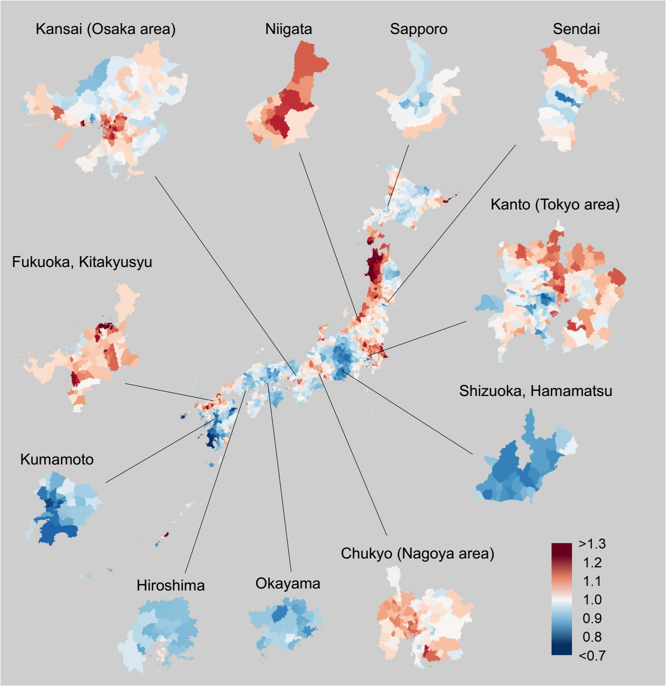

#### Exceedance probability of FBSMR

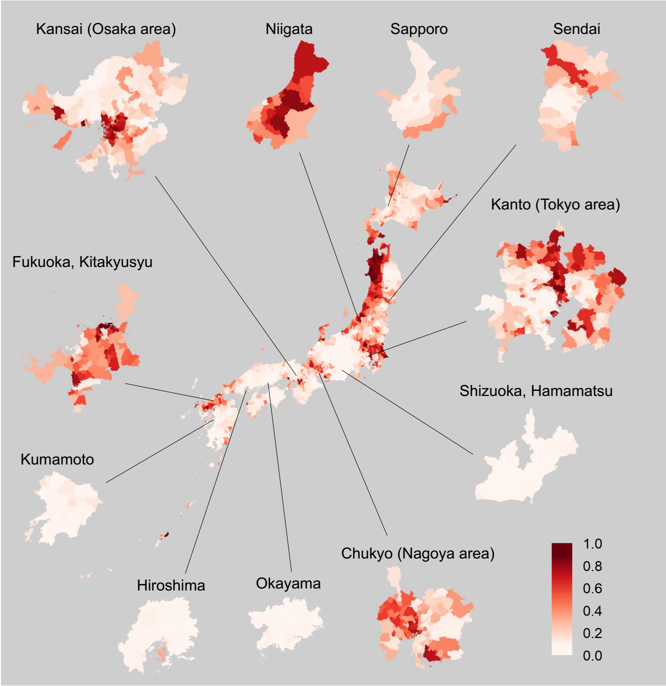

**Supplementary Figure 5. Geographic Distribution of FBSIR/FBSMR and Their Exceedance Probabilities by Sex and Municipality for Each Cancer**

Color-coded maps of FBSIR and FBSMR (ranging from red [high] to blue [low]) and their exceedance probabilities of all cancers are shown for males (A) and females (B). The exceedance probabilities represent the probabilities of FBSIR/FBSMR values being greater than 1.1 within individual municipalities. Major central cities and their surrounding areas are broken out. As this study follows census-based regional definitions for consistency, some discrepancies between municipality names and their corresponding prefectures arise (e.g., 'Hiroshima' refers to the Hiroshima metropolitan area rather than Hiroshima Prefecture). FBSIR, fully Bayesian estimate of standardized incidence ratio; FBSMR, fully Bayesian estimate of standardized mortality ratio. (A) Stomach; (B) Colorectum; (C) Lung; (D) Liver; (E) Pancreas; (F) Breast; (G) Prostate; (H) Cervix.

### (A) Stomach, Female

#### FBSIR

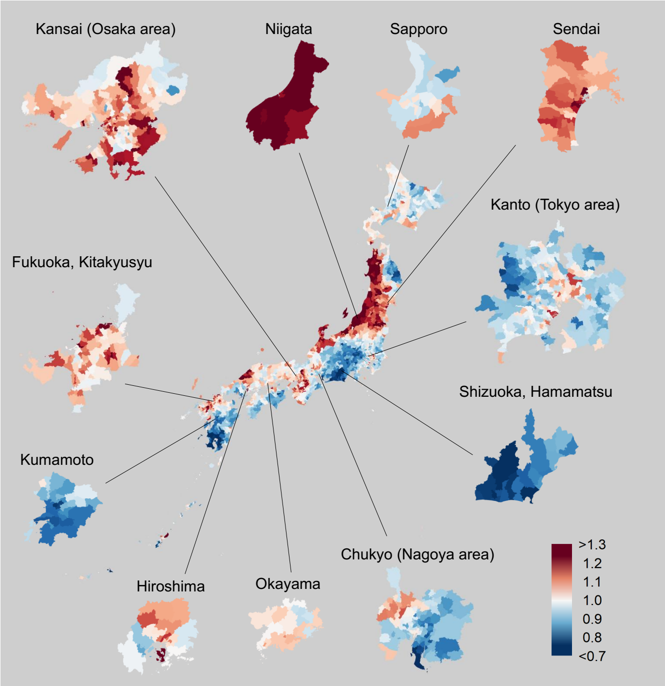

#### Exceedance probability of FBSIR

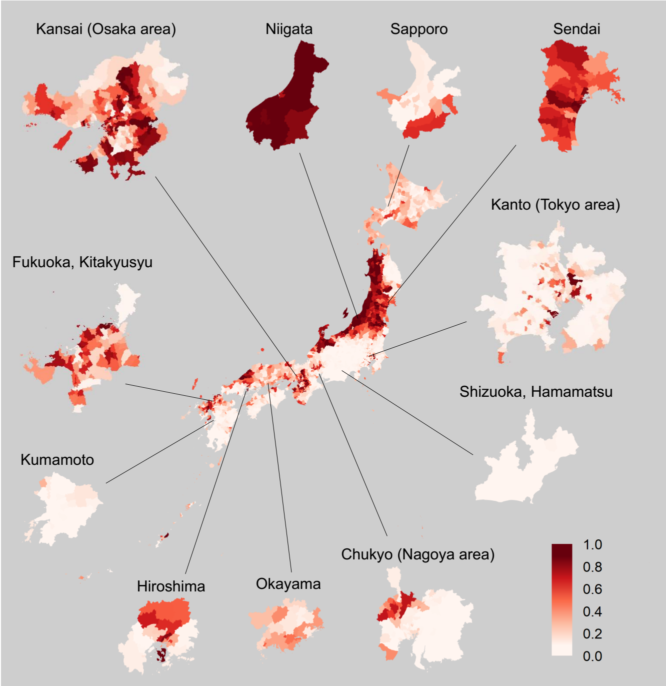

#### FBSMR

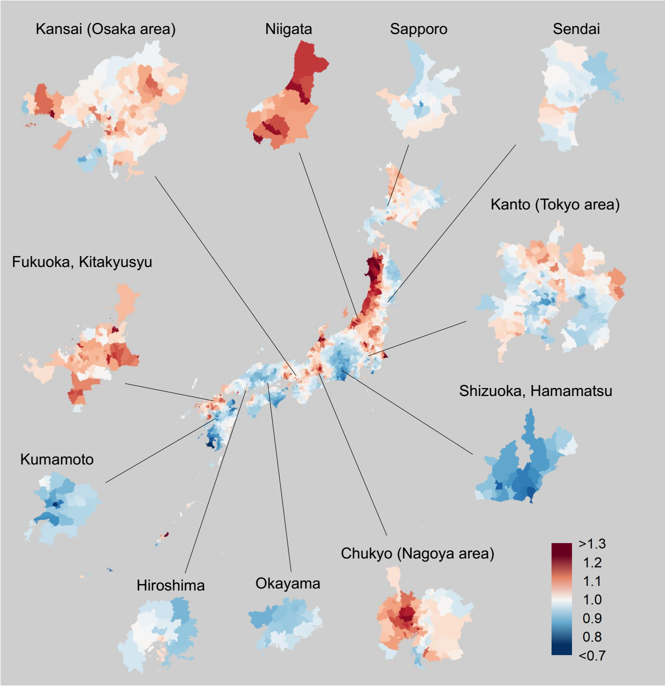

#### Exceedance probability of FBSMR

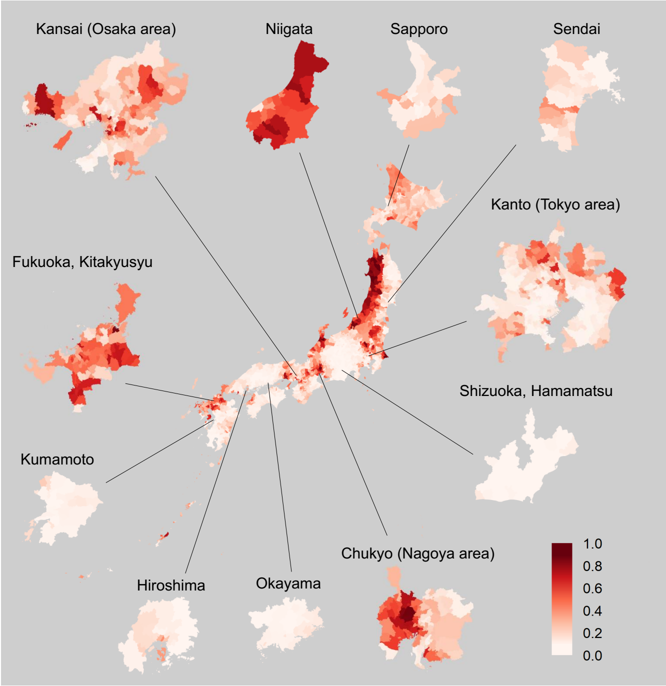

**Supplementary Figure 5. Geographic Distribution of FBSIR/FBSMR and Their Exceedance Probabilities by Sex and Municipality for Each Cancer**

Color-coded maps of FBSIR and FBSMR (ranging from red [high] to blue [low]) and their exceedance probabilities of all cancers are shown for males (A) and females (B). The exceedance probabilities represent the probabilities of FBSIR/FBSMR values being greater than 1.1 within individual municipalities. Major central cities and their surrounding areas are broken out. As this study follows census-based regional definitions for consistency, some discrepancies between municipality names and their corresponding prefectures arise (e.g., 'Hiroshima' refers to the Hiroshima metropolitan area rather than Hiroshima Prefecture). FBSIR, fully Bayesian estimate of standardized incidence ratio; FBSMR, fully Bayesian estimate of standardized mortality ratio. (A) Stomach; (B) Colorectum; (C) Lung; (D) Liver; (E) Pancreas; (F) Breast; (G) Prostate; (H) Cervix.

### (B) Colorectum, Male

#### FBSIR

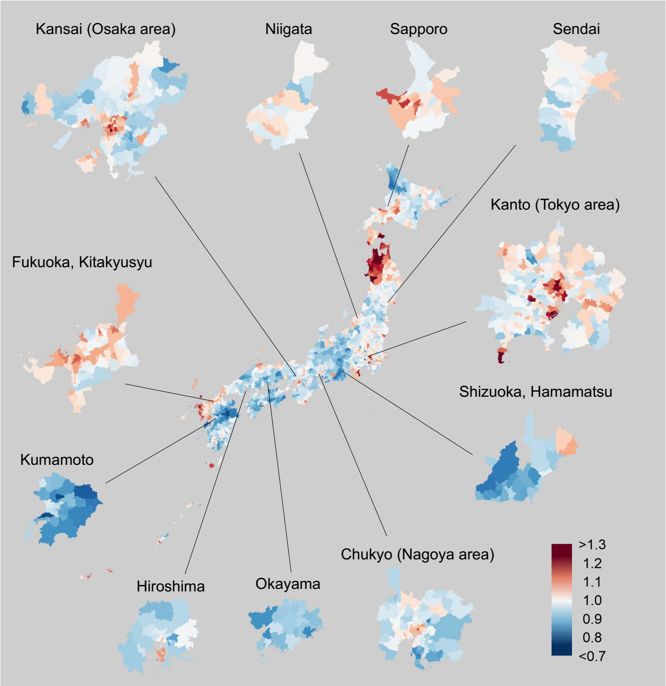

#### Exceedance probability of FBSIR

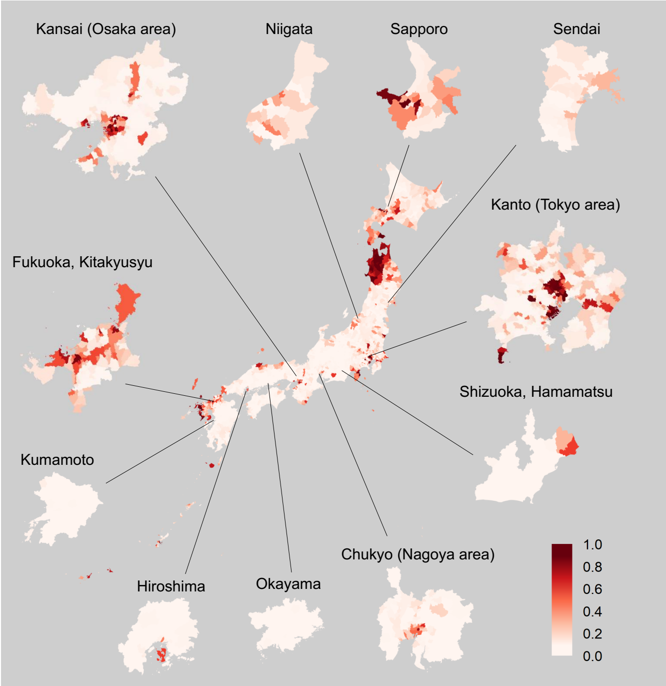

#### FBSMR

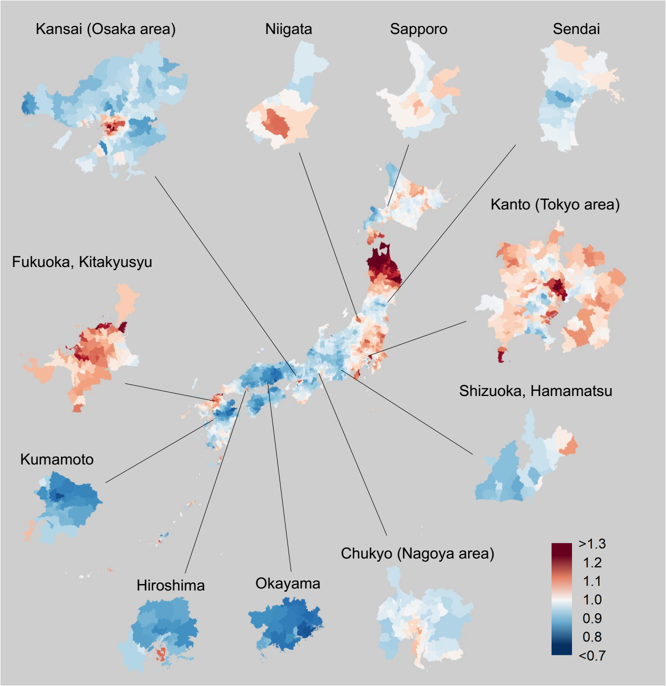

#### Exceedance probability of FBSMR

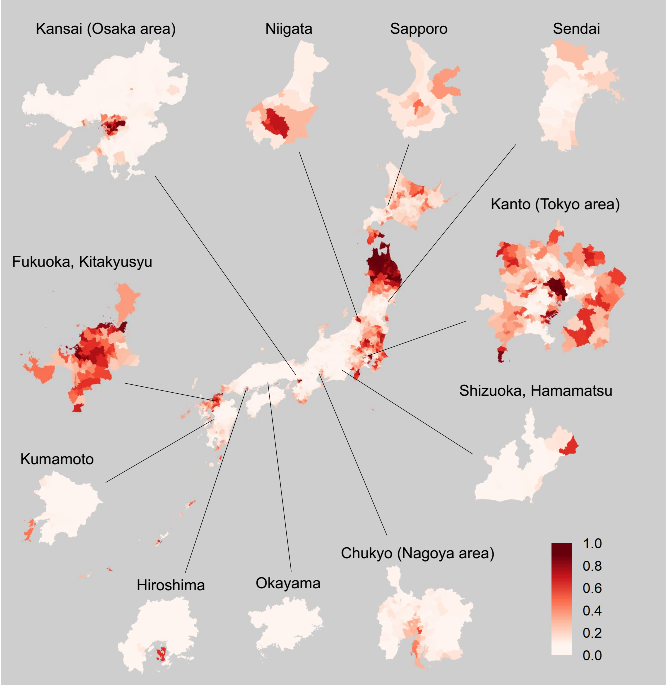

**Supplementary Figure 5. Geographic Distribution of FBSIR/FBSMR and Their Exceedance Probabilities by Sex and Municipality for Each Cancer**

Color-coded maps of FBSIR and FBSMR (ranging from red [high] to blue [low]) and their exceedance probabilities of all cancers are shown for males (A) and females (B). The exceedance probabilities represent the probabilities of FBSIR/FBSMR values being greater than 1.1 within individual municipalities. Major central cities and their surrounding areas are broken out. As this study follows census-based regional definitions for consistency, some discrepancies between municipality names and their corresponding prefectures arise (e.g., 'Hiroshima' refers to the Hiroshima metropolitan area rather than Hiroshima Prefecture). FBSIR, fully Bayesian estimate of standardized incidence ratio; FBSMR, fully Bayesian estimate of standardized mortality ratio. (A) Stomach; (B) Colorectum; (C) Lung; (D) Liver; (E) Pancreas; (F) Breast; (G) Prostate; (H) Cervix.

### (B) Colorectum, Female

#### FBSIR

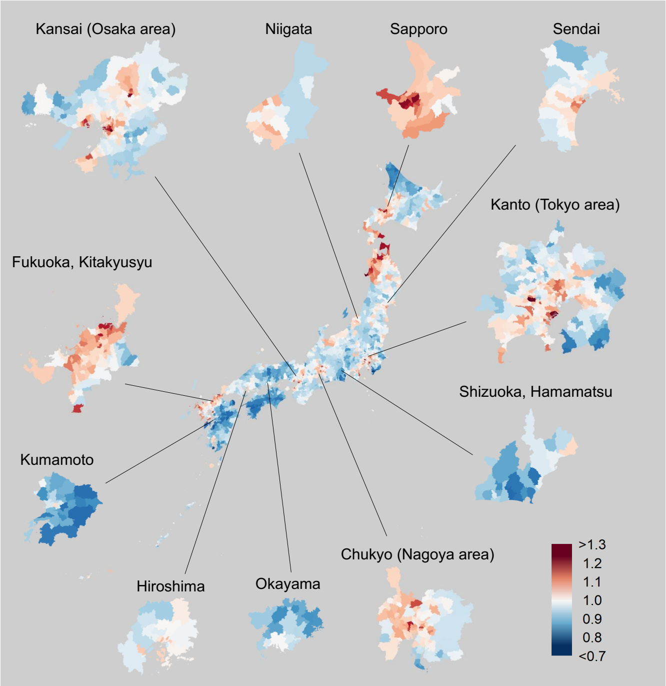

#### Exceedance probability of FBSIR

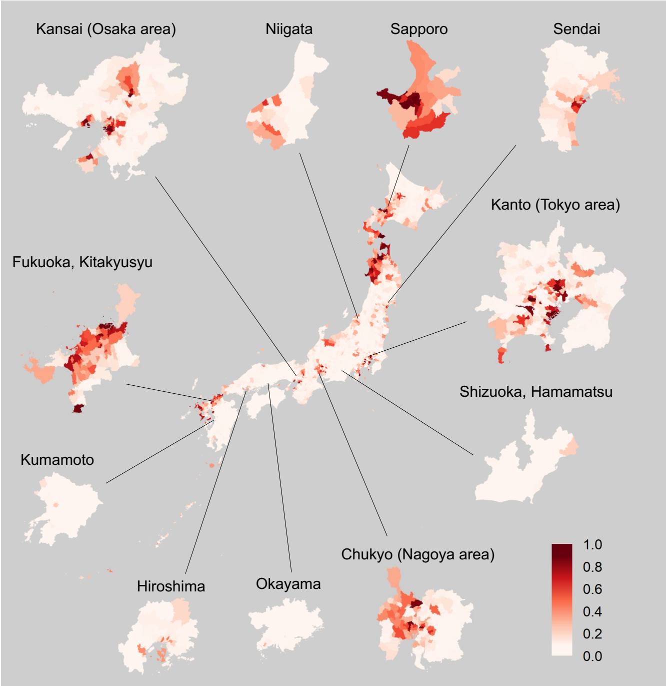

#### FBSMR

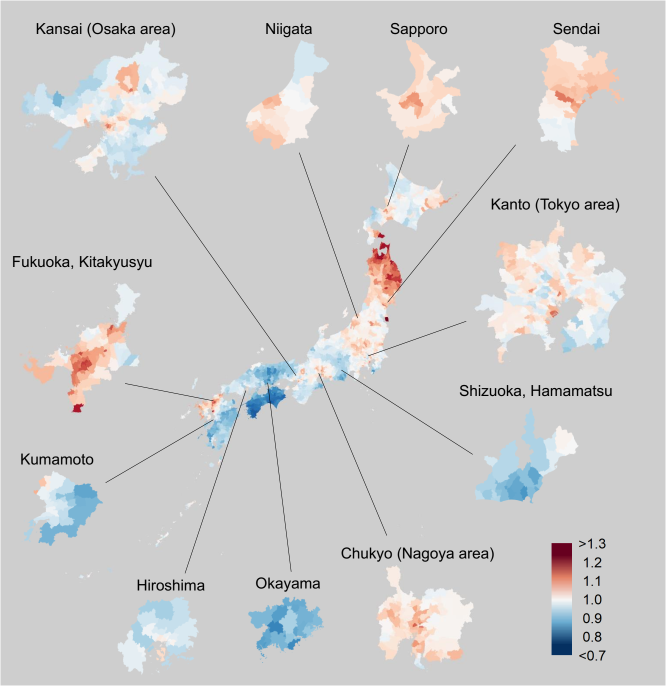

#### Exceedance probability of FBSMR

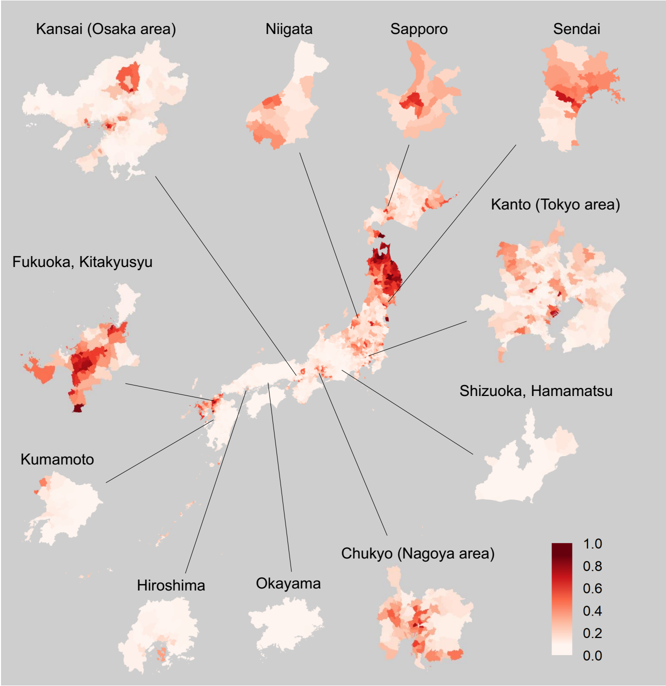

**Supplementary Figure 5. Geographic Distribution of FBSIR/FBSMR and Their Exceedance Probabilities by Sex and Municipality for Each Cancer**

Color-coded maps of FBSIR and FBSMR (ranging from red [high] to blue [low]) and their exceedance probabilities of all cancers are shown for males (A) and females (B). The exceedance probabilities represent the probabilities of FBSIR/FBSMR values being greater than 1.1 within individual municipalities. Major central cities and their surrounding areas are broken out. As this study follows census-based regional definitions for consistency, some discrepancies between municipality names and their corresponding prefectures arise (e.g., 'Hiroshima' refers to the Hiroshima metropolitan area rather than Hiroshima Prefecture). FBSIR, fully Bayesian estimate of standardized incidence ratio; FBSMR, fully Bayesian estimate of standardized mortality ratio. (A) Stomach; (B) Colorectum; (C) Lung; (D) Liver; (E) Pancreas; (F) Breast; (G) Prostate; (H) Cervix.

### (C) Lung, Male

#### FBSIR

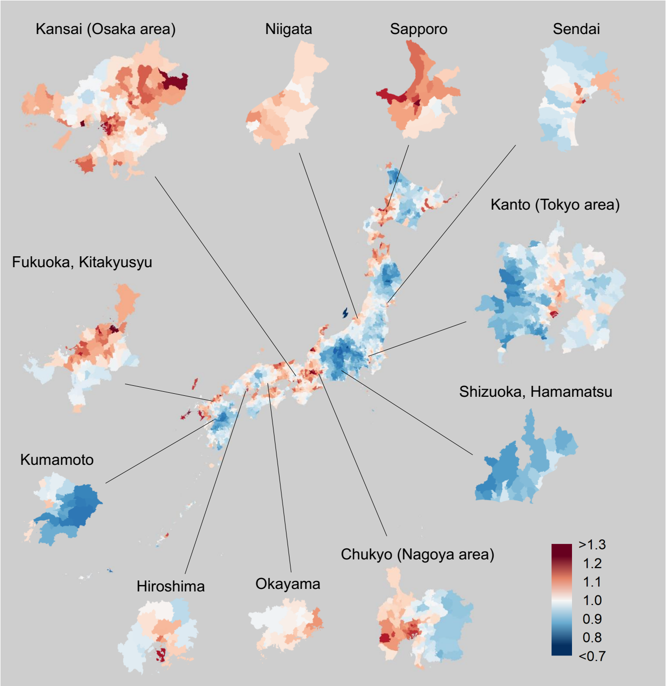

#### Exceedance probability of FBSIR

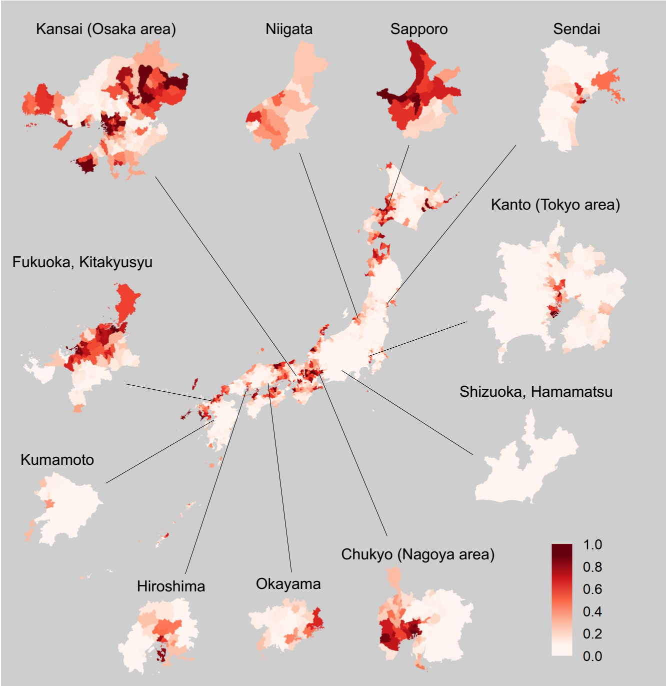

#### FBSMR

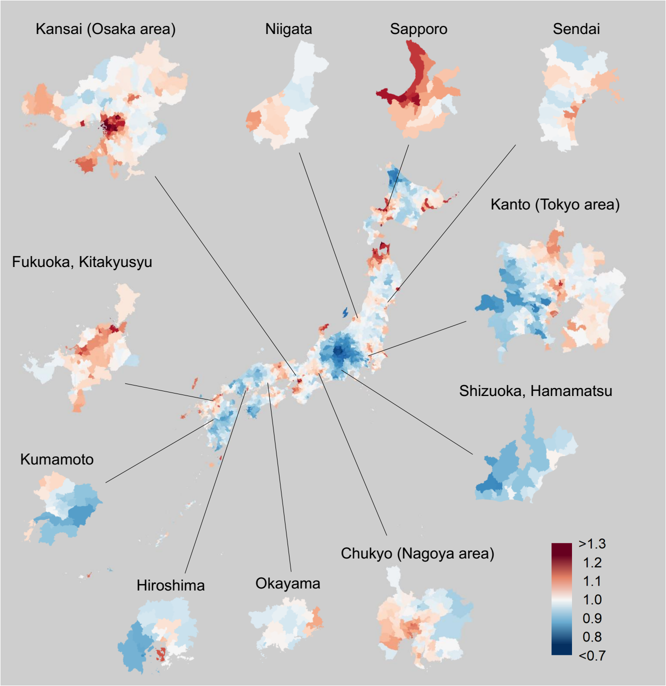

#### Exceedance probability of FBSMR

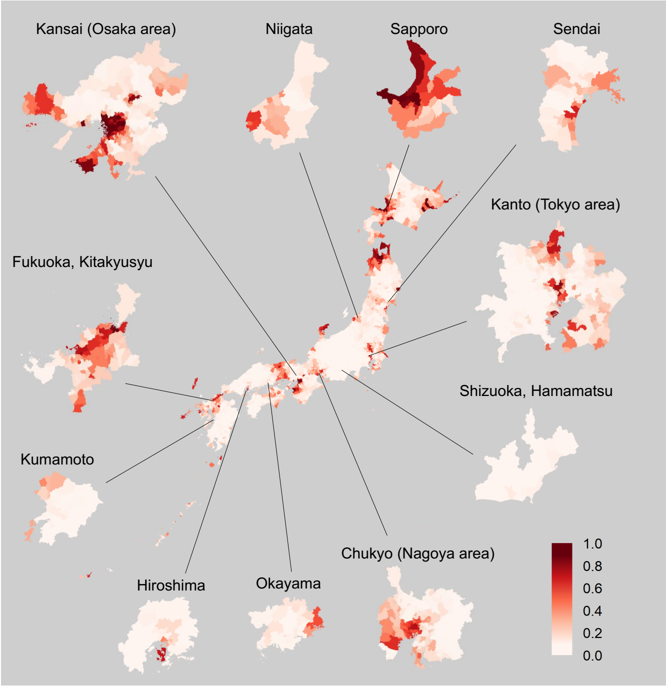

**Supplementary Figure 5. Geographic Distribution of FBSIR/FBSMR and Their Exceedance Probabilities by Sex and Municipality for Each Cancer**

Color-coded maps of FBSIR and FBSMR (ranging from red [high] to blue [low]) and their exceedance probabilities of all cancers are shown for males (A) and females (B). The exceedance probabilities represent the probabilities of FBSIR/FBSMR values being greater than 1.1 within individual municipalities. Major central cities and their surrounding areas are broken out. As this study follows census-based regional definitions for consistency, some discrepancies between municipality names and their corresponding prefectures arise (e.g., 'Hiroshima' refers to the Hiroshima metropolitan area rather than Hiroshima Prefecture). FBSIR, fully Bayesian estimate of standardized incidence ratio; FBSMR, fully Bayesian estimate of standardized mortality ratio. (A) Stomach; (B) Colorectum; (C) Lung; (D) Liver; (E) Pancreas; (F) Breast; (G) Prostate; (H) Cervix.

### (C) Lung, Female

#### FBSIR

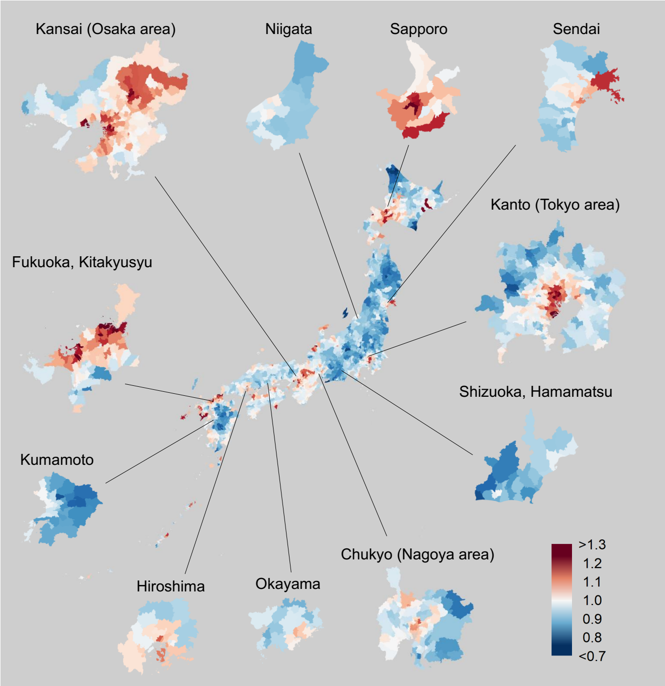

#### Exceedance probability of FBSIR

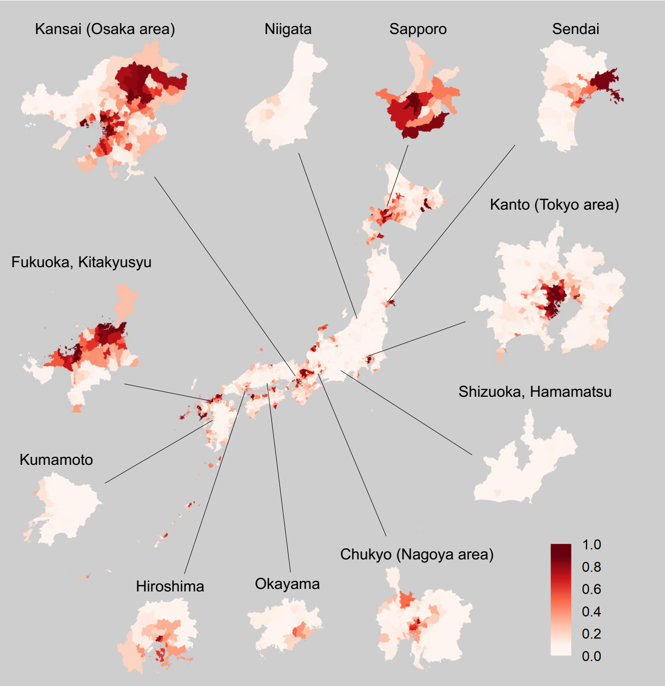

#### FBSMR

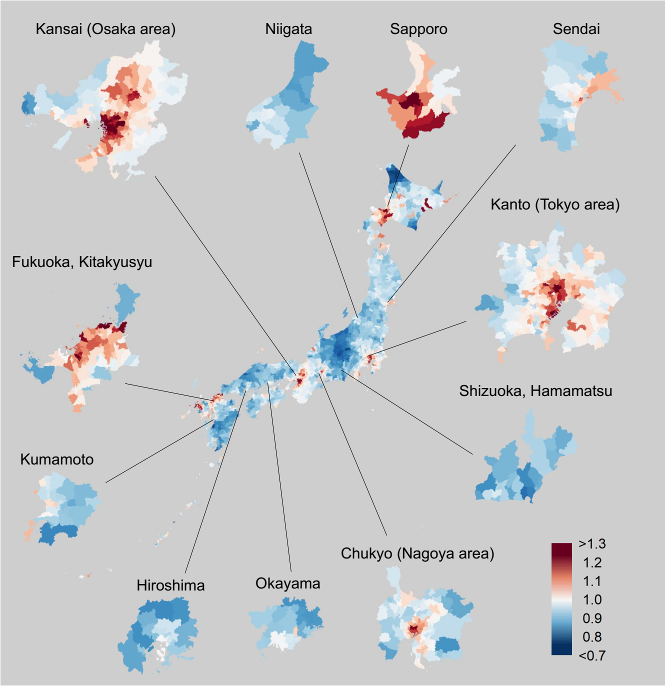

#### Exceedance probability of FBSMR

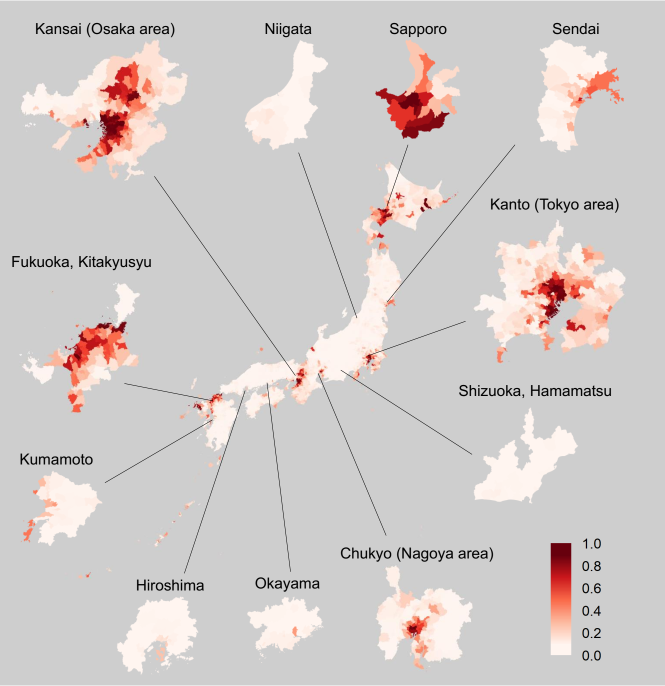

**Supplementary Figure 5. Geographic Distribution of FBSIR/FBSMR and Their Exceedance Probabilities by Sex and Municipality for Each Cancer**

Color-coded maps of FBSIR and FBSMR (ranging from red [high] to blue [low]) and their exceedance probabilities of all cancers are shown for males (A) and females (B). The exceedance probabilities represent the probabilities of FBSIR/FBSMR values being greater than 1.1 within individual municipalities. Major central cities and their surrounding areas are broken out. As this study follows census-based regional definitions for consistency, some discrepancies between municipality names and their corresponding prefectures arise (e.g., 'Hiroshima' refers to the Hiroshima metropolitan area rather than Hiroshima Prefecture). FBSIR, fully Bayesian estimate of standardized incidence ratio; FBSMR, fully Bayesian estimate of standardized mortality ratio. (A) Stomach; (B) Colorectum; (C) Lung; (D) Liver; (E) Pancreas; (F) Breast; (G) Prostate; (H) Cervix.

### (D) Liver, Male

#### FBSIR

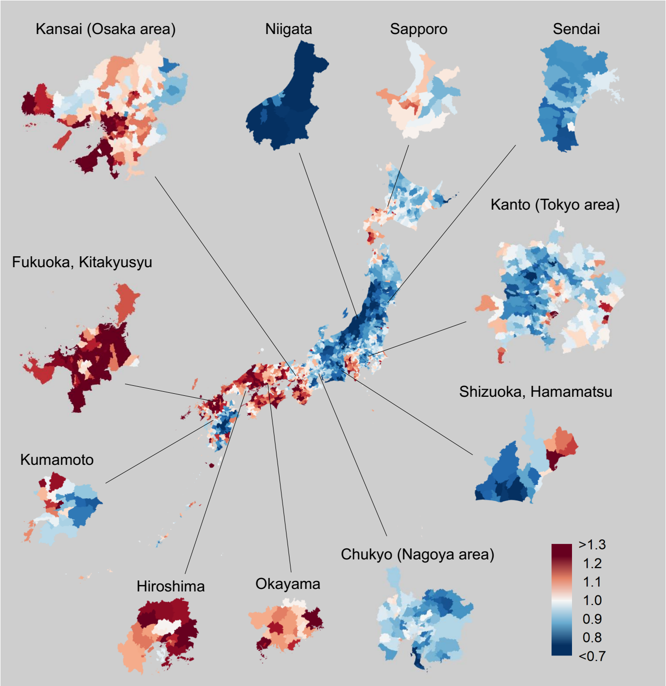

#### Exceedance probability of FBSIR

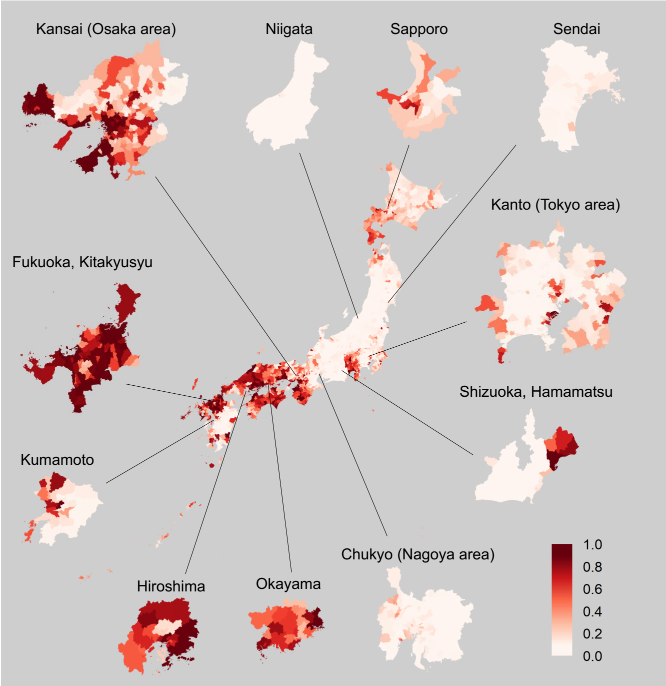

#### FBSMR

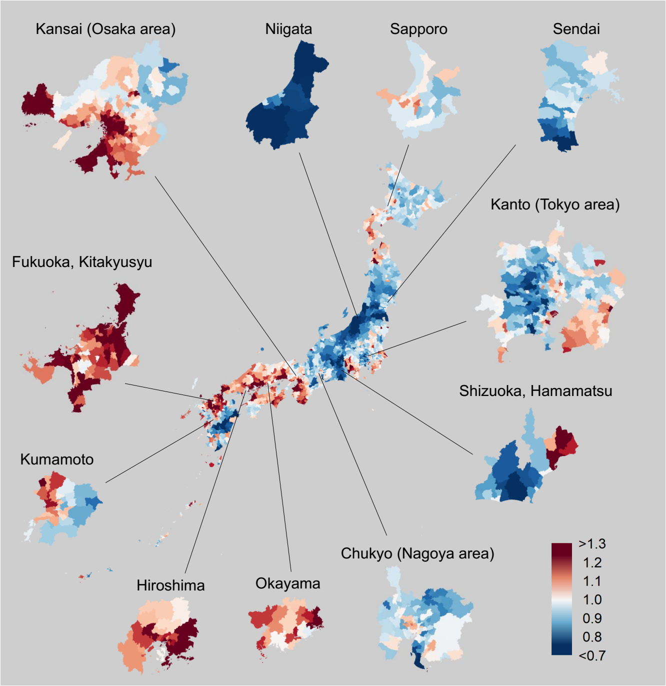

#### Exceedance probability of FBSMR

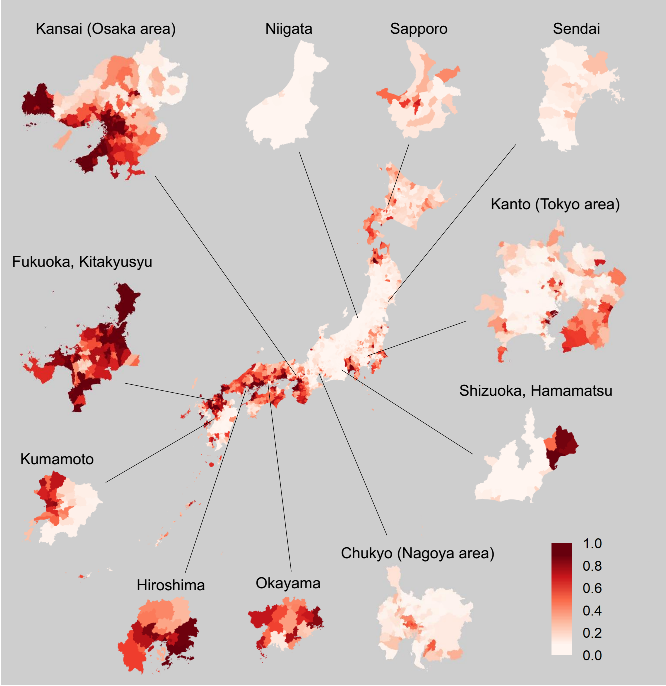

**Supplementary Figure 5. Geographic Distribution of FBSIR/FBSMR and Their Exceedance Probabilities by Sex and Municipality for Each Cancer**

Color-coded maps of FBSIR and FBSMR (ranging from red [high] to blue [low]) and their exceedance probabilities of all cancers are shown for males (A) and females (B). The exceedance probabilities represent the probabilities of FBSIR/FBSMR values being greater than 1.1 within individual municipalities. Major central cities and their surrounding areas are broken out. As this study follows census-based regional definitions for consistency, some discrepancies between municipality names and their corresponding prefectures arise (e.g., 'Hiroshima' refers to the Hiroshima metropolitan area rather than Hiroshima Prefecture). FBSIR, fully Bayesian estimate of standardized incidence ratio; FBSMR, fully Bayesian estimate of standardized mortality ratio. (A) Stomach; (B) Colorectum; (C) Lung; (D) Liver; (E) Pancreas; (F) Breast; (G) Prostate; (H) Cervix.

### (D) Liver, Female

#### FBSIR

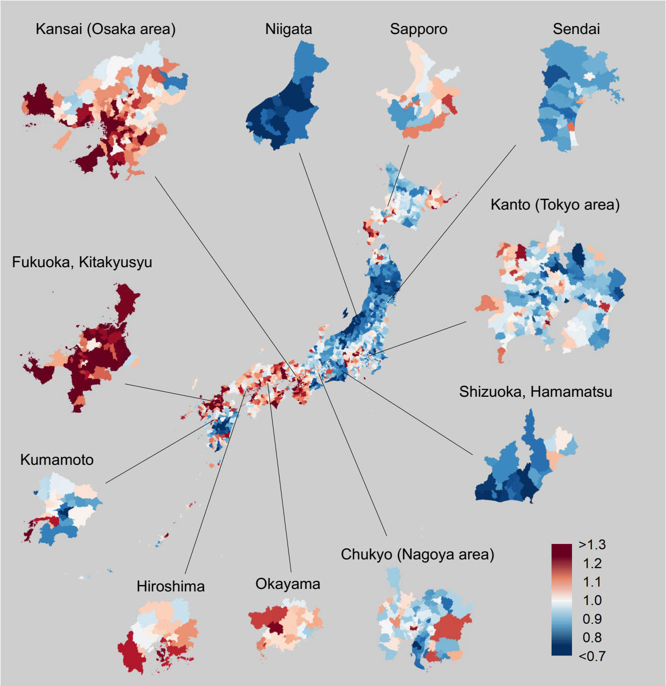

#### Exceedance probability of FBSIR

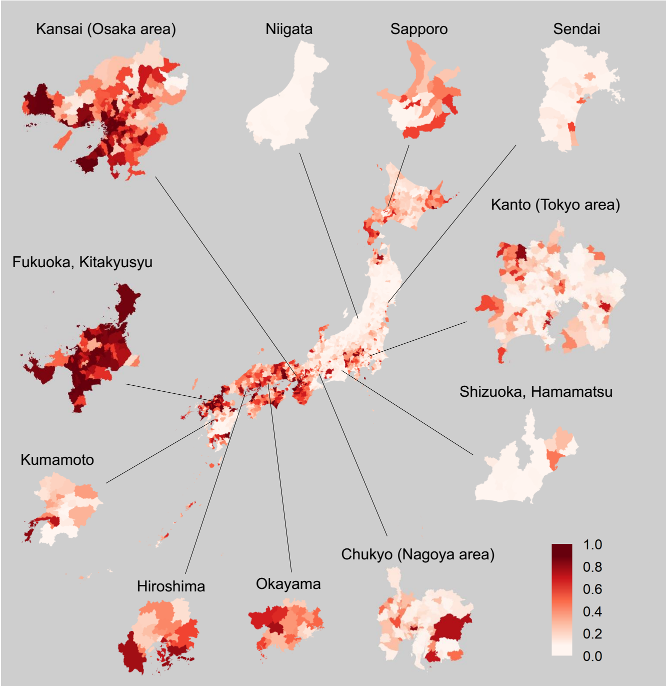

#### FBSMR

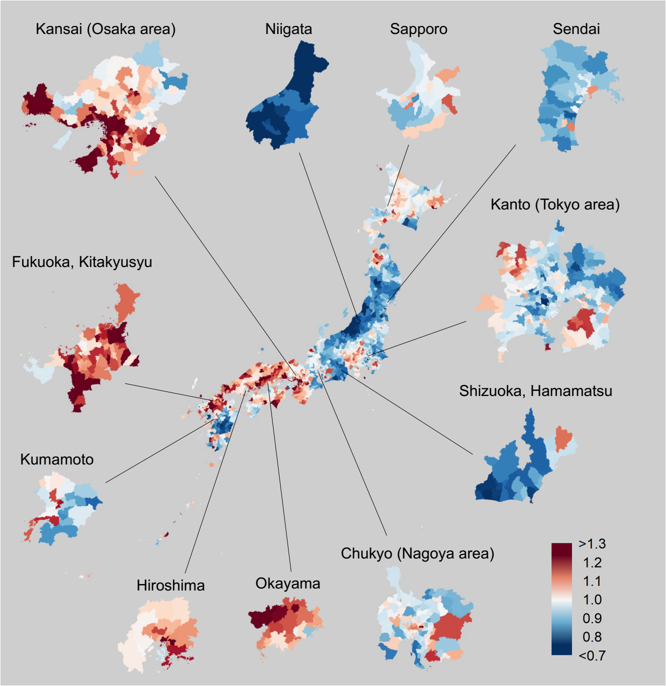

#### Exceedance probability of FBSMR

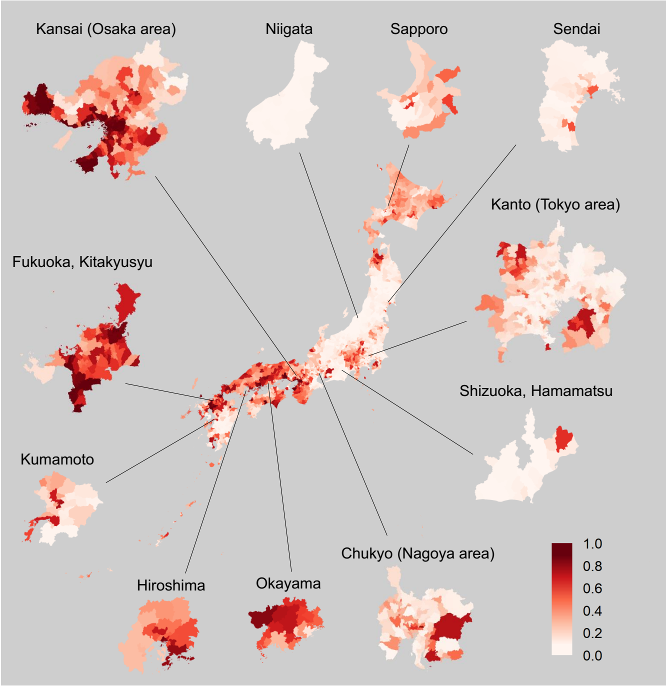

**Supplementary Figure 5. Geographic Distribution of FBSIR/FBSMR and Their Exceedance Probabilities by Sex and Municipality for Each Cancer**

Color-coded maps of FBSIR and FBSMR (ranging from red [high] to blue [low]) and their exceedance probabilities of all cancers are shown for males (A) and females (B). The exceedance probabilities represent the probabilities of FBSIR/FBSMR values being greater than 1.1 within individual municipalities. Major central cities and their surrounding areas are broken out. As this study follows census-based regional definitions for consistency, some discrepancies between municipality names and their corresponding prefectures arise (e.g., 'Hiroshima' refers to the Hiroshima metropolitan area rather than Hiroshima Prefecture). FBSIR, fully Bayesian estimate of standardized incidence ratio; FBSMR, fully Bayesian estimate of standardized mortality ratio. (A) Stomach; (B) Colorectum; (C) Lung; (D) Liver; (E) Pancreas; (F) Breast; (G) Prostate; (H) Cervix.

### (E) Pancreas, Male

#### FBSIR

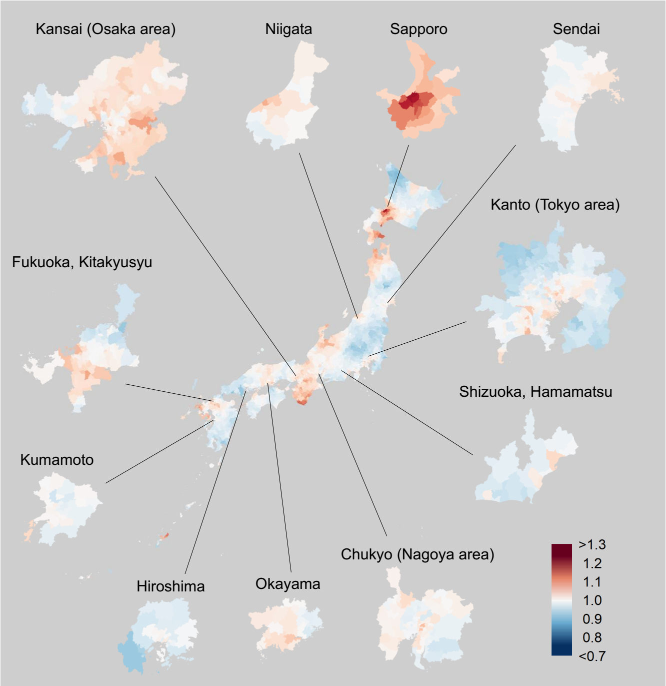

#### Exceedance probability of FBSIR

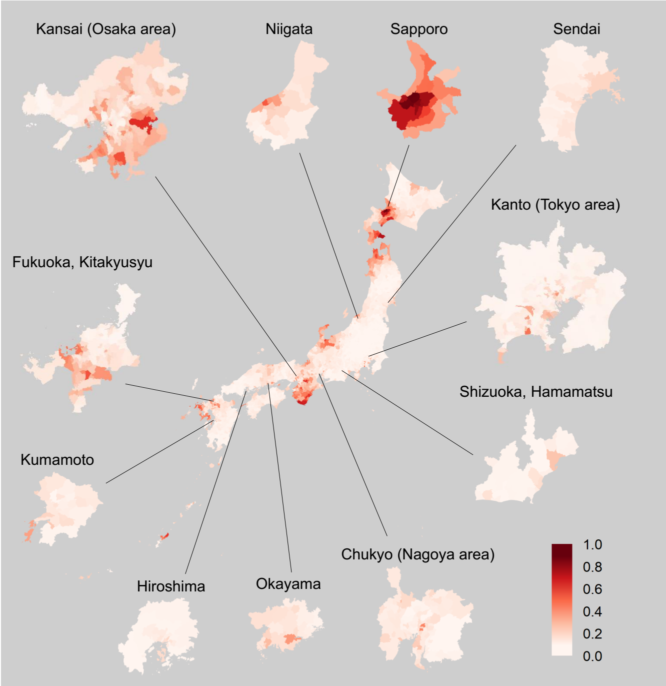

#### FBSMR

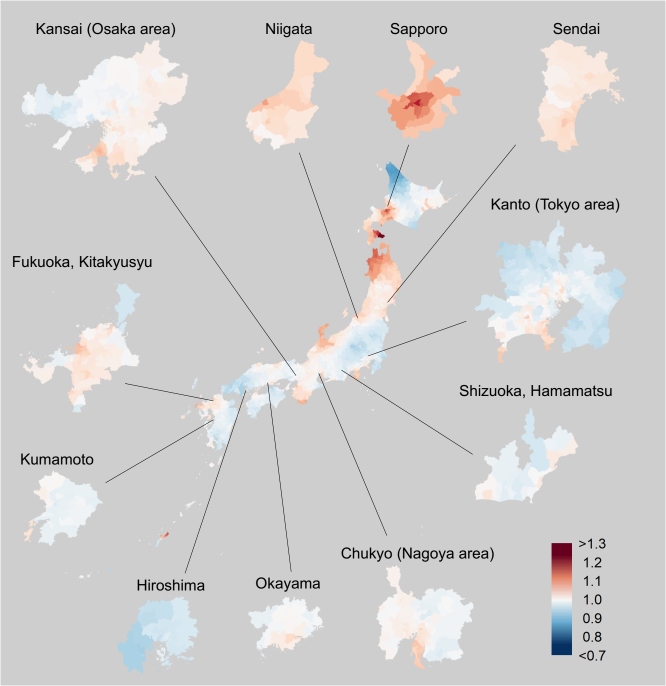

#### Exceedance probability of FBSMR

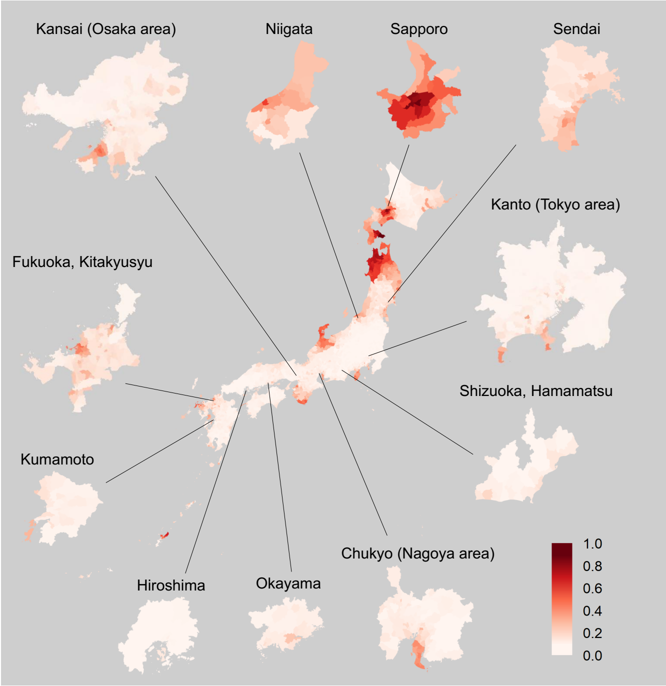

**Supplementary Figure 5. Geographic Distribution of FBSIR/FBSMR and Their Exceedance Probabilities by Sex and Municipality for Each Cancer**

Color-coded maps of FBSIR and FBSMR (ranging from red [high] to blue [low]) and their exceedance probabilities of all cancers are shown for males (A) and females (B). The exceedance probabilities represent the probabilities of FBSIR/FBSMR values being greater than 1.1 within individual municipalities. Major central cities and their surrounding areas are broken out. As this study follows census-based regional definitions for consistency, some discrepancies between municipality names and their corresponding prefectures arise (e.g., 'Hiroshima' refers to the Hiroshima metropolitan area rather than Hiroshima Prefecture). FBSIR, fully Bayesian estimate of standardized incidence ratio; FBSMR, fully Bayesian estimate of standardized mortality ratio. (A) Stomach; (B) Colorectum; (C) Lung; (D) Liver; (E) Pancreas; (F) Breast; (G) Prostate; (H) Cervix.

### (E) Pancreas, Female

#### FBSIR

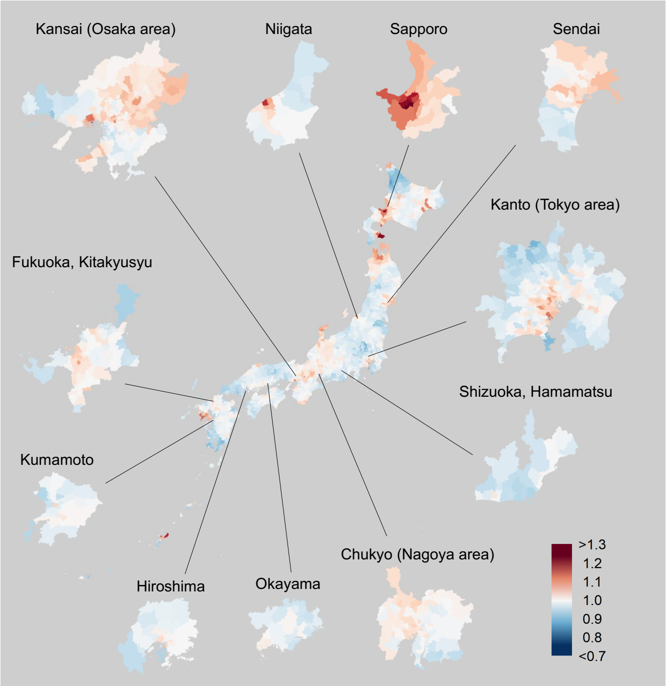

#### Exceedance probability of FBSIR

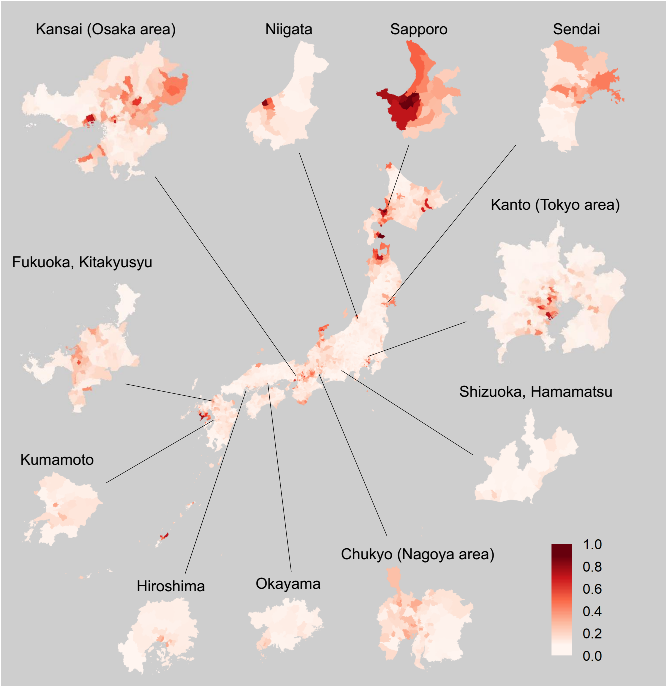

#### FBSMR

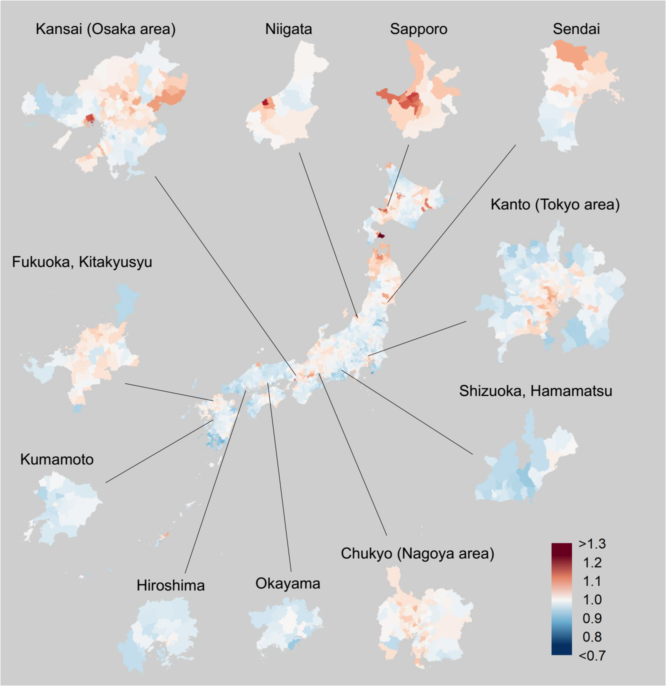

#### Exceedance probability of FBSMR

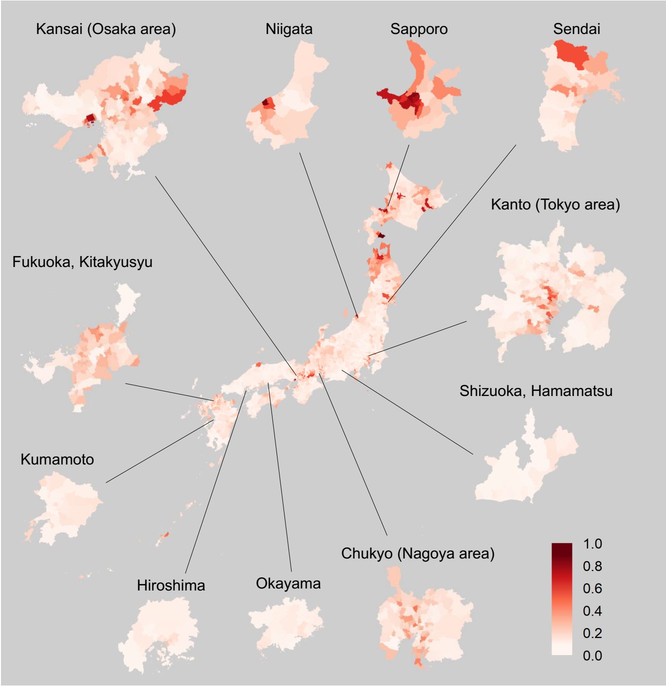

**Supplementary Figure 5. Geographic Distribution of FBSIR/FBSMR and Their Exceedance Probabilities by Sex and Municipality for Each Cancer**

Color-coded maps of FBSIR and FBSMR (ranging from red [high] to blue [low]) and their exceedance probabilities of all cancers are shown for males (A) and females (B). The exceedance probabilities represent the probabilities of FBSIR/FBSMR values being greater than 1.1 within individual municipalities. Major central cities and their surrounding areas are broken out. As this study follows census-based regional definitions for consistency, some discrepancies between municipality names and their corresponding prefectures arise (e.g., 'Hiroshima' refers to the Hiroshima metropolitan area rather than Hiroshima Prefecture). FBSIR, fully Bayesian estimate of standardized incidence ratio; FBSMR, fully Bayesian estimate of standardized mortality ratio. (A) Stomach; (B) Colorectum; (C) Lung; (D) Liver; (E) Pancreas; (F) Breast; (G) Prostate; (H) Cervix.

### (F) Breast, Female

#### FBSIR

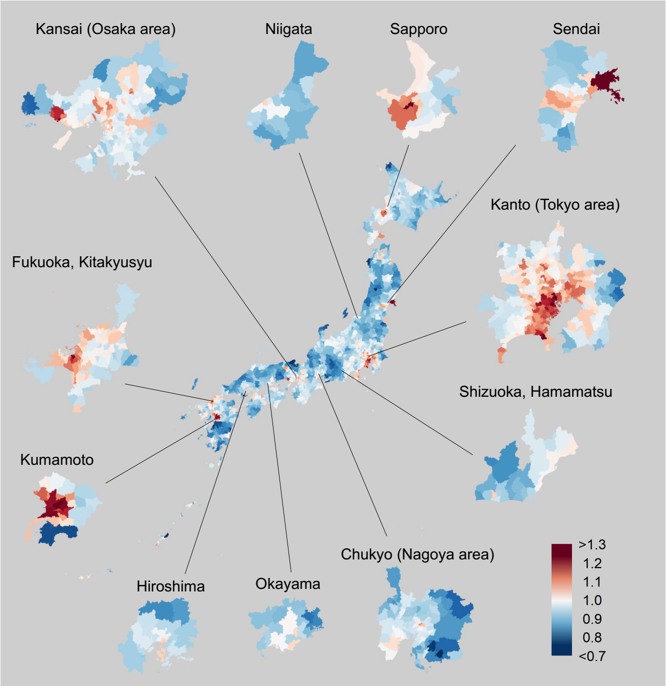

#### Exceedance probability of FBSIR

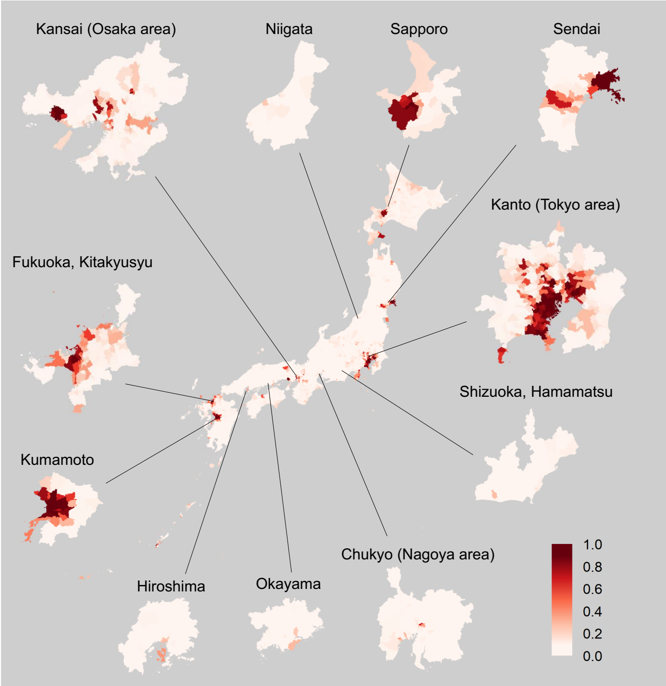

#### FBSMR

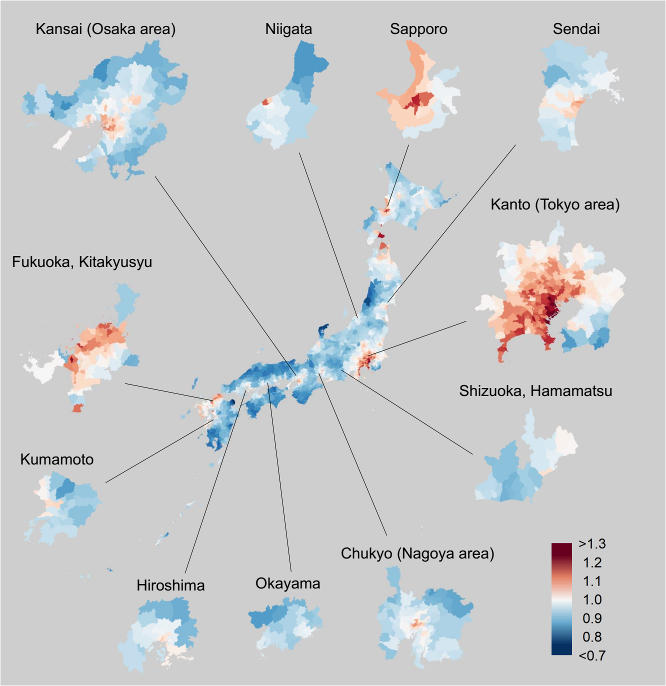

#### Exceedance probability of FBSMR

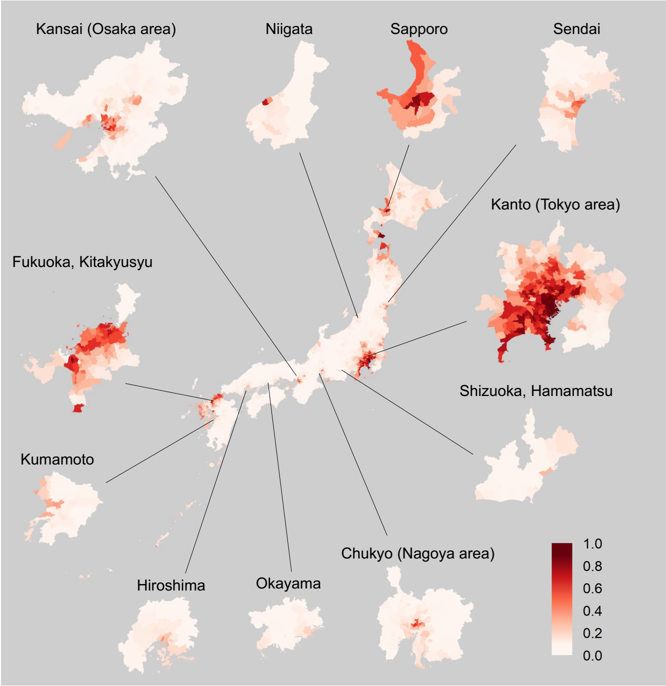

**Supplementary Figure 5. Geographic Distribution of FBSIR/FBSMR and Their Exceedance Probabilities by Sex and Municipality for Each Cancer**

Color-coded maps of FBSIR and FBSMR (ranging from red [high] to blue [low]) and their exceedance probabilities of all cancers are shown for males (A) and females (B). The exceedance probabilities represent the probabilities of FBSIR/FBSMR values being greater than 1.1 within individual municipalities. Major central cities and their surrounding areas are broken out. As this study follows census-based regional definitions for consistency, some discrepancies between municipality names and their corresponding prefectures arise (e.g., 'Hiroshima' refers to the Hiroshima metropolitan area rather than Hiroshima Prefecture). FBSIR, fully Bayesian estimate of standardized incidence ratio; FBSMR, fully Bayesian estimate of standardized mortality ratio. (A) Stomach; (B) Colorectum; (C) Lung; (D) Liver; (E) Pancreas; (F) Breast; (G) Prostate; (H) Cervix.

### (G) Prostate, Male

#### FBSIR

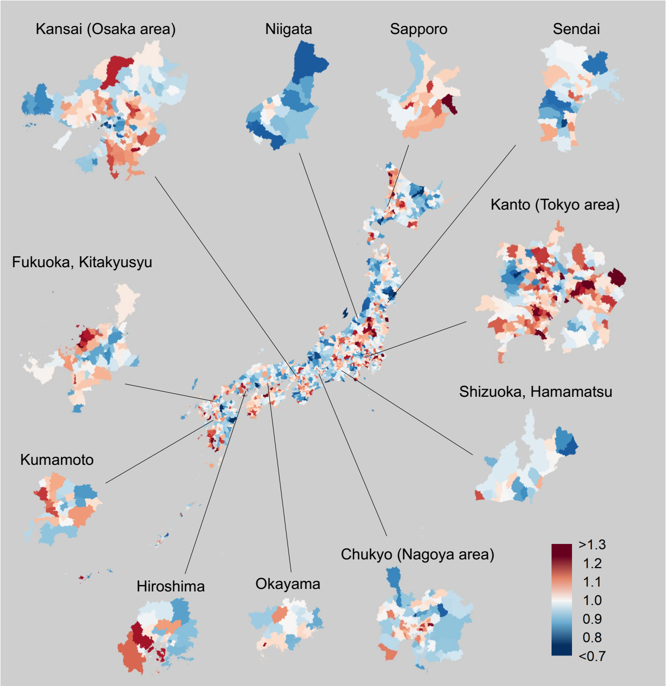

#### Exceedance probability of FBSIR

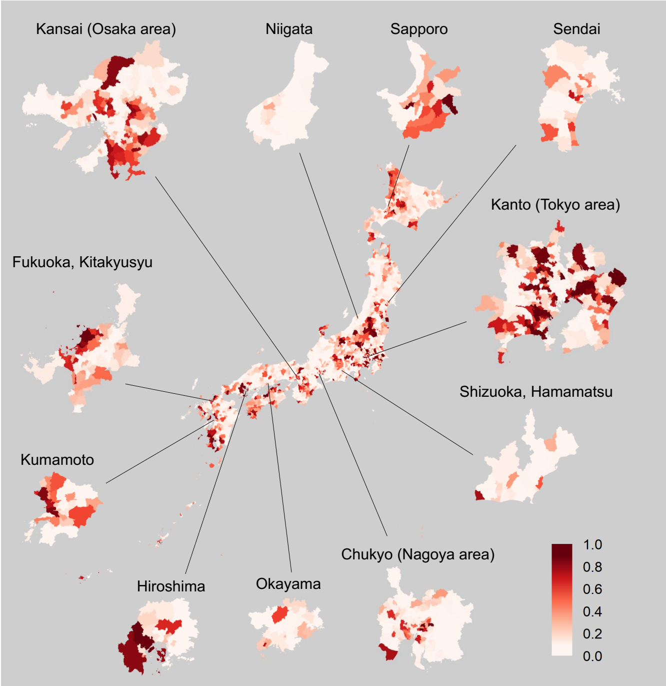

#### FBSMR

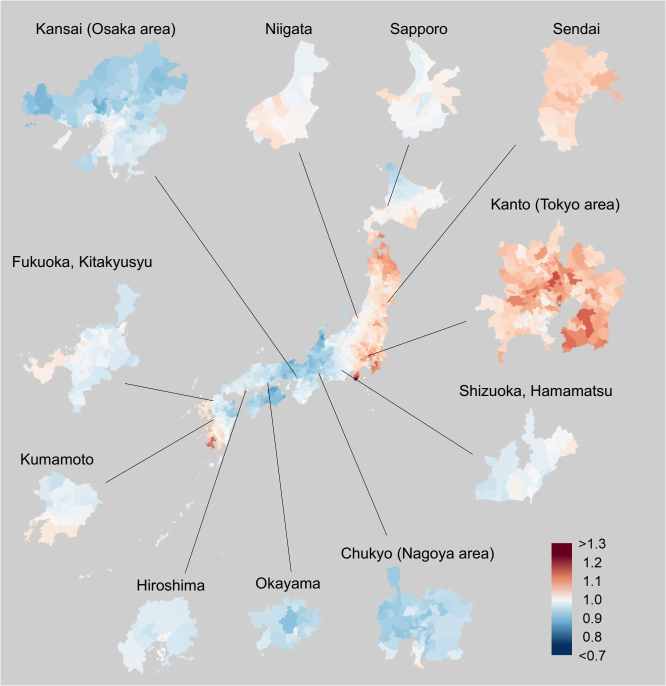

#### Exceedance probability of FBSMR

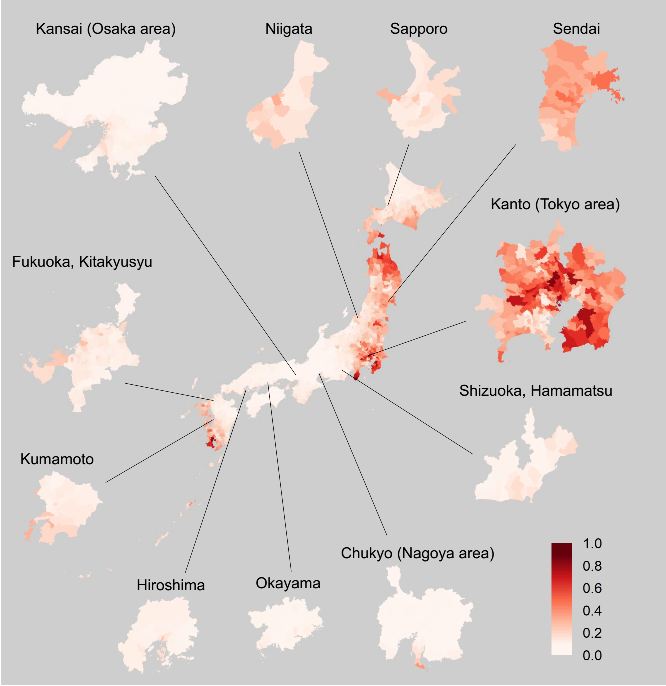

**Supplementary Figure 5. Geographic Distribution of FBSIR/FBSMR and Their Exceedance Probabilities by Sex and Municipality for Each Cancer**

Color-coded maps of FBSIR and FBSMR (ranging from red [high] to blue [low]) and their exceedance probabilities of all cancers are shown for males (A) and females (B). The exceedance probabilities represent the probabilities of FBSIR/FBSMR values being greater than 1.1 within individual municipalities. Major central cities and their surrounding areas are broken out. As this study follows census-based regional definitions for consistency, some discrepancies between municipality names and their corresponding prefectures arise (e.g., 'Hiroshima' refers to the Hiroshima metropolitan area rather than Hiroshima Prefecture). FBSIR, fully Bayesian estimate of standardized incidence ratio; FBSMR, fully Bayesian estimate of standardized mortality ratio. (A) Stomach; (B) Colorectum; (C) Lung; (D) Liver; (E) Pancreas; (F) Breast; (G) Prostate; (H) Cervix.

### (H) Cervix, Female

#### FBSIR

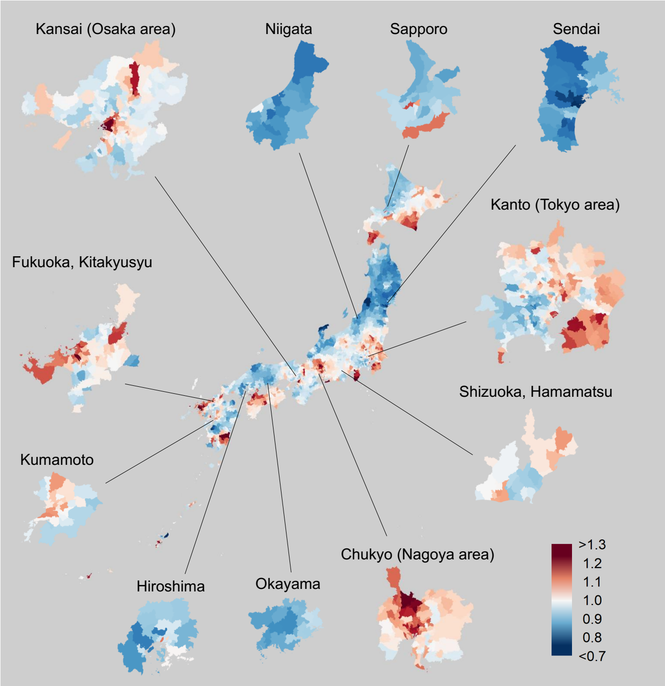

#### Exceedance probability of FBSIR

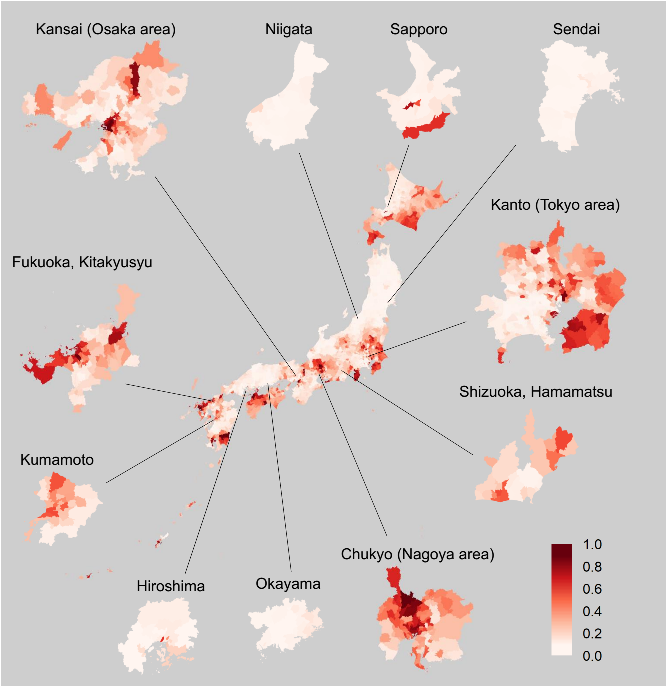

#### FBSMR

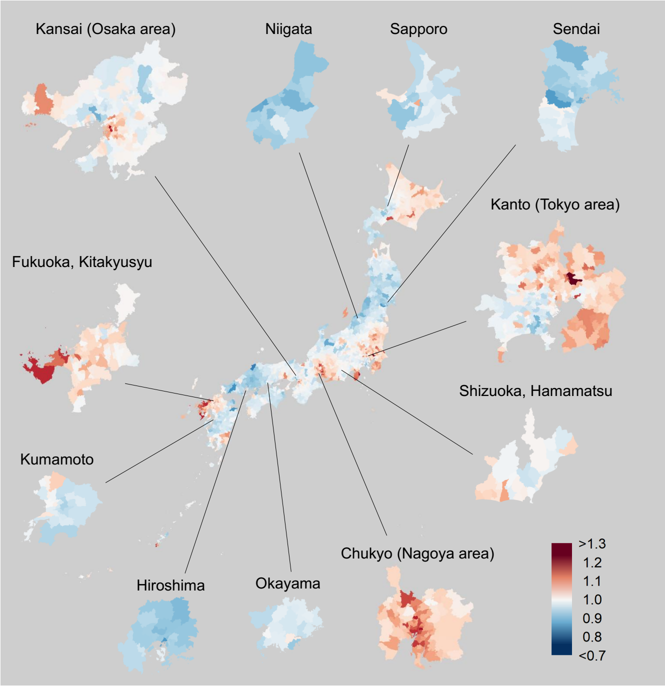

#### Exceedance probability of FBSMR

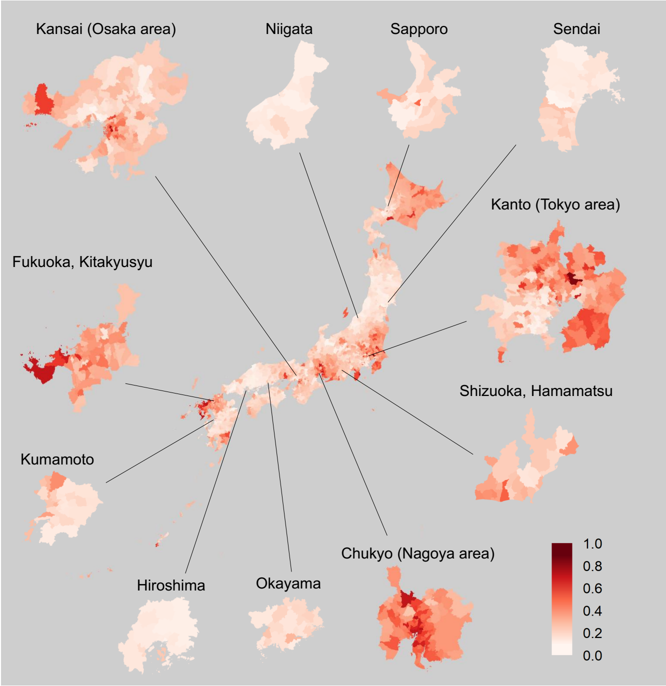

**Supplementary Figure 6. Histograms of the Probability Integral Transform (PIT) per Municipality by Cancer Site and Sex for Incidence and Mortality**

**Incidence, Male**

**Probability Integral Transform（PIT）**

**All Cancer**

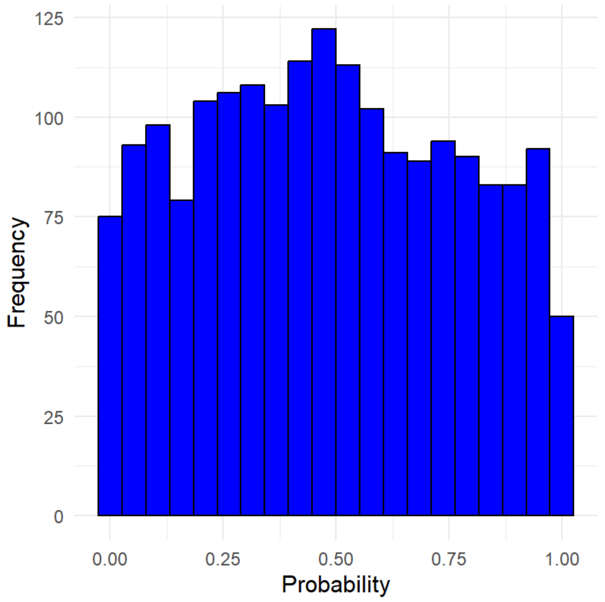

**Stomach**

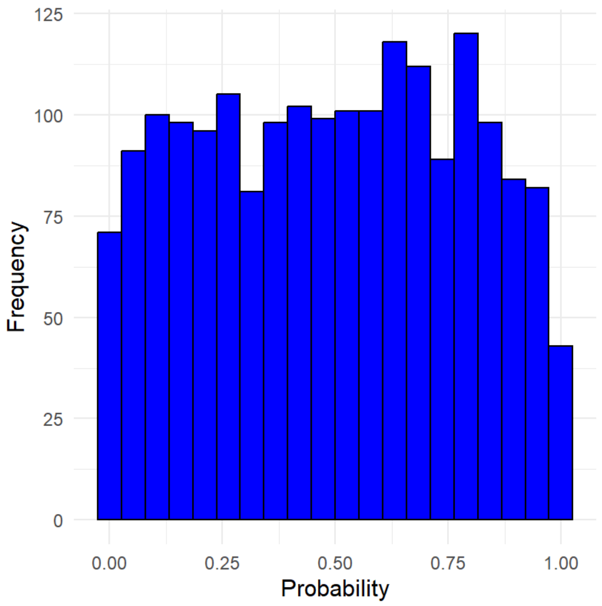

**Colorectum**

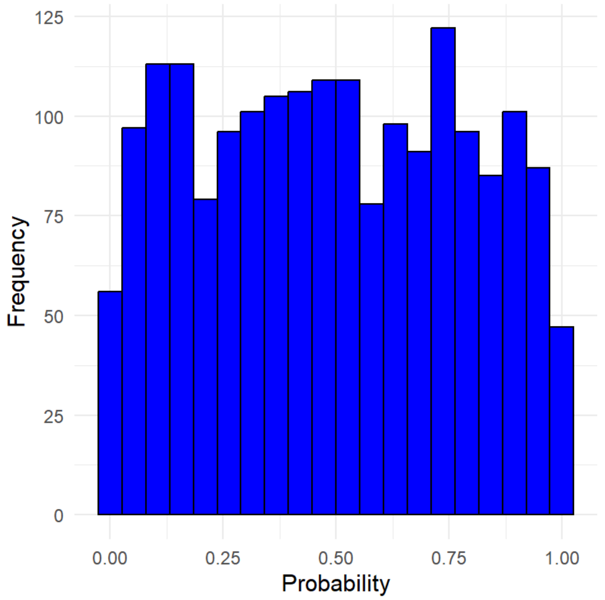

**Lung**

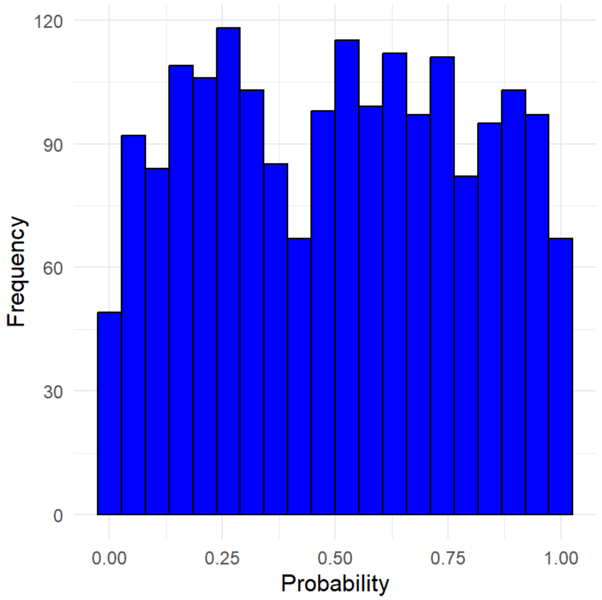

**Supplementary Figure 6. Histograms of the Probability Integral Transform (PIT) per Municipality by Cancer Site and Sex for Incidence and Mortality**

### Incidence, Male

### Probability Integral Transform（PIT）

#### Liver

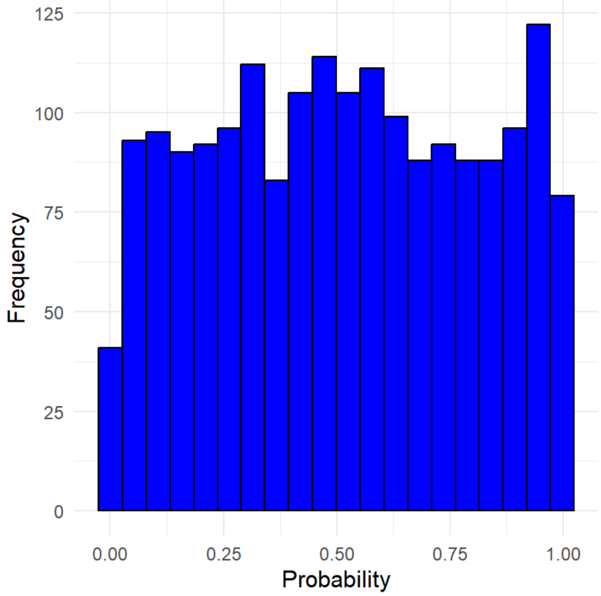

#### Pancreas

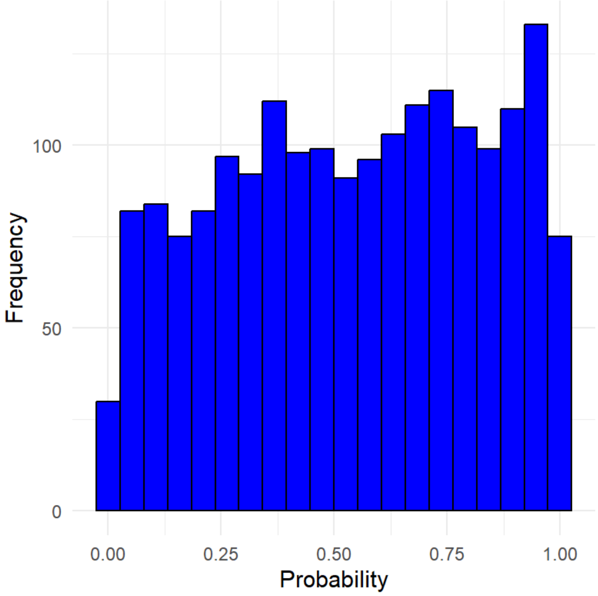

#### Prostate

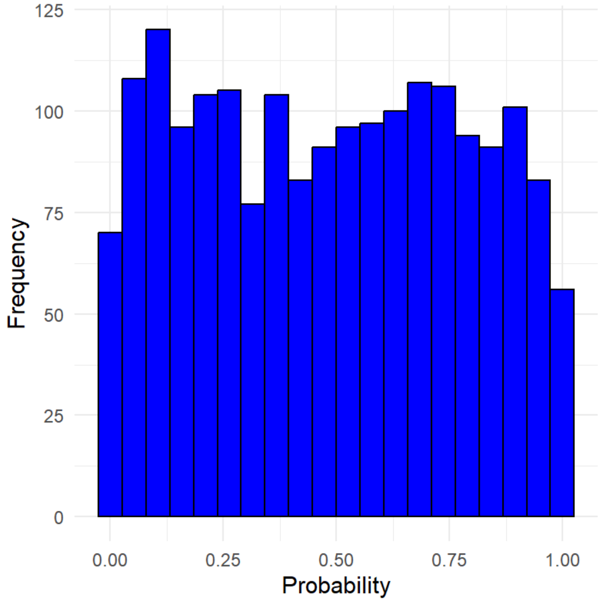

**Supplementary Figure 6. Histograms of the Probability Integral Transform (PIT) per Municipality by Cancer Site and Sex for Incidence and Mortality**

**Mortality, Male**

**Probability Integral Transform（PIT）**

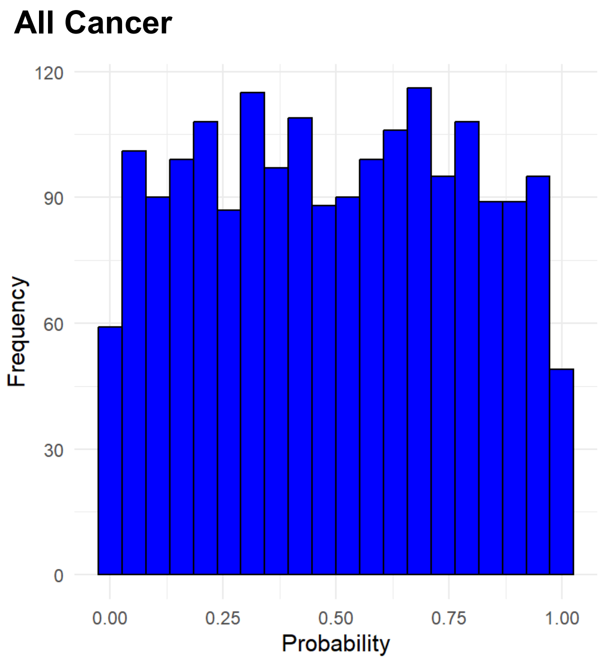

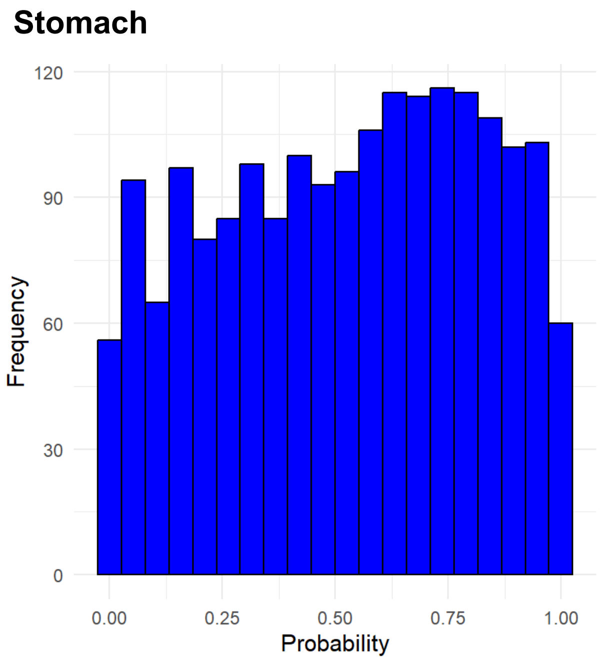

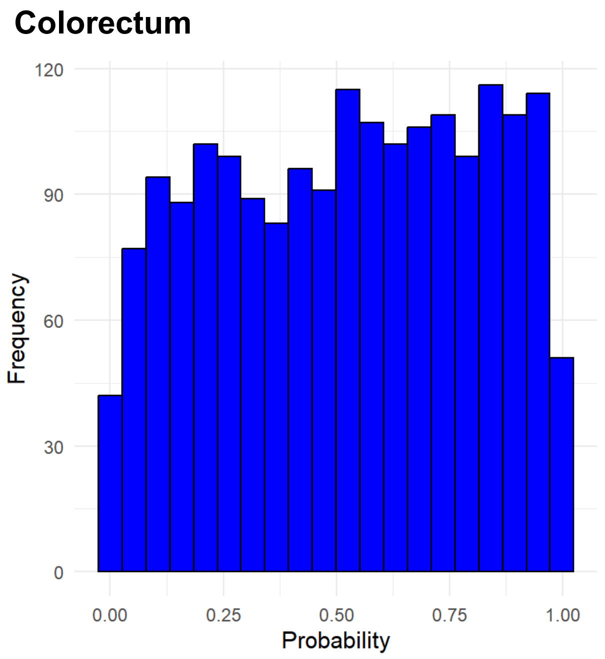

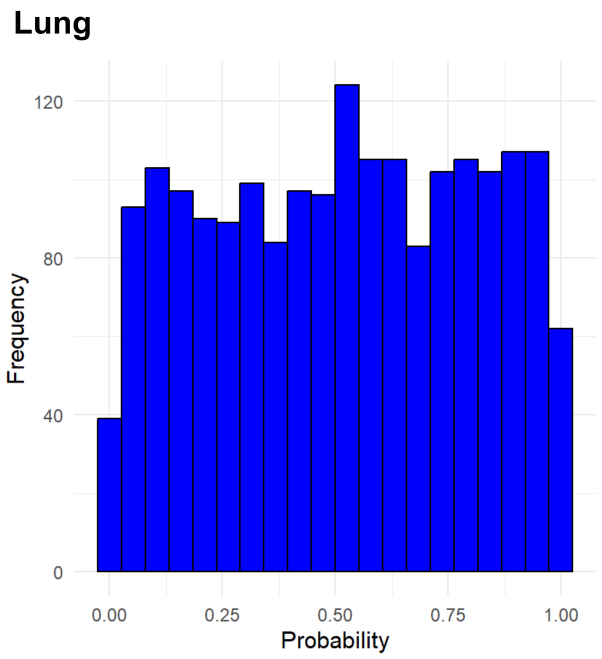

**Supplementary Figure 6. Histograms of the Probability Integral Transform (PIT) per Municipality by Cancer Site and Sex for Incidence and Mortality**

### Mortality, Male

### Probability Integral Transform（PIT）

#### Liver

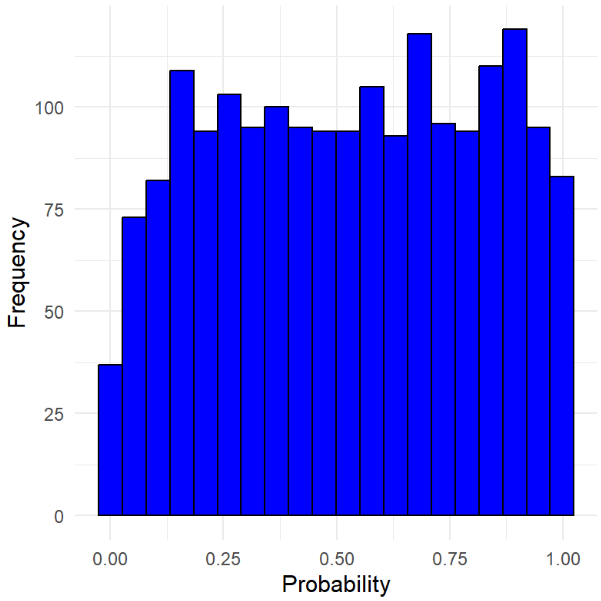

#### Pancreas

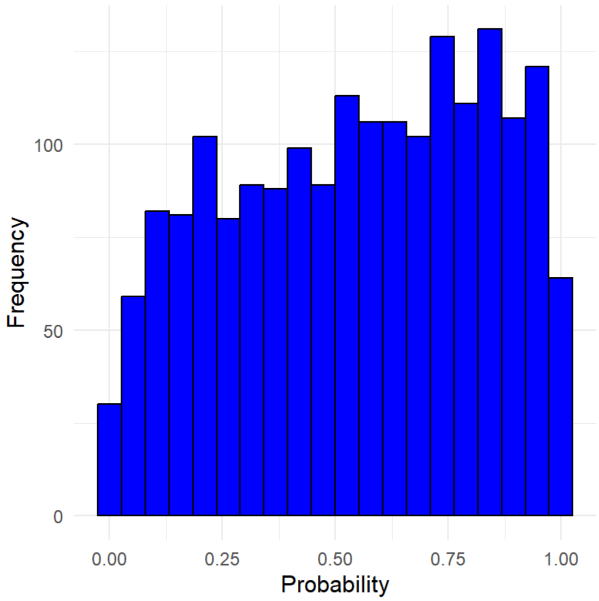

#### Prostate

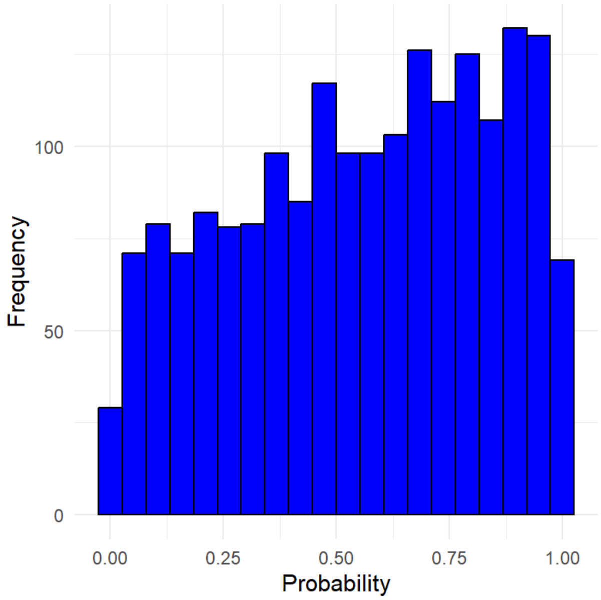

**Supplementary Figure 6. Histograms of the Probability Integral Transform (PIT) per Municipality by Cancer Site and Sex for Incidence and Mortality**

### Incidence, Female

### Probability Integral Transform（PIT）

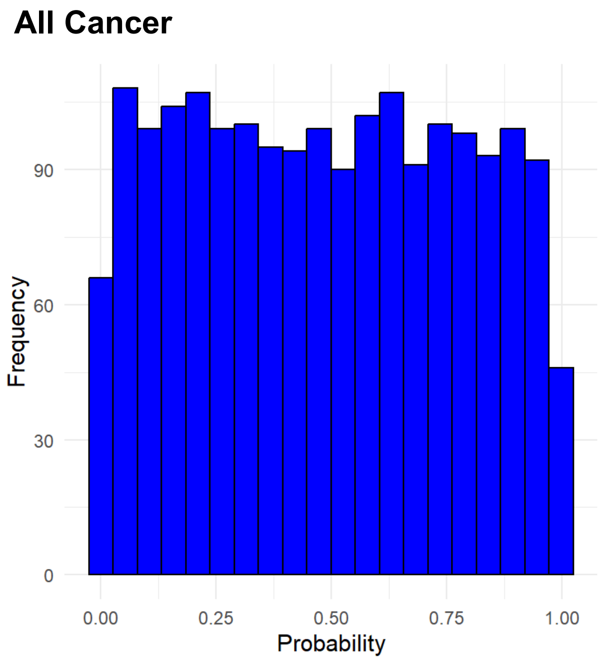

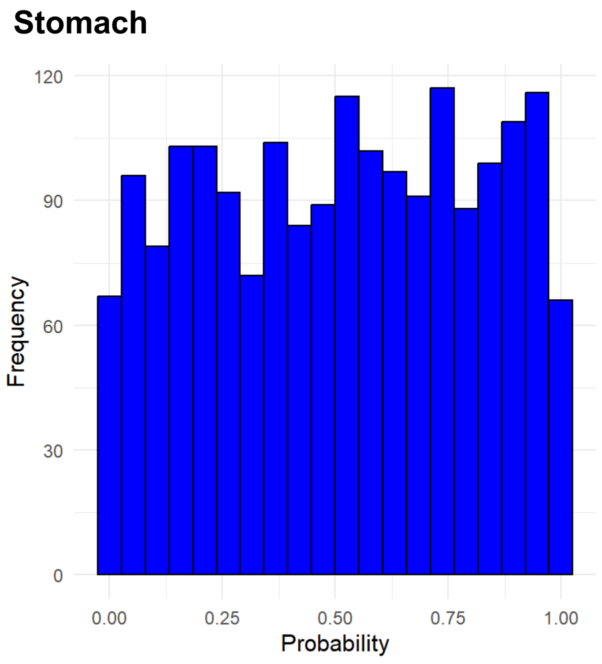

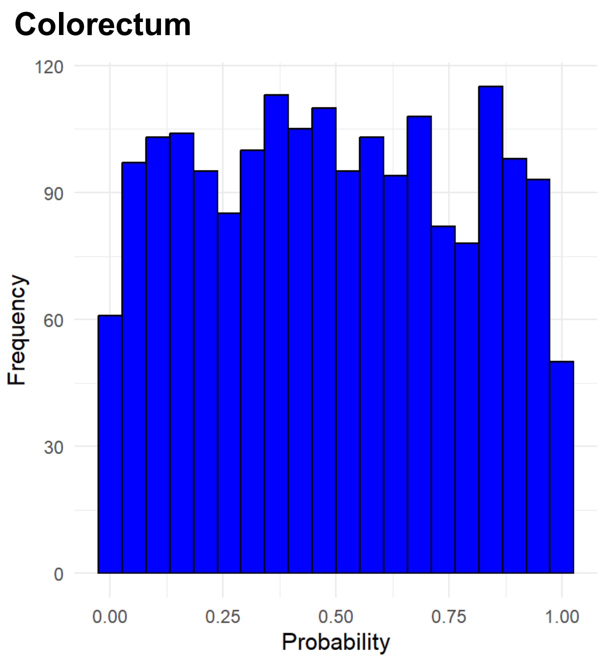

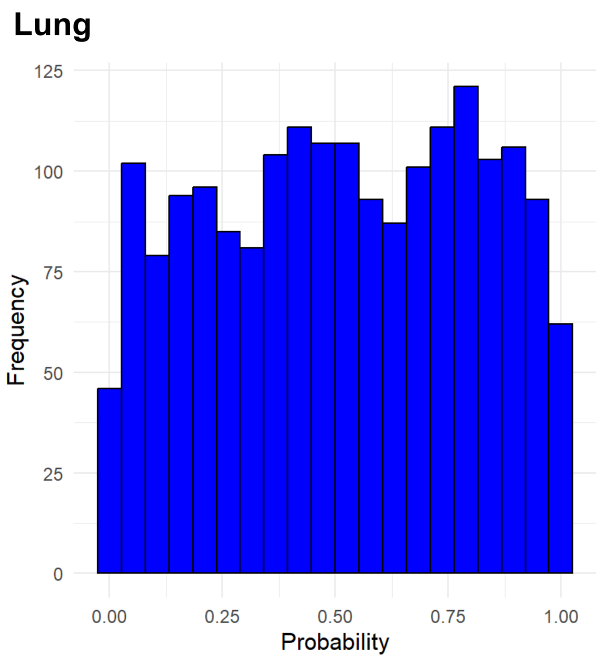

**Supplementary Figure 6. Histograms of the Probability Integral Transform (PIT) per Municipality by Cancer Site and Sex for Incidence and Mortality**

### Incidence, Female

### Probability Integral Transform（PIT）

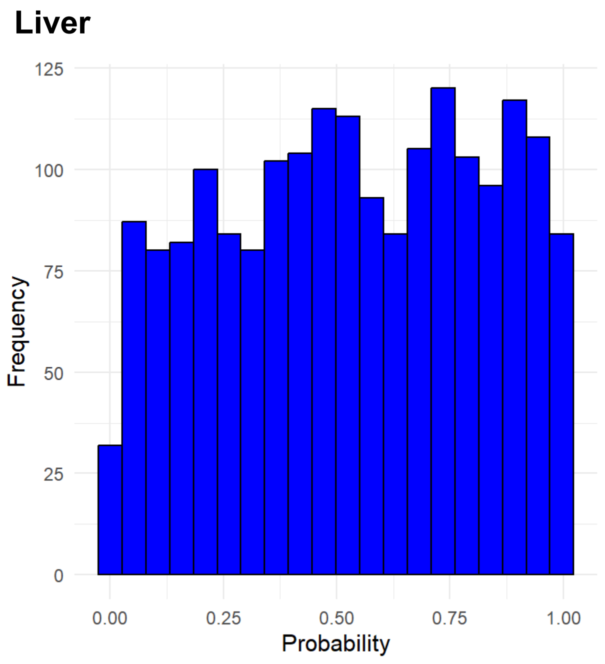

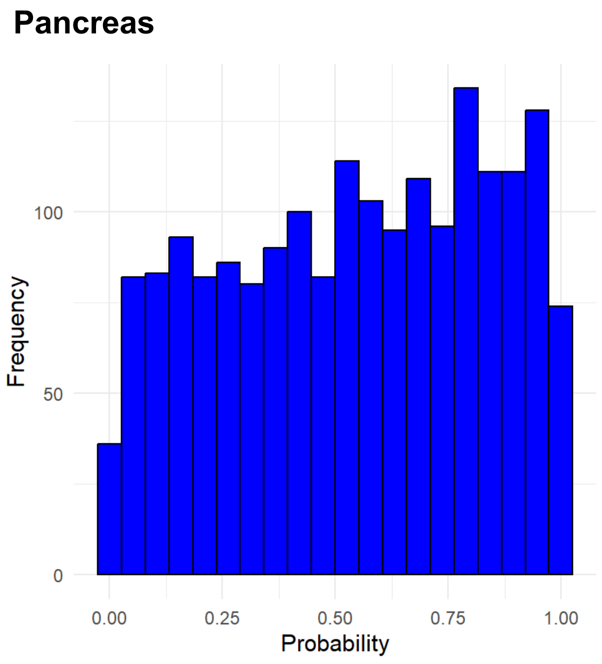

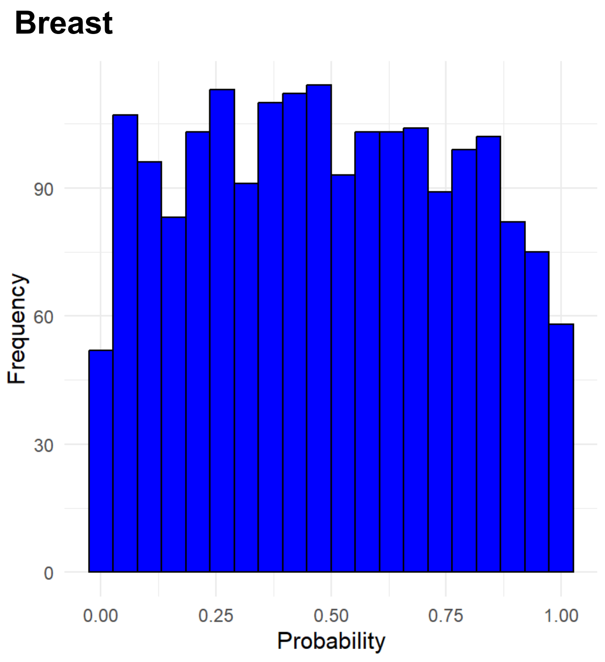

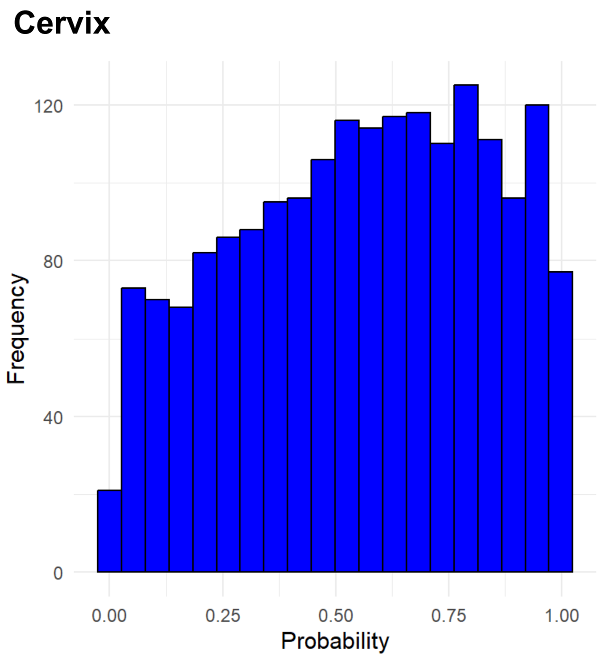

**Supplementary Figure 6. Histograms of the Probability Integral Transform (PIT) per Municipality by Cancer Site and Sex for Incidence and Mortality**

### Mortality, Female

### Probability Integral Transform（PIT）

#### All Cancer

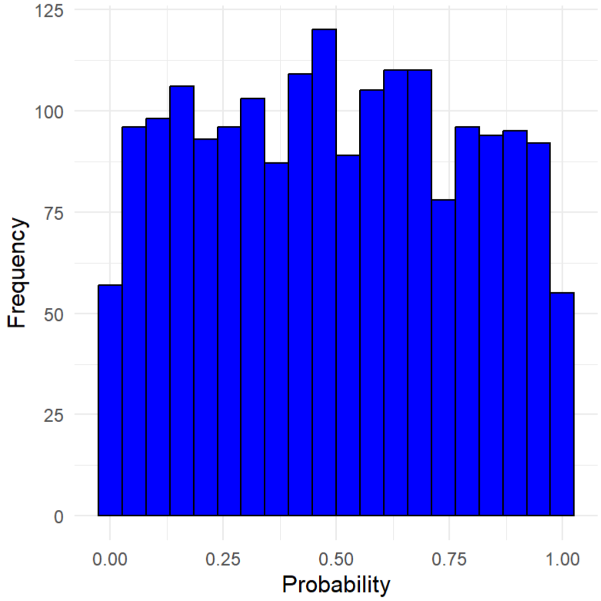

#### Stomach

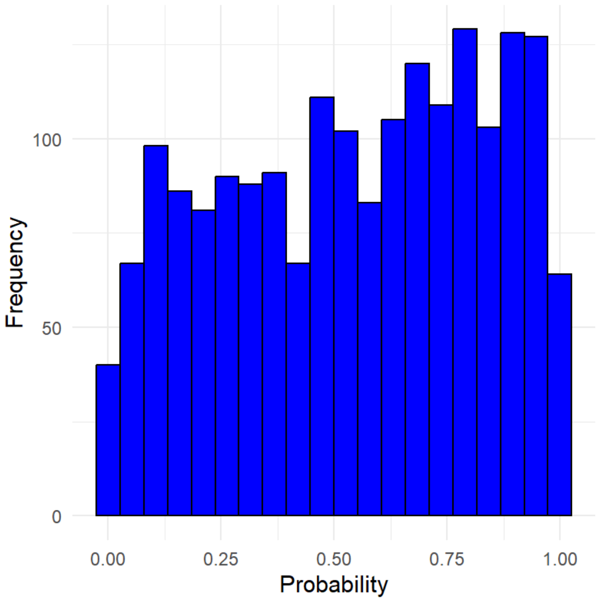

#### Colorectum

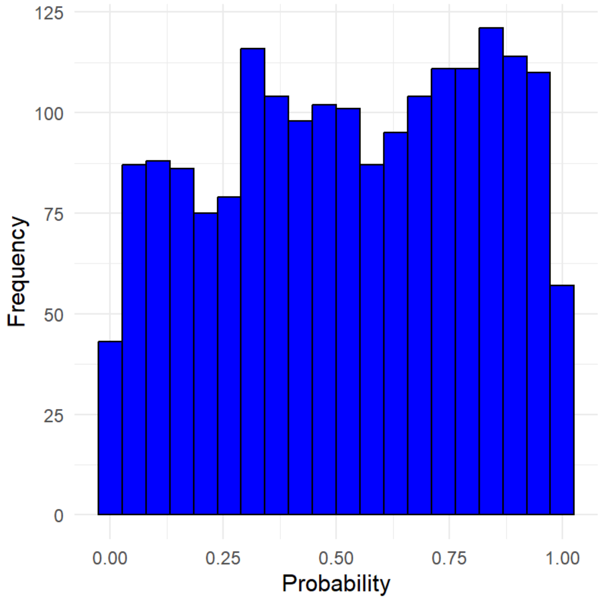

#### Lung

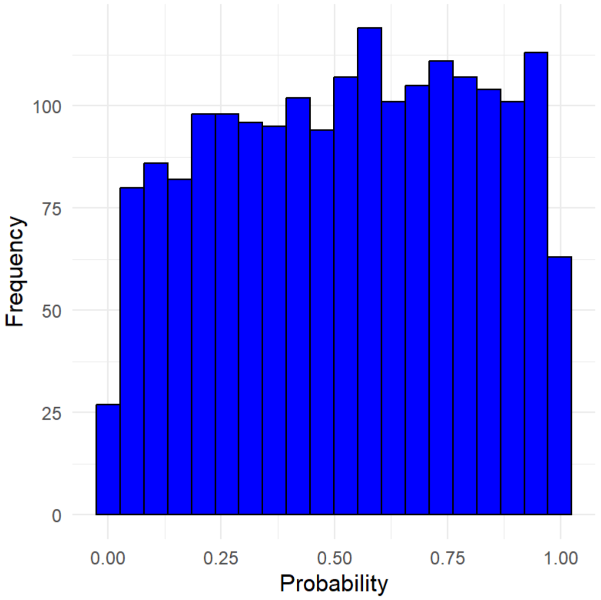

**Supplementary Figure 6. Histograms of the Probability Integral Transform (PIT) per Municipality by Cancer Site and Sex for Incidence and Mortality**

### Mortality, Female

### Probability Integral Transform（PIT）

#### Liver

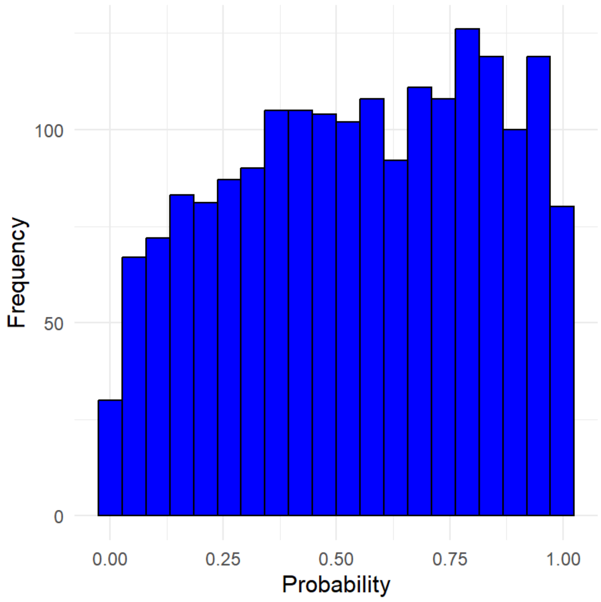

#### Pancreas

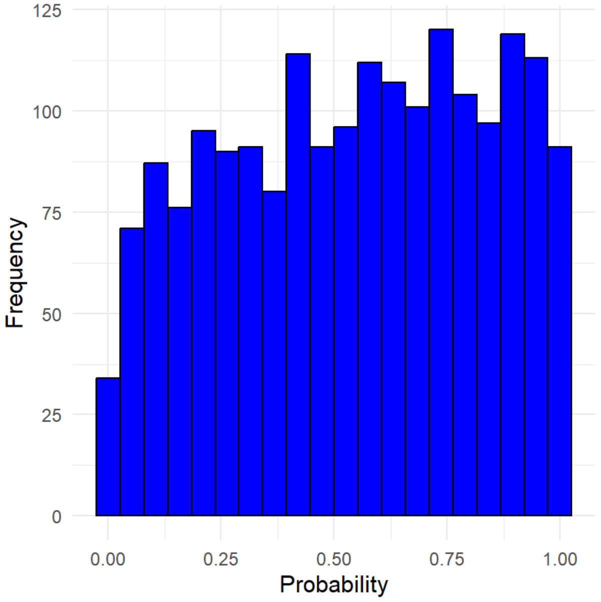

#### Breast

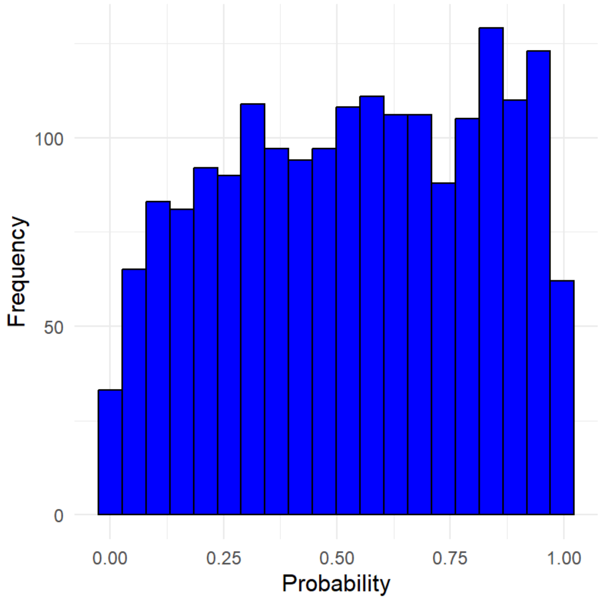

#### Cervix

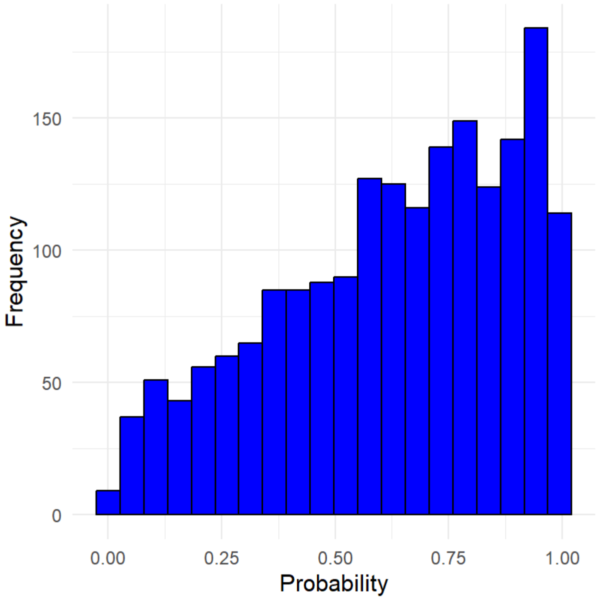

**Supplementary Figure 7. Posterior Predictive Check (PPC): Scatterplots of Posterior Predictions vs. Observed Counts and Histograms of Posterior Predictive P-Values per Municipality by Cancer Site and Sex for Incidence and Mortality**

| Incidence, Male | Predicted and observed counts | Posterior predictive p values |
|---|---|---|
| All Cancer | | |
| Stomach | | |
| Colorectum | | |
| Lung | | |

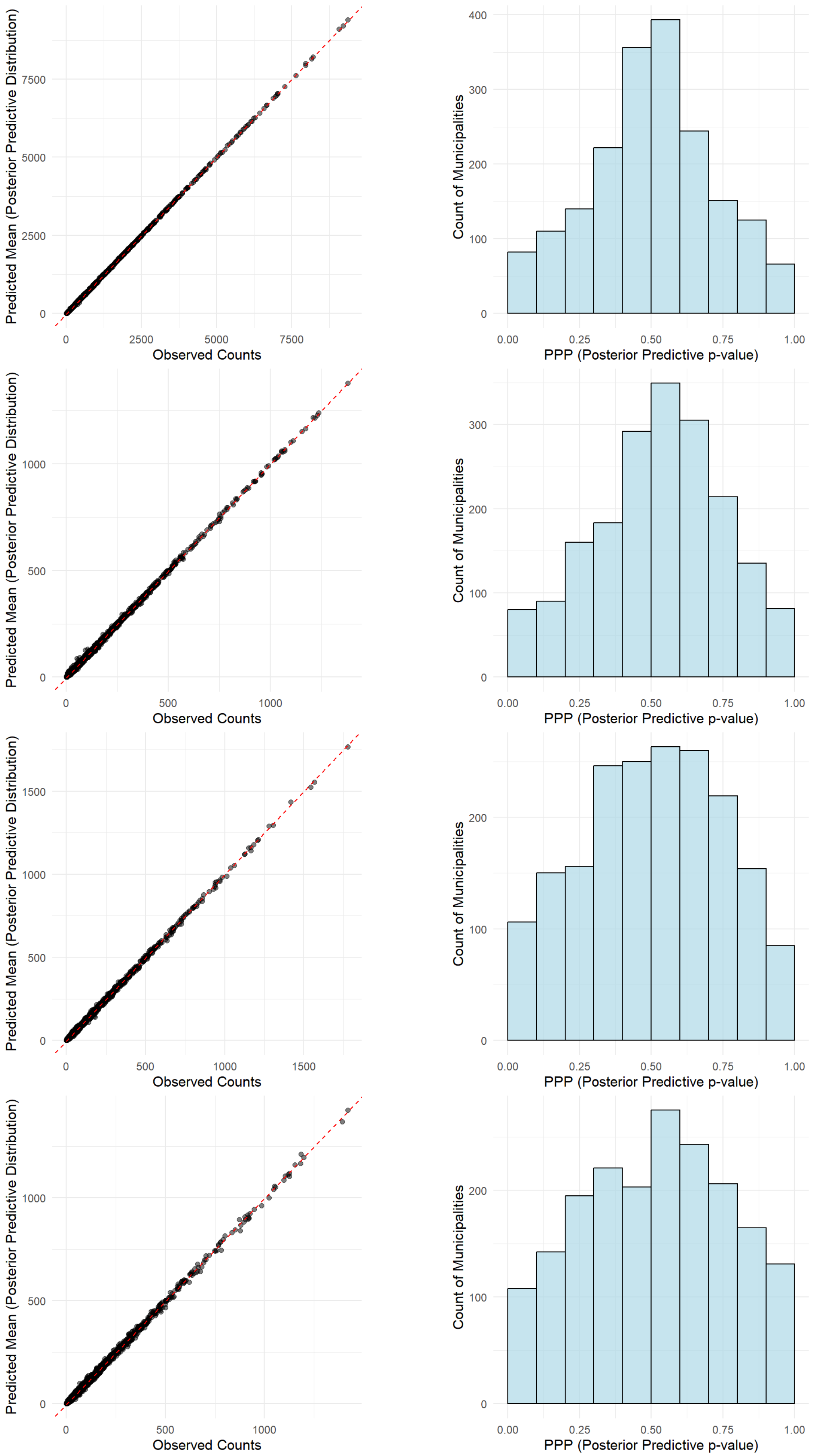

**Supplementary Figure 7. Posterior Predictive Check (PPC): Scatterplots of Posterior Predictions vs. Observed Counts and Histograms of Posterior Predictive P-Values per Municipality by Cancer Site and Sex for Incidence and Mortality**

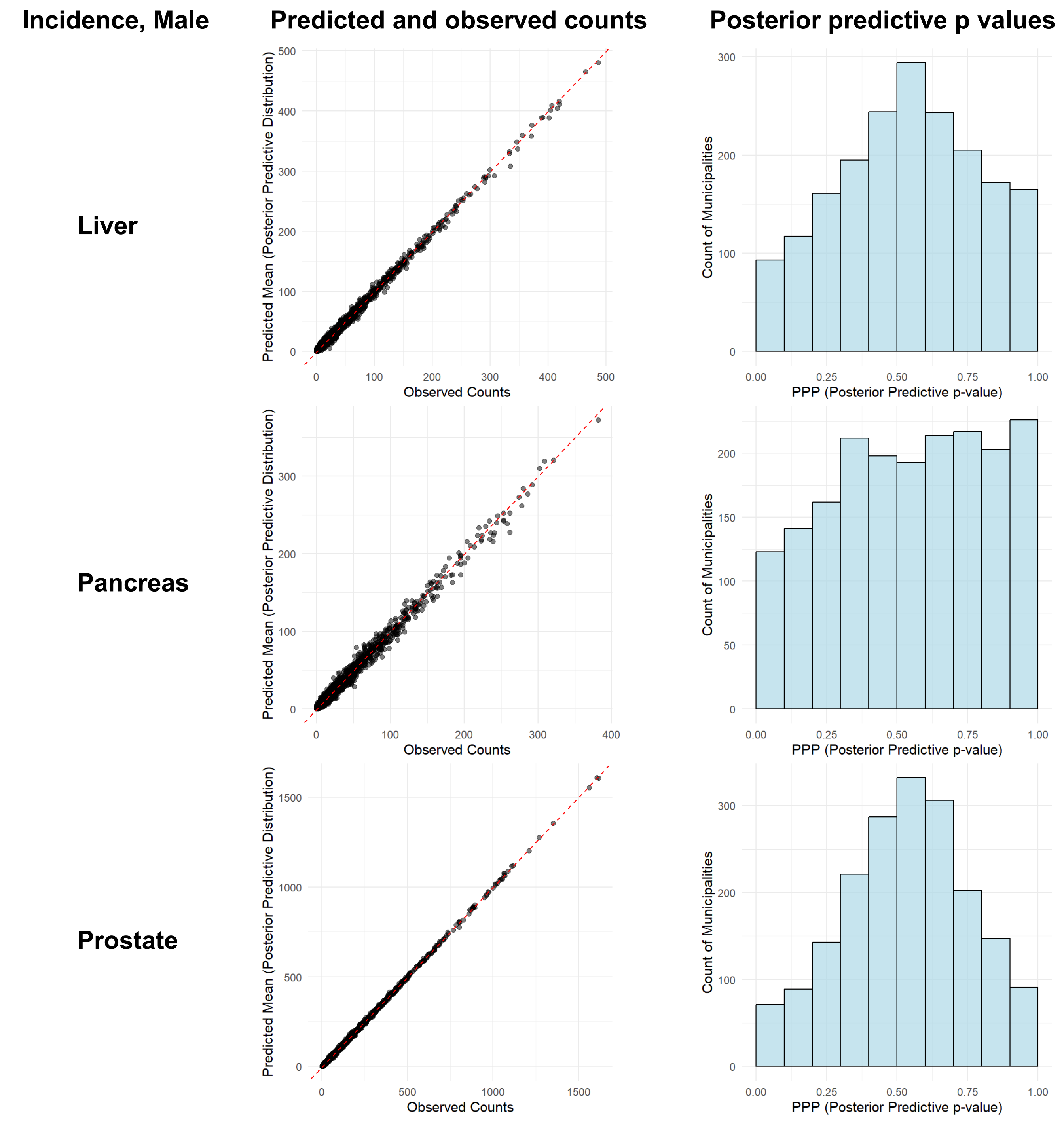

**Supplementary Figure 7. Posterior Predictive Check (PPC): Scatterplots of Posterior Predictions vs. Observed Counts and Histograms of Posterior Predictive P-Values per Municipality by Cancer Site and Sex for Incidence and Mortality**

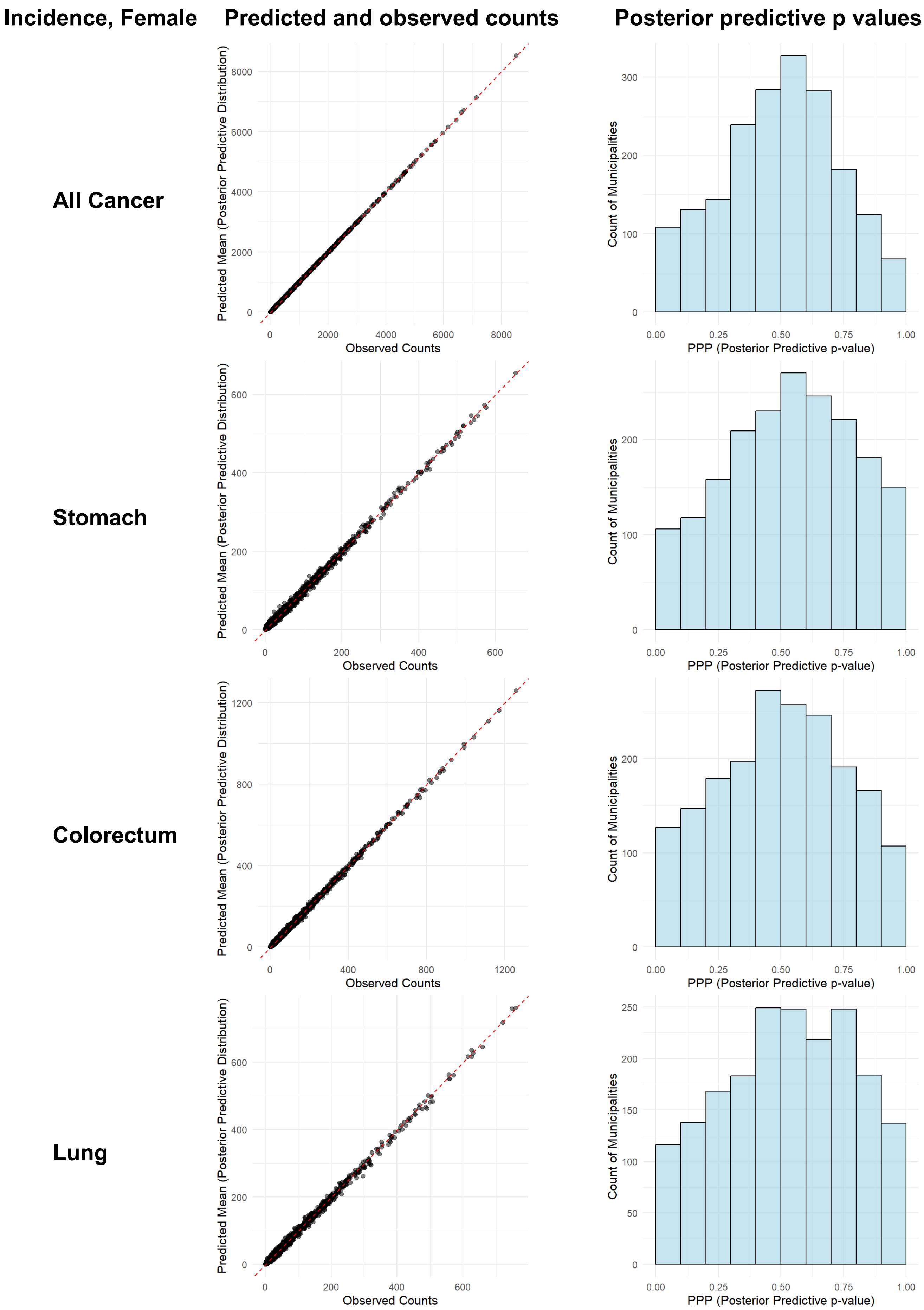

**Supplementary Figure 7. Posterior Predictive Check (PPC): Scatterplots of Posterior Predictions vs. Observed Counts and Histograms of Posterior Predictive P-Values per Municipality by Cancer Site and Sex for Incidence and Mortality**

| Incidence, Female | Predicted and observed counts | Posterior predictive p values |
|---|---|---|
| **Liver** | 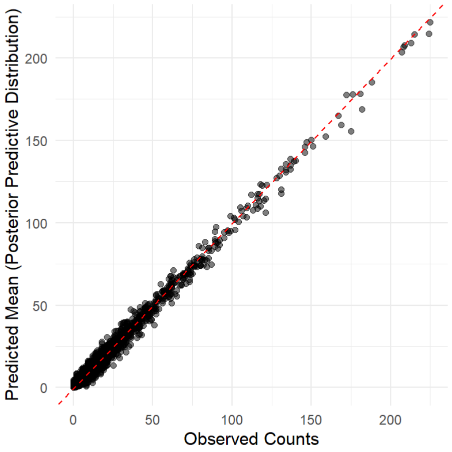 | 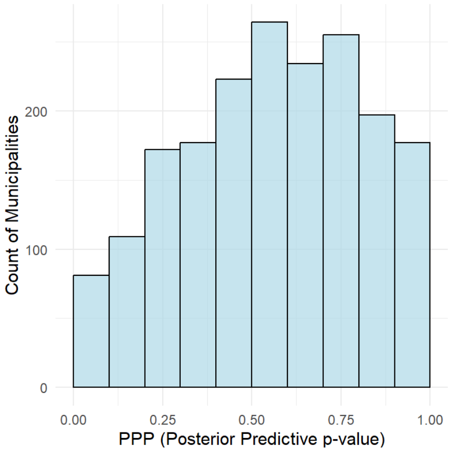 |
| **Pancreas** | 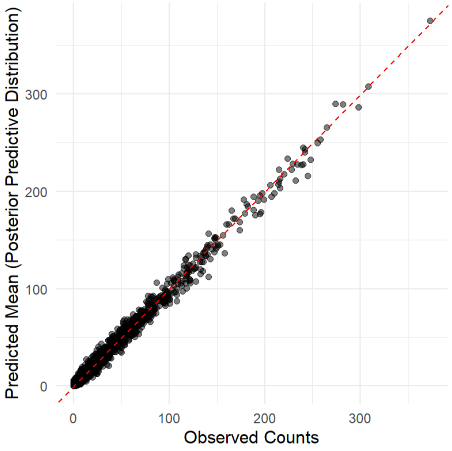 | 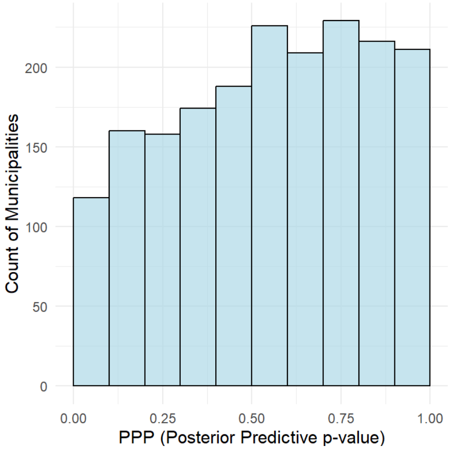 |
| **Breast** | 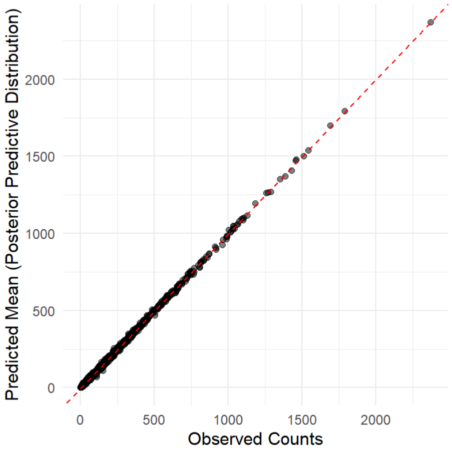 | 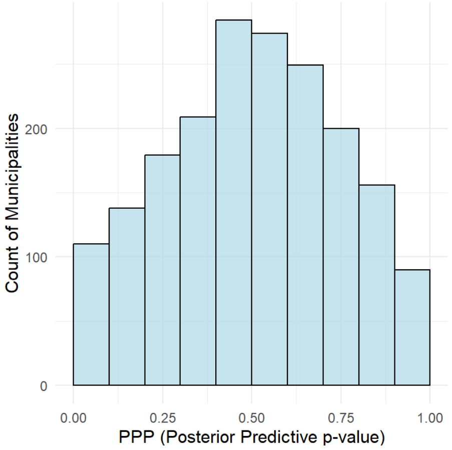 |
| **Cervix** | 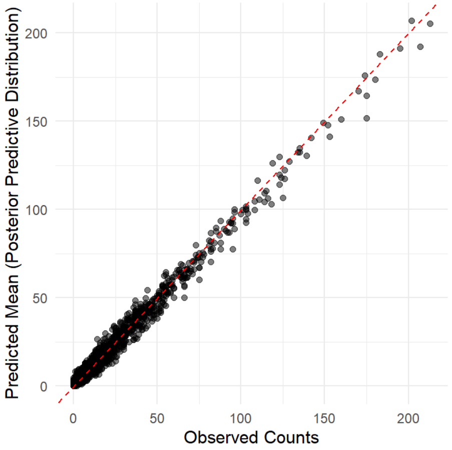 | 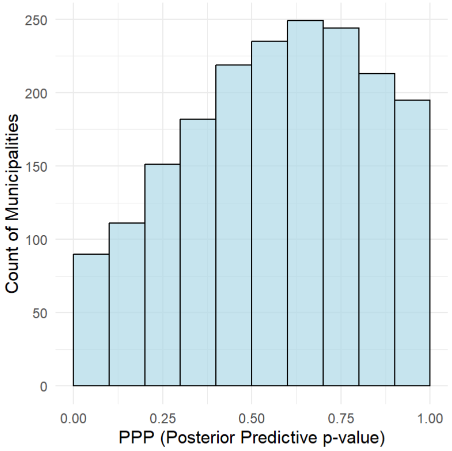 |

**Supplementary Figure 7. Posterior Predictive Check (PPC): Scatterplots of Posterior Predictions vs. Observed Counts and Histograms of Posterior Predictive P-Values per Municipality by Cancer Site and Sex for Incidence and Mortality**

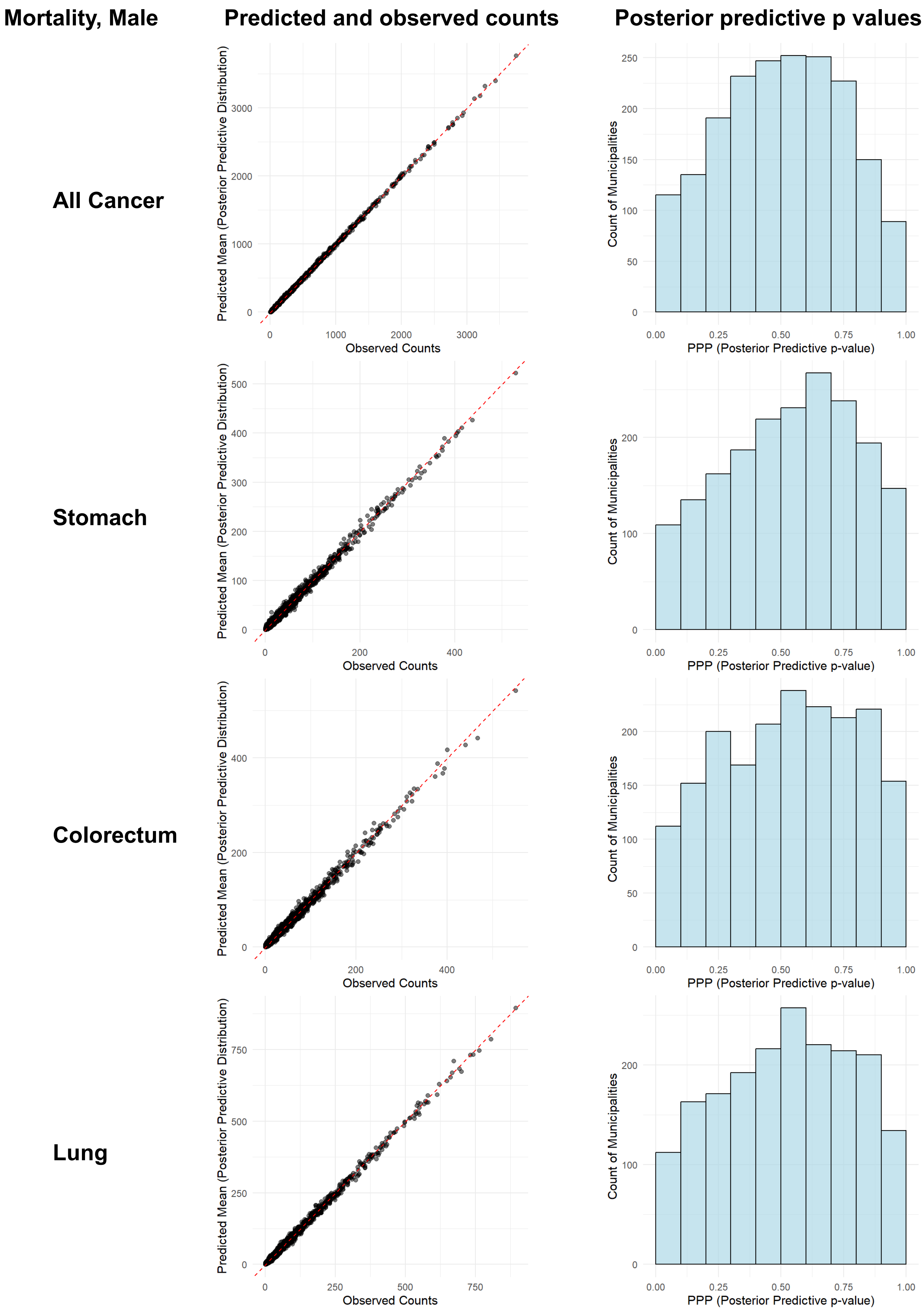

**Supplementary Figure 7. Posterior Predictive Check (PPC): Scatterplots of Posterior Predictions vs. Observed Counts and Histograms of Posterior Predictive P-Values per Municipality by Cancer Site and Sex for Incidence and Mortality**

| Mortality, Male | Predicted and observed counts | Posterior predictive p values |
|---|---|---|
| **Liver** | | |
| **Pancreas** | | |
| **Prostate** | | |

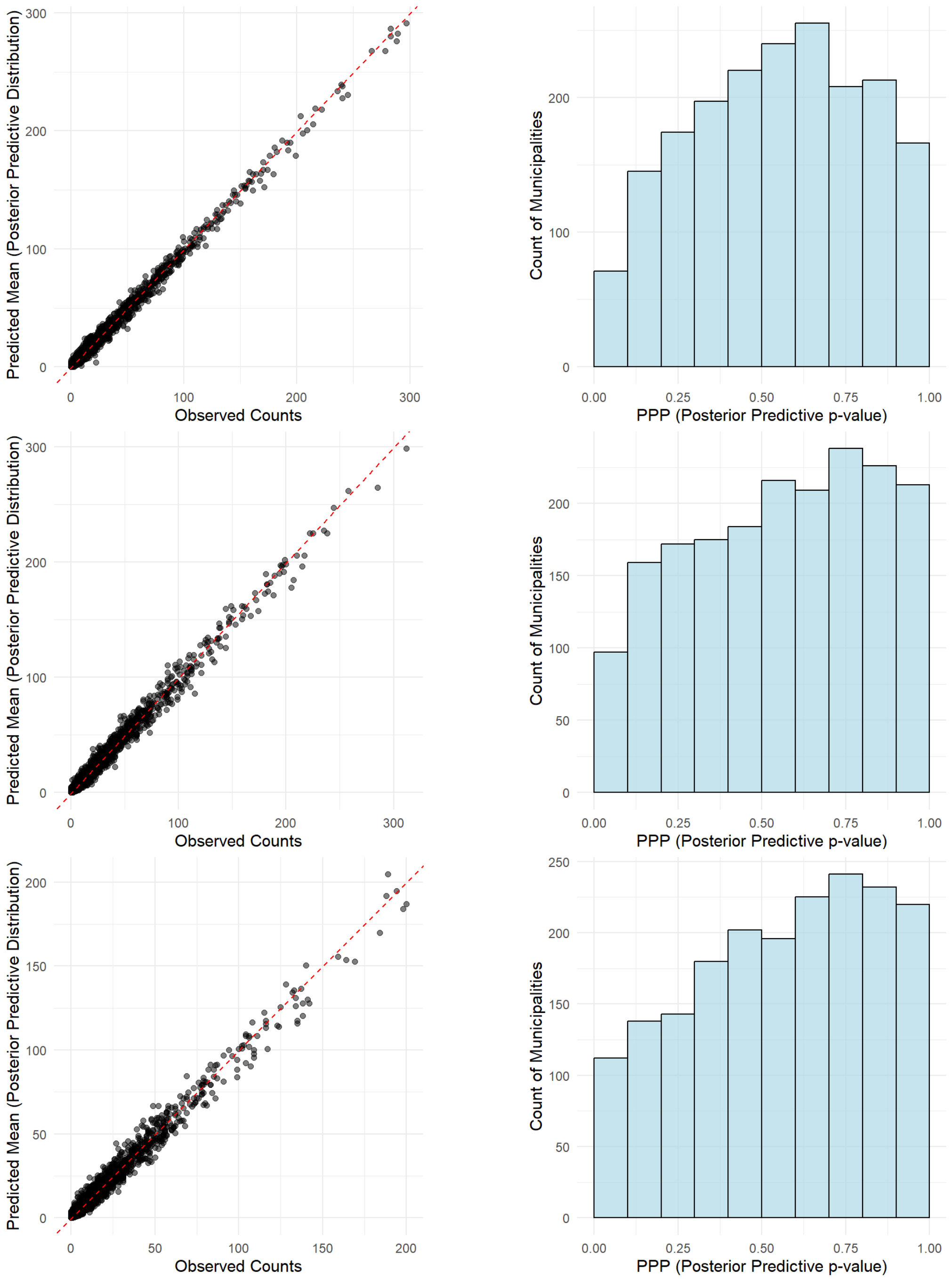

**Supplementary Figure 7. Posterior Predictive Check (PPC): Scatterplots of Posterior Predictions vs. Observed Counts and Histograms of Posterior Predictive P-Values per Municipality by Cancer Site and Sex for Incidence and Mortality**

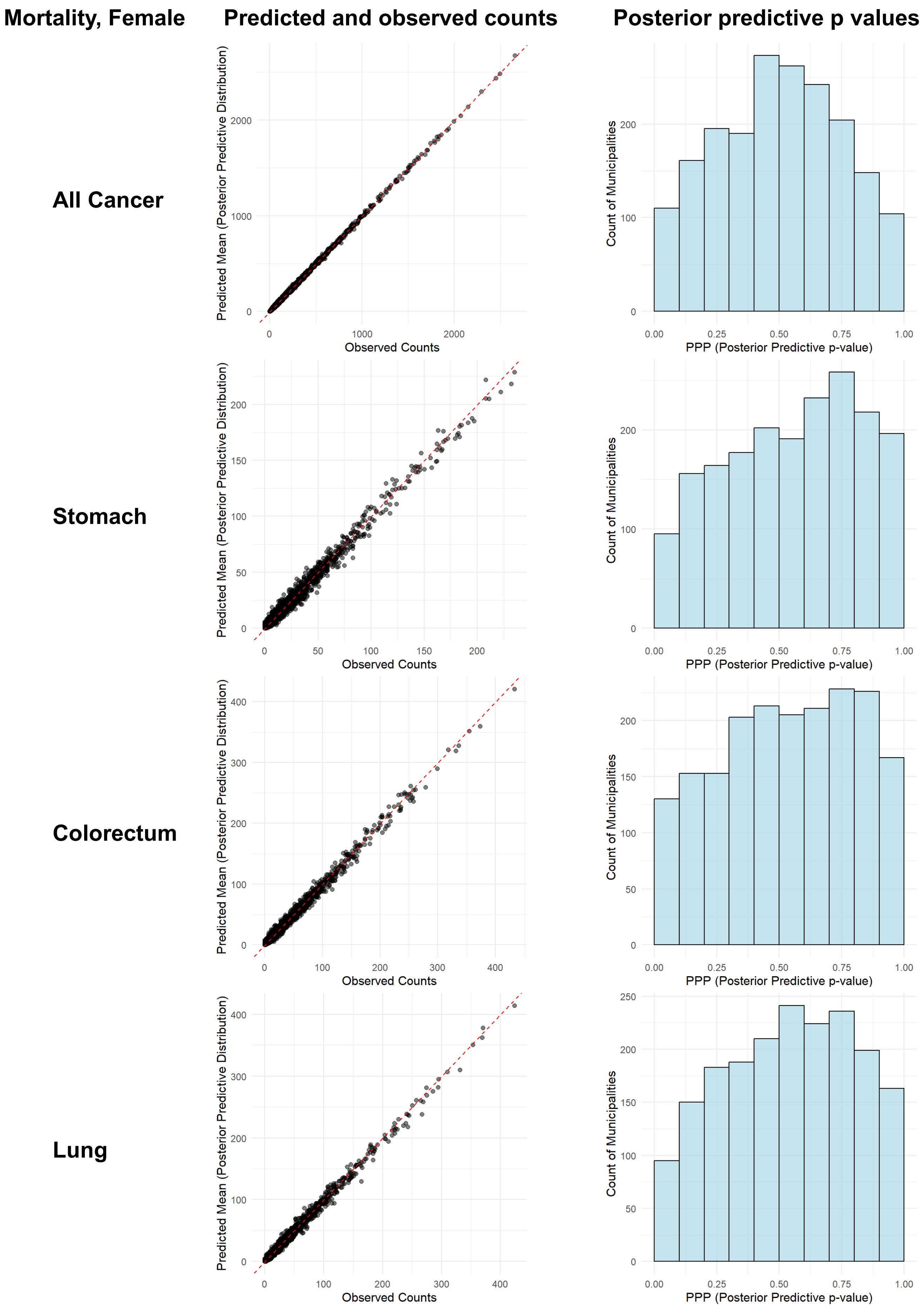

**Supplementary Figure 7. Posterior Predictive Check (PPC): Scatterplots of Posterior Predictions vs. Observed Counts and Histograms of Posterior Predictive P-Values per Municipality by Cancer Site and Sex for Incidence and Mortality**

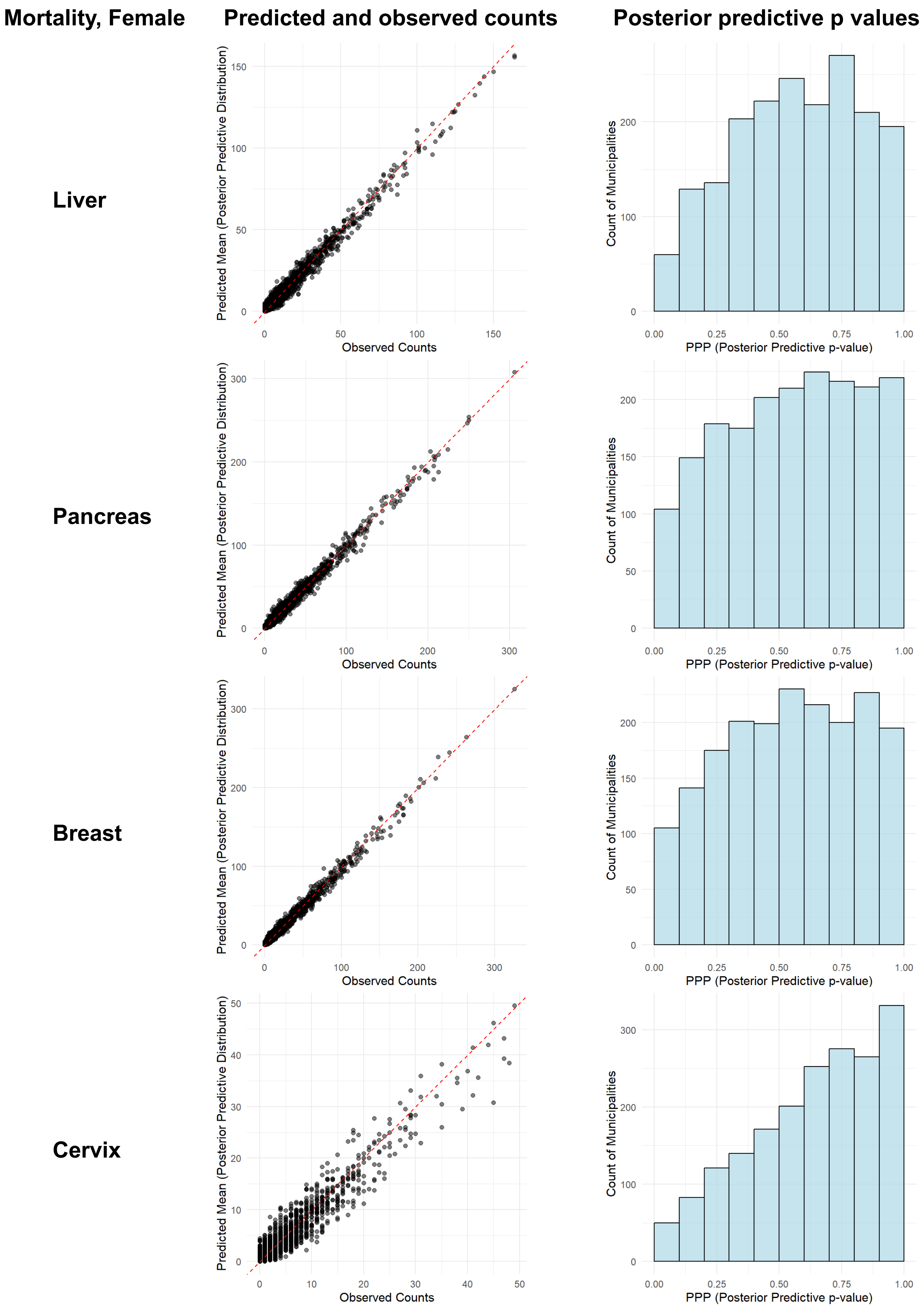

**Supplementary Figure 8. Impact of normADI (based on 2010 Census) on FBSIR/FBSMR by Sex and Urbanicity**
Forest plots illustrating the risk ratio (RR) and its 95% credible interval (CR) of FBSIR/FBSMR for the hypothetical worst ADI (normADI = 1) compared to the hypothetical best ADI (normADI = 0) for males (A) and females (B). Evidence of a difference was considered when the 95% CR of the RR did not include 1. Heterogeneity estimates marked with a single asterisk indicated notable variation. ADI, Areal Deprivation Index; FBSIR, fully Bayesian estimate of standardized incidence ratio; FBSMR, fully Bayesian estimate of standardized mortality ratio; Het, heterogeneity through levels of urbanicity (suburban vs. urban; rural vs. urban); normADI, normalized ADI; inc, incidence; mor, mortality.

### (A) Male

| Site | Urbanicity | RR_inc (95% CR) | Het RR_inc (95% CR) | | RR_mor (95% CR) | Het RR_mor (95% CR) | |
|---|---|---|---|---|---|---|---|
| **All** | All | 1.00 (0.99 to 1.02) | | | 1.11 (1.09 to 1.13) | | |
| | Urban | 1.04 (1.00 to 1.08) | | | 1.20 (1.16 to 1.25) | | |
| | Suburban | 1.02 (1.00 to 1.05) | 0.98 (0.94 to 1.03) | | 1.11 (1.08 to 1.14) | 0.92 (0.88 to 0.96) | * |
| | Rural | 0.99 (0.96 to 1.02) | 0.95 (0.91 to 1.00) | * | 1.08 (1.05 to 1.12) | 0.90 (0.85 to 0.95) | * |
| **Stomach** | All | 0.83 (0.80 to 0.86) | | | 0.99 (0.94 to 1.03) | | |
| | Urban | 0.99 (0.91 to 1.06) | | | 1.23 (1.13 to 1.33) | | |
| | Suburban | 0.90 (0.86 to 0.95) | 0.91 (0.84 to 1.00) | * | 1.00 (0.94 to 1.07) | 0.82 (0.74 to 0.90) | * |
| | Rural | 0.71 (0.67 to 0.76) | 0.72 (0.65 to 0.80) | * | 0.83 (0.77 to 0.90) | 0.68 (0.60 to 0.76) | * |
| **Colorectum** | All | 1.06 (1.03 to 1.10) | | | 1.19 (1.14 to 1.24) | | |
| | Urban | 1.16 (1.09 to 1.23) | | | 1.31 (1.22 to 1.41) | | |
| | Suburban | 1.05 (1.01 to 1.10) | 0.91 (0.85 to 0.98) | * | 1.19 (1.12 to 1.26) | 0.90 (0.83 to 0.99) | * |
| | Rural | 1.03 (0.97 to 1.09) | 0.89 (0.82 to 0.97) | * | 1.12 (1.03 to 1.21) | 0.85 (0.76 to 0.95) | * |
| **Lung** | All | 1.16 (1.13 to 1.20) | | | 1.19 (1.15 to 1.23) | | |
| | Urban | 1.23 (1.16 to 1.30) | | | 1.28 (1.20 to 1.36) | | |
| | Suburban | 1.16 (1.11 to 1.21) | 0.94 (0.89 to 1.01) | | 1.18 (1.13 to 1.24) | 0.92 (0.86 to 0.99) | * |
| | Rural | 1.12 (1.06 to 1.19) | 0.91 (0.84 to 0.99) | * | 1.15 (1.08 to 1.22) | 0.90 (0.82 to 0.98) | * |
| **Liver** | All | 1.37 (1.30 to 1.44) | | | 1.33 (1.26 to 1.41) | | |
| | Urban | 1.37 (1.25 to 1.52) | | | 1.48 (1.32 to 1.65) | | |
| | Suburban | 1.48 (1.38 to 1.59) | 1.08 (0.96 to 1.21) | | 1.44 (1.33 to 1.57) | 0.98 (0.86 to 1.11) | |
| | Rural | 1.35 (1.23 to 1.48) | 0.98 (0.85 to 1.13) | | 1.24 (1.11 to 1.38) | 0.84 (0.72 to 0.98) | * |
| **Pancreas** | All | 0.98 (0.94 to 1.02) | | | 1.01 (0.96 to 1.05) | | |
| | Urban | 0.99 (0.92 to 1.06) | | | 1.04 (0.96 to 1.12) | | |
| | Suburban | 0.95 (0.89 to 1.01) | 0.96 (0.88 to 1.05) | | 1.00 (0.94 to 1.06) | 0.96 (0.88 to 1.05) | |
| | Rural | 1.04 (0.96 to 1.12) | 1.05 (0.95 to 1.17) | | 1.00 (0.92 to 1.08) | 0.96 (0.86 to 1.08) | |
| **Prostate** | All | 0.89 (0.85 to 0.92) | | | 1.09 (1.04 to 1.15) | | |
| | Urban | 0.83 (0.77 to 0.91) | | | 1.18 (1.08 to 1.29) | | |
| | Suburban | 0.89 (0.84 to 0.94) | 1.07 (0.97 to 1.18) | | 1.01 (0.94 to 1.09) | 0.86 (0.77 to 0.95) | * |
| | Rural | 0.94 (0.88 to 1.01) | 1.13 (1.01 to 1.26) | * | 1.19 (1.08 to 1.31) | 1.01 (0.88 to 1.15) | |

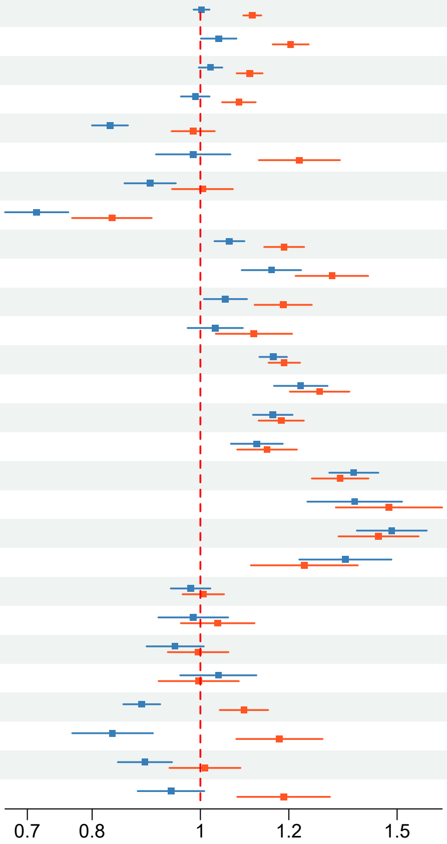

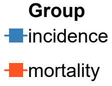

**Supplementary Figure 8. Impact of normADI (based on 2010 Census) on FBSIR/FBSMR by Sex and Urbanicity**
Forest plots illustrating the risk ratio (RR) and its 95% credible interval (CR) of FBSIR/FBSMR for the hypothetical worst ADI (normADI = 1) compared to the hypothetical best ADI (normADI = 0) for males (A) and females (B). Evidence of a difference was considered when the 95% CR of the RR did not include 1. Heterogeneity estimates marked with a single asterisk indicated notable variation. ADI, Areal Deprivation Index; FBSIR, fully Bayesian estimate of standardized incidence ratio; FBSMR, fully Bayesian estimate of standardized mortality ratio; Het, heterogeneity through levels of urbanicity (suburban vs. urban; rural vs. urban); normADI, normalized ADI; inc, incidence; mor, mortality.

### (B) Female

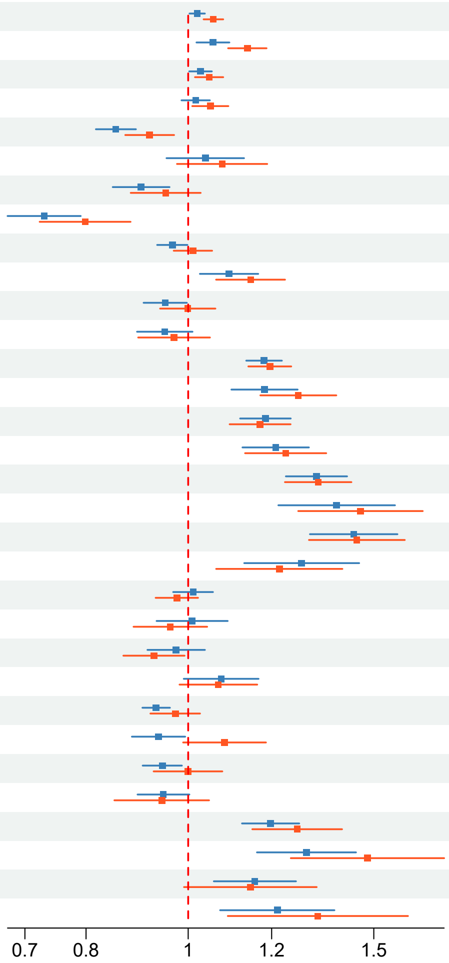

| Site | Urbanicity | RR_inc (95% CR) | Het RR_inc (95% CR) | | RR_mor (95% CR) | Het RR_mor (95% CR) | |
|---|---|---|---|---|---|---|---|
| **All** | All | 1.02 (1.00 to 1.04) | | | 1.06 (1.03 to 1.08) | | |
| | Urban | 1.06 (1.02 to 1.09) | | | 1.14 (1.09 to 1.19) | | |
| | Suburban | 1.03 (1.00 to 1.05) | 0.97 (0.93 to 1.01) | | 1.05 (1.01 to 1.08) | 0.92 (0.88 to 0.97) | * |
| | Rural | 1.02 (0.99 to 1.05) | 0.96 (0.92 to 1.01) | | 1.05 (1.01 to 1.09) | 0.92 (0.87 to 0.98) | * |
| **Stomach** | All | 0.85 (0.82 to 0.89) | | | 0.92 (0.87 to 0.97) | | |
| | Urban | 1.04 (0.95 to 1.13) | | | 1.08 (0.98 to 1.19) | | |
| | Suburban | 0.90 (0.85 to 0.96) | 0.87 (0.79 to 0.96) | * | 0.95 (0.88 to 1.03) | 0.88 (0.79 to 0.99) | * |
| | Rural | 0.73 (0.67 to 0.79) | 0.70 (0.63 to 0.79) | * | 0.80 (0.72 to 0.88) | 0.74 (0.64 to 0.85) | * |
| **Colorectum** | All | 0.97 (0.93 to 1.00) | | | 1.01 (0.97 to 1.05) | | |
| | Urban | 1.09 (1.03 to 1.17) | | | 1.15 (1.06 to 1.24) | | |
| | Suburban | 0.95 (0.91 to 1.00) | 0.87 (0.81 to 0.94) | * | 1.00 (0.94 to 1.06) | 0.87 (0.80 to 0.95) | * |
| | Rural | 0.95 (0.89 to 1.01) | 0.87 (0.80 to 0.95) | * | 0.97 (0.90 to 1.05) | 0.85 (0.76 to 0.94) | * |
| **Lung** | All | 1.18 (1.14 to 1.23) | | | 1.20 (1.14 to 1.25) | | |
| | Urban | 1.18 (1.10 to 1.27) | | | 1.27 (1.17 to 1.38) | | |
| | Suburban | 1.18 (1.12 to 1.25) | 1.00 (0.92 to 1.09) | | 1.17 (1.09 to 1.25) | 0.92 (0.83 to 1.02) | |
| | Rural | 1.21 (1.13 to 1.30) | 1.02 (0.92 to 1.14) | | 1.24 (1.13 to 1.35) | 0.97 (0.86 to 1.10) | |
| **Liver** | All | 1.32 (1.24 to 1.41) | | | 1.33 (1.24 to 1.43) | | |
| | Urban | 1.38 (1.22 to 1.57) | | | 1.46 (1.27 to 1.67) | | |
| | Suburban | 1.44 (1.30 to 1.58) | 1.04 (0.89 to 1.20) | | 1.45 (1.30 to 1.61) | 0.99 (0.85 to 1.16) | |
| | Rural | 1.28 (1.13 to 1.45) | 0.93 (0.77 to 1.11) | | 1.22 (1.06 to 1.40) | 0.84 (0.69 to 1.02) | |
| **Pancreas** | All | 1.01 (0.97 to 1.06) | | | 0.98 (0.93 to 1.02) | | |
| | Urban | 1.01 (0.93 to 1.09) | | | 0.96 (0.89 to 1.04) | | |
| | Suburban | 0.97 (0.91 to 1.04) | 0.97 (0.88 to 1.06) | | 0.93 (0.87 to 0.99) | 0.96 (0.87 to 1.07) | |
| | Rural | 1.07 (0.99 to 1.17) | 1.07 (0.95 to 1.19) | | 1.07 (0.98 to 1.16) | 1.11 (0.99 to 1.25) | |
| **Breast** | All | 0.93 (0.90 to 0.96) | | | 0.97 (0.92 to 1.03) | | |
| | Urban | 0.94 (0.88 to 0.99) | | | 1.08 (0.99 to 1.19) | | |
| | Suburban | 0.94 (0.91 to 0.99) | 1.01 (0.94 to 1.08) | | 1.00 (0.93 to 1.08) | 0.92 (0.83 to 1.03) | |
| | Rural | 0.95 (0.90 to 1.00) | 1.01 (0.93 to 1.10) | | 0.94 (0.85 to 1.05) | 0.87 (0.76 to 1.00) | |
| **Cervix** | All | 1.20 (1.12 to 1.27) | | | 1.27 (1.15 to 1.40) | | |
| | Urban | 1.29 (1.16 to 1.44) | | | 1.48 (1.25 to 1.75) | | |
| | Suburban | 1.16 (1.06 to 1.27) | 0.89 (0.78 to 1.02) | | 1.15 (0.99 to 1.32) | 0.77 (0.63 to 0.95) | * |
| | Rural | 1.22 (1.07 to 1.38) | 0.94 (0.79 to 1.11) | | 1.33 (1.09 to 1.62) | 0.90 (0.69 to 1.16) | |

**Supplementary Figure 9. Distribution of the Prevalence of BMI≥25, Diabetes, Dyslipidemia, Insufficient Exercise, Smoking, and Drinking Across the Three Urbanicity Categories by Sex**

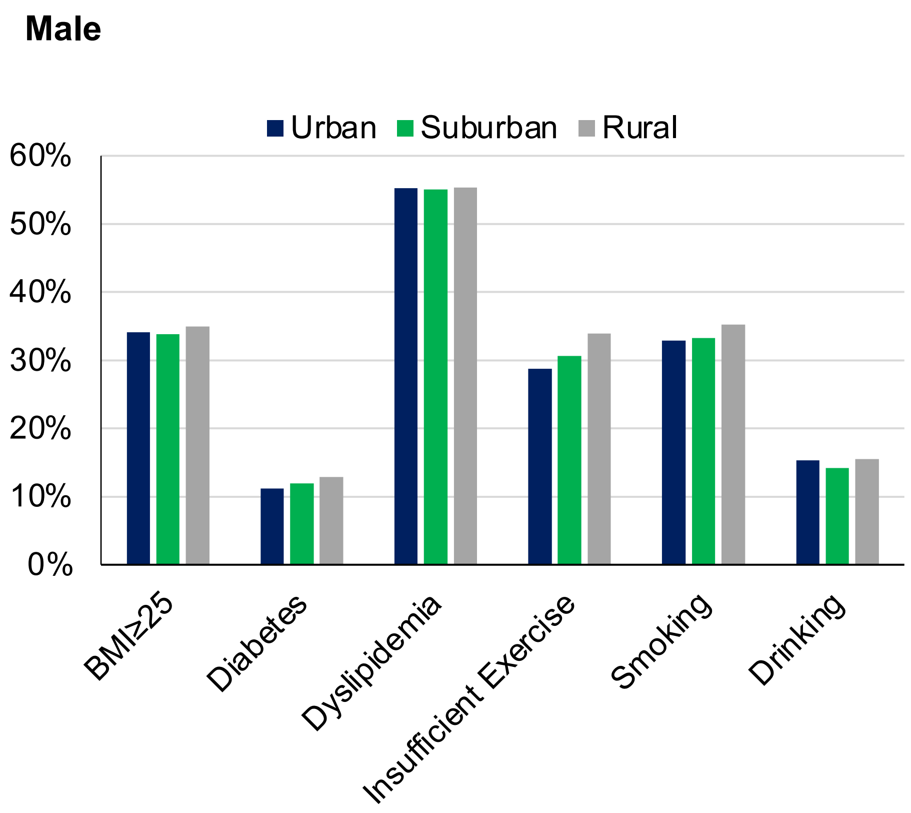

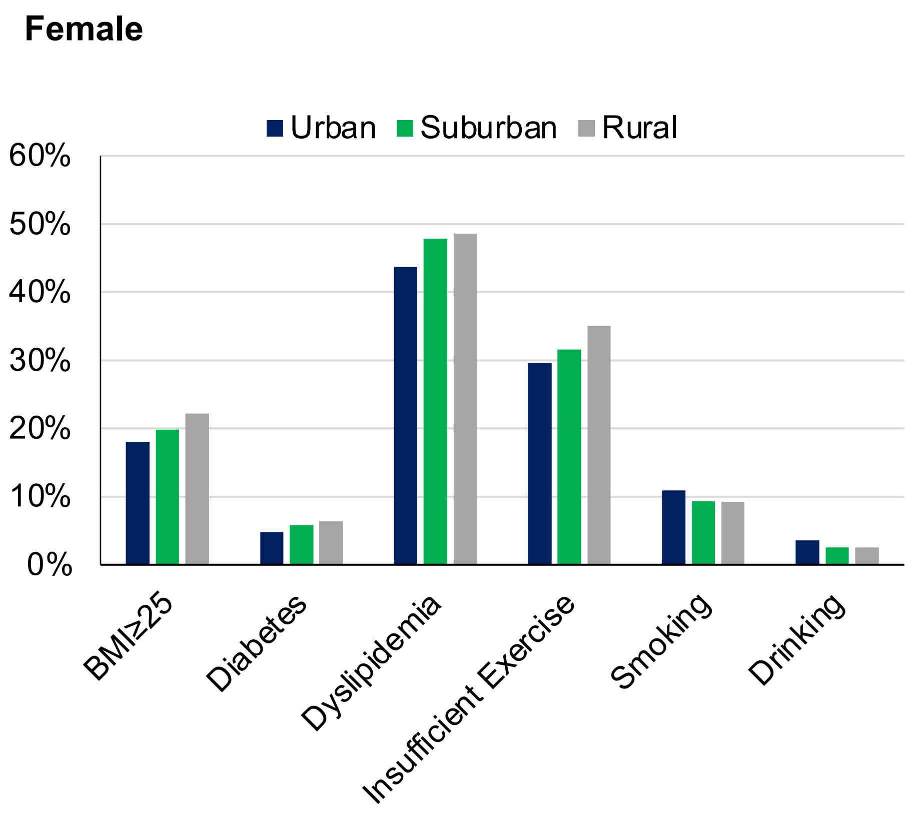

**Supplementary Figure 10. Conceptual Directed Acyclic Graph (DAG) Outlining the Assumed Causal Relationships in Our Adjusted Models**

The adjusted risk ratio estimates the direct effect* of ADI on liver cancer FBSIR/FBSMR, reflecting the impact of ADI through pathways independent of mediators (i.e., obesity, diabetes, dyslipidemia, insufficient exercise, smoking, and drinking). This adjustment was confirmed to be correct in DAGitty (https://www.dagitty.net/). Certain paths that do not directly link the exposure and outcome are represented with dashed lines for visual clarity. ADI, Areal Deprivation Index; FBSIR, fully Bayesian estimate of standardized incidence ratio; FBSMR, fully Bayesian estimate of standardized mortality ratio.

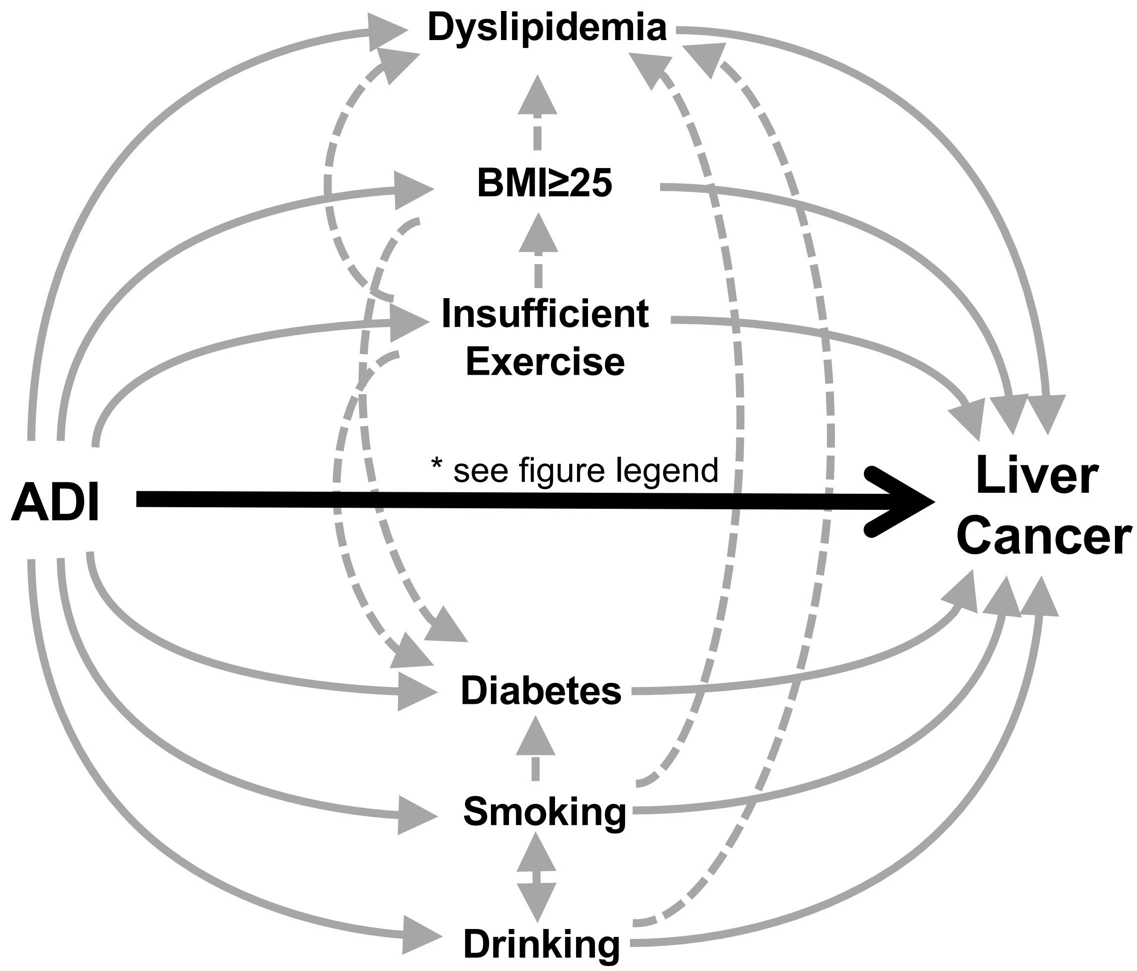
